# Cognitive Reference Charts for Precision Psychiatry

**DOI:** 10.64898/2026.11.26.26369065

**Authors:** Charlotte Fraza, Barbora Rehak Buckova, Lea C. Michel, Stijn de Boer, Camilla Bärthel Flaaten, Linn Sofie Sæther, Viktoria Birkenæs, Giulia Cattarinussi, David Shiers, Veenu Gupta, Ole Andreas Andreassen, Lars T. Westlye, Torill Ueland, Paola Dazzan, Andre F. Marquand

## Abstract

Cognitive impairments are typically assessed in mental health research by comparing a person’s score to norms derived from a reference group. However, these norms are dependent on the underlying sample characteristics and constrained by the methods used for their estimation. In this work, we propose a new approach for estimating cognitive reference charts that overcomes these limitations by learning the optimal norms for each sample based on large-scale data. Simulating how different norming methods perform under realistic data conditions, we found that normative models based on Hierarchical Bayesian Regression (HBR) accounted best for site effects and distributional characteristics such as skewness, kurtosis, and heteroscedasticity. We used these HBR normative models to establish a large-scale cognitive sample (N=14,344, age=5-100 years) that allows for accurate assessment of individual deviation scores across the lifespan. We then assembled another HBR model for older adults (N=34,720) to demonstrate the utility of the cognitive reference charts to the wider research community by showing its application in a smaller sample through federated learning. Accompanied by an interactive website, these charts can easily be transferred to new, unseen datasets, enabling the integration of clinical datasets without compromising data security.

## Introduction

Impairments in cognitive performance are a primary feature of many neurological and psychiatric disorders^1,2^ and can be amongst the most sensitive early markers of illness onset^3^. To accurately assess cognitive function at the individual level, specific population-based cognitive norms are used to evaluate an individual’s cognitive score for a given test, thereby determining whether their performance is higher or lower than expected given certain demographic variables (e.g. age and sex). However, despite the widespread application of these norms, their suitability for a given population is rarely assessed. For instance, existing norms are typically derived from ‘WEIRD’ (Western, Educated, Industrialised, Rich and Democratic) samples^4^. Although efforts have been made to mitigate these WEIRD biases by, for example adjusting for language and socioeconomic status^5,6^, or by estimating country specific norms^7^, small sample sizes often limit their ability to capture full population variability^5^. As a result, these norms are often highly localised and biased towards particular demographics, study settings, and the period during which the reference data were acquired. Together, all these factors restrict the generalisability and can lead to severe bias when applied to participants dissimilar to the population used to estimate the norms^8,9^.

Moreover, limited progress has been made in assessing the appropriateness of the methods used to estimate the norms. Currently, two main approaches are used to derive cognitive norms: binning methods and regression-based norming^10–12^, creating discrete or continuous norms, respectively. These methods rely on specific assumptions that are often not tested, such as a linear relationship between covariates and outcomes, normally distributed and homoscedastic residuals, and the use of representative population samples. Moreover, simple binning or linear models cannot accommodate the heterogeneity that arises when combining data across cohorts with differing demographic or study characteristics.

In practice, the cognitive datasets on which these norms are estimated may violate the distributional assumptions of the algorithm used, undermining the validity of the estimated norms. Besides, current large datasets are often aggregated multi-cohort data, especially within brain-behaviour studies^13^, requiring consideration of several layers of heterogeneity across cohorts^14^. In addition, inaccurate normative estimates can disproportionately affect the extremes of the distribution, leading to severe over- or underestimation of the tails, which, in a clinical setting, are often of primary interest^15^. This may lead to misclassification of cognitive impairment severity, with downstream consequences for clinical decision-making, treatment planning, and access to appropriate interventions.

In this paper, we introduce a new approach to construct more accurate, scalable, and generalisable cognitive norms using normative modelling^16^, inspired by the growth chart models quantifying height, weight, or neuroimaging features ^17–20^. The key difference in this method is that we implement a Bayesian machine learning step whereby existing norms are adjusted (‘transferred’) to a target cohort prior to inference. This guarantees that normed scores are properly calibrated to the target population and can also accommodate differences in the test distributions (e.g. language differences or variations in test design). Like classical norms, this still allows us to calculate a z-score or deviation score for every individual from this reference chart while considering relevant covariates or random effects such as age, sex, and acquisition site. Methods based on normative modelling, due to their Bayesian nature, are particularly appealing for this use case. First, they can quantify different types of uncertainty, including epistemic uncertainty (reflecting measurement noise or modelling uncertainty that reduces with increasing sample size) and aleatoric uncertainty (reflecting variation between individuals that does not decrease with sample size). Second, they flexibly capture heteroscedasticity, nonlinearity, and non-Gaussian distributions^21,22^.

This approach also allows models to be transferred or adapted to a local site without the need to share the data, in a federated learning manner^21,22^. Federated learning enables sites to share model parameters without exchanging raw data. This is crucial for developing accurate and generalisable norms as it allows a reference model to be recalibrated for the particular characteristics of a given sample (e.g. a different language, a different test variant, or a different sociodemographic background). Moreover, it allows for decentralised inference, which is important as many clinical cognitive datasets often cannot be shared centrally due to ethical and legal constraints such as General Data Protection Regulation (GDPR) in Europe and the Health Insurance Portability and Accountability Act (HIPAA) in the USA. Lastly, when cognitive assessments are not identical across cohorts, or when there are differences in test administration, scoring procedures, or task versions, the federated learning framework still allows for the normative model to be applied to these datasets. Thus, the flexibility of this method and the use of federated learning make these normative models particularly suited for deriving cognitive reference charts that are generalisable and adaptable across multiple datasets with complex distributional shapes and potential data-sharing constraints.

We refer to these normative models as *cognitive reference charts*. These charts are first fit using a large cognition dataset representative of a population level using normative modelling techniques, then recalibrated (e.g. transferred) to independent samples using independent data within a Bayesian machine learning framework. The charts are designed to overcome the limitations of traditional estimated small-sample norms from single sites.

In this paper, we: (i) demonstrate the advantages of cognitive reference charts relative to the binned and regression-based norming approaches through a simulation-based analysis; (ii) introduce cognitive reference charts derived from large-scale population-level cognitive data aggregated across multiple cohorts, using Hierarchical Bayesian regression (HBR) (Figure 1). Next, we (iii) show their potential clinical utility by demonstrating how these charts can be used to assess deviation scores for individuals with symptoms of psychosis. Afterwards, we (iv) illustrate their federated learning utility by demonstrating how norms estimated on a large sample can be adapted to a smaller dataset. We used the UK Biobank dataset to construct a new cognitive reference chart and transferred these norms to the smaller HCP aging dataset, demonstrating how these methods remove potential risks of sharing sensitive data. Finally, we (v) ensure that the cognitive reference charts are accessible to the wider scientific community by making them freely available through an interactive online platform that allows users to visualise population trajectories for selected cognitive tests and download the accompanying reference files to compute deviation (z-)scores for their own datasets.

**Figure 1.**
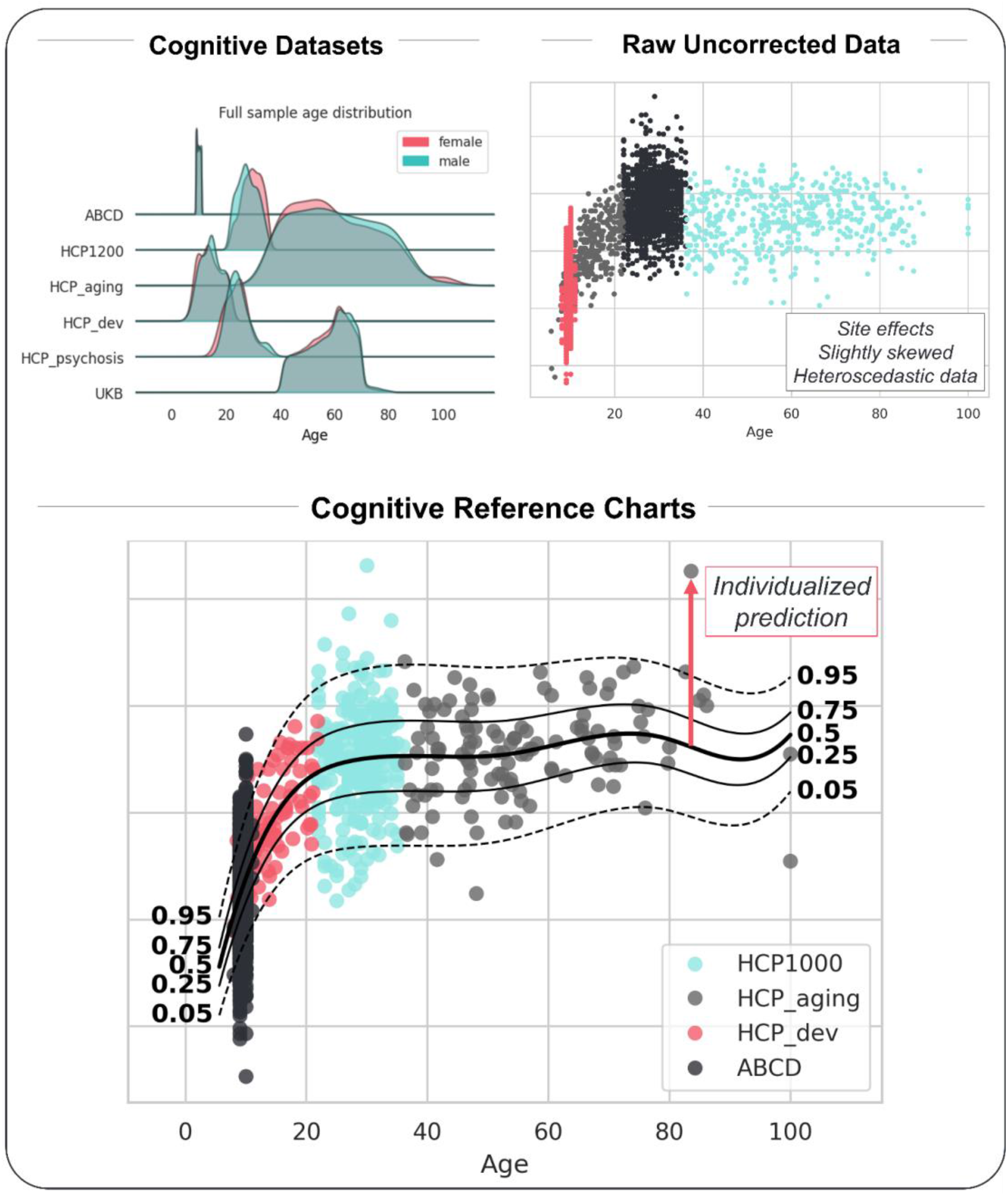
Estimation of the cognitive reference charts: (Top Left) Datasets included in the cognitive reference charting analysis datasets (HCP1200^23,24^, HCP Development^25^, HCP Aging^26^, UK Biobank^27,28^, HCP Early Psychosis^29^, and ABCD^30^). (Top Right) Example of uncorrected cognitive data illustrating site effects, skewness, kurtosis, and heteroscedasticity. (Bottom) Example of a cognitive reference chart based on hierarchical Bayesian methods, which removes batch effects and accounts for distributional differences across datasets, resulting in normally distributed individual deviation scores.

## Results

### Simulation

To illustrate our approach, we first simulated a cognition dataset, with 500 data points (or patients, with 1% patient rate corresponding to 5 patients) for different scenarios representing the severity of site effects, heteroscedasticity, and residual distribution shape. These 3 factors were modelled with three degrees of severity (e.g. none, mild, and strong site effects), resulting in 27 scenarios. These scenarios produced a total of 13,500 simulated observations. We derived cognitive norms for each scenario through three approaches: standardised binned norms, regression-based LOESS norms (i.e. non-parametric regression)^31^, and our generated HBR norms with a Gaussian likelihood distribution^21,32^.

In general, a more flexible HBR model can be fitted to match the different simulated scenarios (e.g. adapting model parameters to the extreme edge cases), as discussed in the methods section. However, here we decided to keep in line with the chosen HBR model settings (see *Simulation study details* in Methods) based on real cognitive data to avoid overfitting for edge cases in our simulation, which would not accurately represent most cognitive samples.

The HBR normative modelling methods outperform both the standardised binned norms and the regression-based LOESS norms approaches by accurately correcting for site effects, skewness, and heteroscedasticity. Figure 2 shows the results for the raw data, which include simulated site effects (mild), heteroscedasticity (mild), and a skewed distribution. The most important feature of model fit is calibration (e.g. whether the estimated centiles match the observed centiles). This can be visualised using Q-Q plots. In Figure 2, the Q-Q-plots illustrate that only the HBR method produces approximate z-scores (e.g. fully standardised scores following a normal distribution).

**Figure 2.**
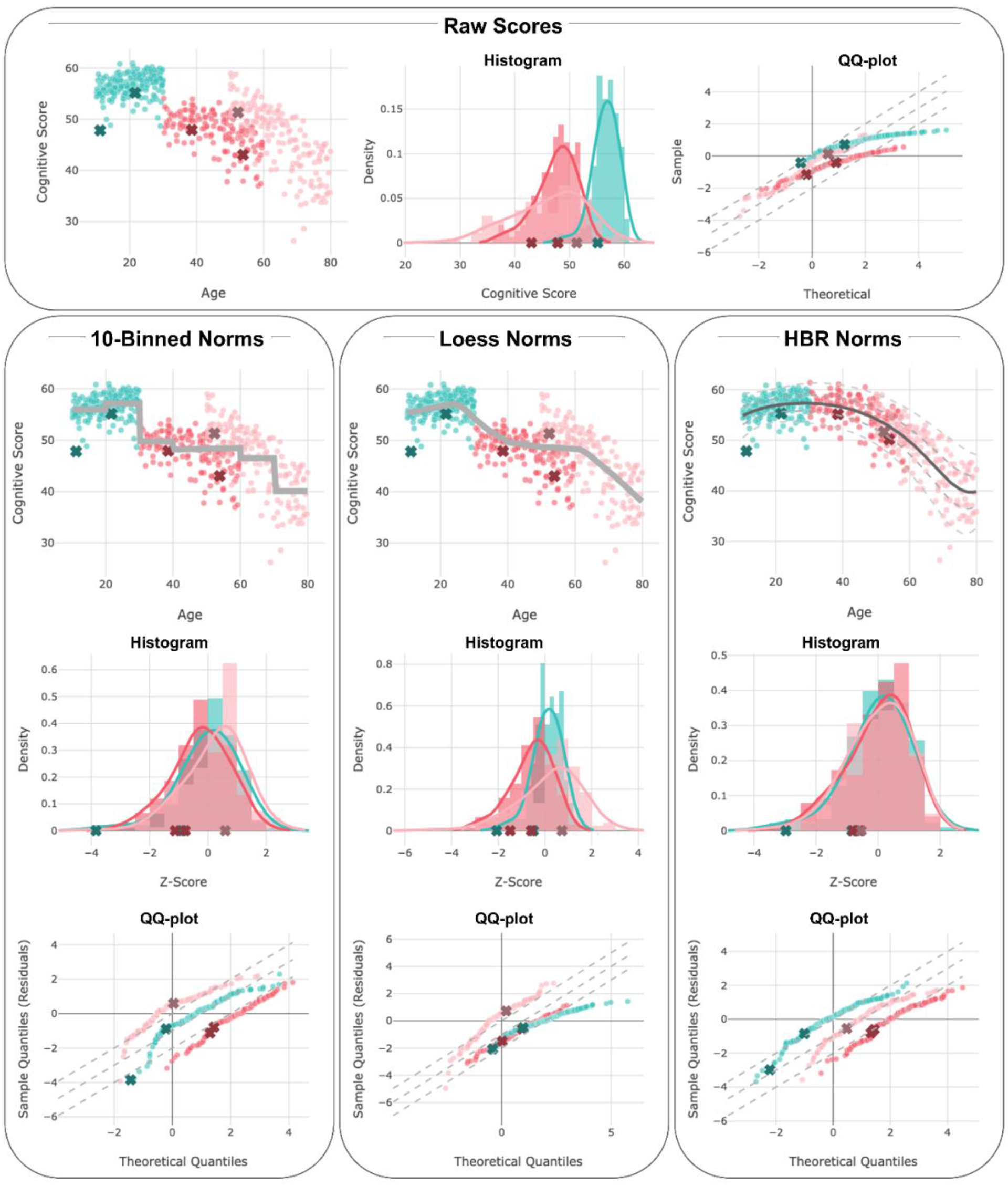
Comparing the performance of different norming approaches; Here applied to simulated cognition scores with characteristics commonly observed in cognitive data, including site effects, skewed distributions, and heteroscedasticity. Three norming methods are evaluated: 10-year binned norms, LOESS-regression-based norms, and Hierarchical Bayesian Regression (HBR) norms. The comparison includes scatterplots and histograms of the resulting normed scores, as well as Q-Q plots comparing the normed scores to theoretical quantiles to assess normality. Simulated patient data is indicated with an “x”. See also for an interactive version of all the plots.

Conversely, binned norms methods lead to discontinuous estimates that can misestimate individual scores if an individual falls between two bins (see binning effect in Figure 5). For example, while they might be estimated as deviant to one particular bin, if they were moved up one age bin, they might be considered within the norm. Using the wrong norms, for example, those not taking into account site effects or heteroskedasticity, can further over-or underestimate the actual position of individuals, and all these factors can interact with each other in deciding where someone should be placed.

We provide additional results in the Supplementary Material. In Supplementary Table 1, we show the model evaluation statistics for all the different simulation scenarios. In Supplementary Figure 2, we highlight the case where, in the presence of strong site effects, LOESS regression norms can overfit. In Supplementary Figure 3, we highlight how certain model evaluation criteria (e.g. R^2^) are dependent on the noise distribution, and in these cases, the HBR model can fit well according to model evaluation criteria assessing normality of the deviation scores, but the R^2^ can be low due to a low signal-to-noise ratio.

For didactic purposes and to highlight the importance of well-calibrated norms, we provide an interactive simulation on the website designed to stress-test norming approaches under realistic distributional violations: https://predictive-clinical-neuroscience.github.io/Cognitive_Charts/. The website gives access to the cognitive normative models developed to assess individual cognitive performance relative to the population-based reference curves. Users can explore various covariates settings and assess how the cognitive scores vary across age and sex. Furthermore, the normative curves are shown for both the simulation dataset and all the features of the publicly available datasets we use in the evaluation (see methods). The platform also allows users to compare different norming methods, including traditional binning, LOESS, and the here-proposed HBR method, allowing users to visualise the impact of different methodological choices on the estimated cognitive scores.

### Developing Cognitive Reference Charts for Common Cognitive Tasks

Next, we applied the HBR model separately for the 16 cognitive tests from the National Institute of Health (NIH) battery. Whilst our approach can be used for any cognitive battery, we chose the NIH test battery because it is used widely across studies, many of which provide publicly available data. In more detail, we estimate cognitive reference charts for each NIH variable (Figure 3 and Figure *4*, and Supplementary Figures 4 to 8). For each figure, we show the raw scores, the default age-adjusted scores (‘scaled measures’) for the NIH tests (e.g. those provided out of the box by the NIH toolbox), and the harmonised HBR scores in addition to Q-Q-plots and histograms of both the standard normed scores and the derived z-scores.

**Figure 3.**
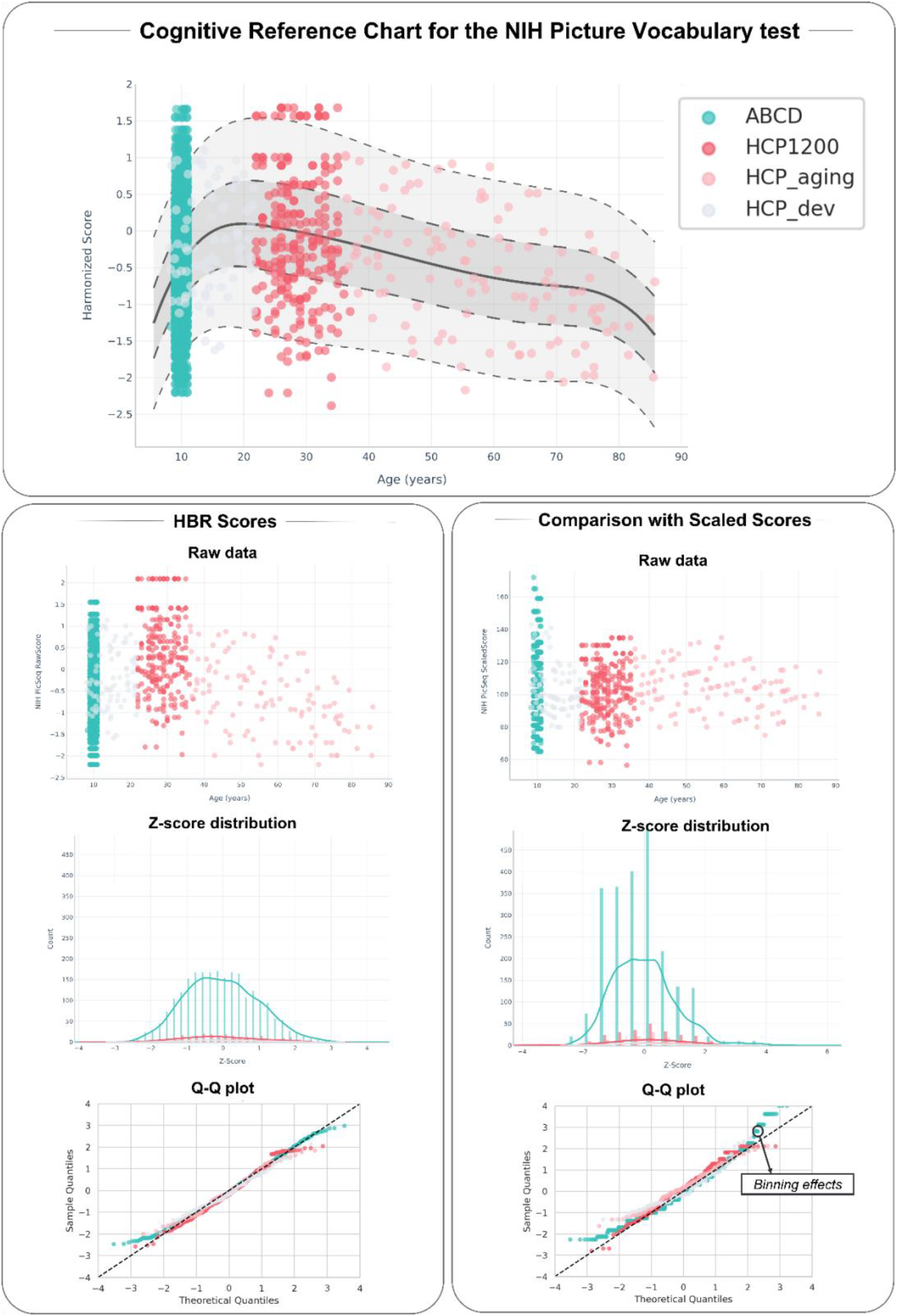
Cognitive reference charts for the NIH Picture Vocabulary test. The top panel shows the cognitive reference chart made with the HBR model. The left panel shows the scatter plot of the raw scores by age, a distributional plot, and Q-Q plot. The right panel presents the scaled scores, including a scatter plot, a distributional plot, and Q-Q plot, highlighting issues with binning.

**Figure 4.**
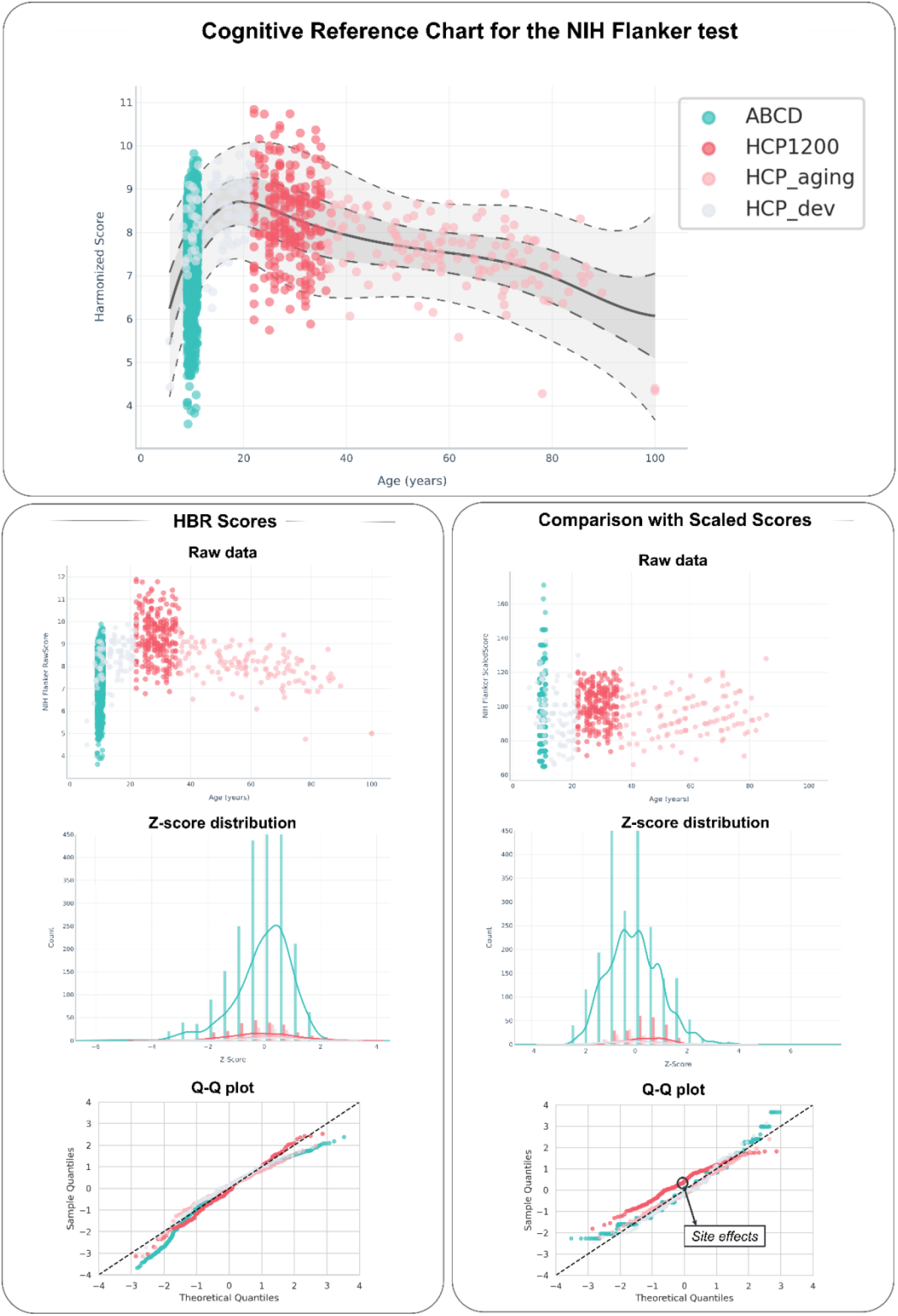
Cognitive reference chart for the NIH Flanker test. The top panel shows the cognitive reference chart made with the HBR model. The left panel shows the scatter plot of the raw scores by age, a distributional plot, and Q-Q plot. The centiles for ABCD remain suboptimally calibrated, probably due to the narrow age range. The right panel presents the scaled scores, including a scatter plot, a distributional plot, and Q-Q plot, highlighting potential site effects.

The results highlight several problems with the default measures. For certain cognitive tests, such as the NIH Picture Vocabulary test (Figure 3), the standardised scores are binned, and therefore the scaled measures are not well calibrated. In contrast, the scores derived from the HBR cognitive reference charts are continuously scored and Gaussian, indicating that the HBR method corrects for skewness, kurtosis, and heteroscedasticity. In other tests, such as the Flanker test (Figure 4), there are slight site effects present that are not fully corrected for in the scaled scores, despite essentially identical versions of the tests being used across studies. The HBR method corrects for these issues, making the results comparable across sites and allowing us to combine the data into a single cognitive reference chart. Moreover, the HBR normative modelling method incorporates uncertainty estimates through the adjustment of the centiles (making it harder to achieve an extreme score under high uncertainty), which is advantageous in age ranges with sparse data (e.g. 80+ years). We also provide the cognitive reference charts for the Rey Auditory Verbal Learning Task in Supplementary Figures 9 to 17, which show similar trends to the NIH cognitive reference charts.

More detailed evaluation metrics for the HBR models are shown in Table showing the mean absolute centile error (MACE^32^): calibration of predicted uncertainty, MSLL: mean standardised log-likelihood, R²: explained variance (accuracy), RMSE: Root Mean Square Error, and the Shapiro-Wilk: for a distributional check or residual normality.

Finally, in Figure 5, we show a pairwise comparison between raw, scaled scores, and HBR z-standardised for the Flanker test scores, see Figure 18 in the Supplementary Material for the comparisons across all cognitive measures from the NIH toolbox included in the datasets, highlighting again common issues that arise in the scaled scores due to binning or site effects. Here, we can observe the effects of binning, especially in the ABCD data (in blue), where different bin sizes were used for different age ranges. We also observe clear site effects (e.g. non-overlapping site-specific regression lines) for almost all the datasets present in both the raw and the scaled scores, showing that if these effects are not taken into account, they permeate into the raw and scaled scores. Notice also that these effects are consistent across the age range (e.g. constant offset over time), and can introduce distortions such that the adjusted test scores have different psychometric properties than the raw scores for different age ranges.

**Figure 5.**
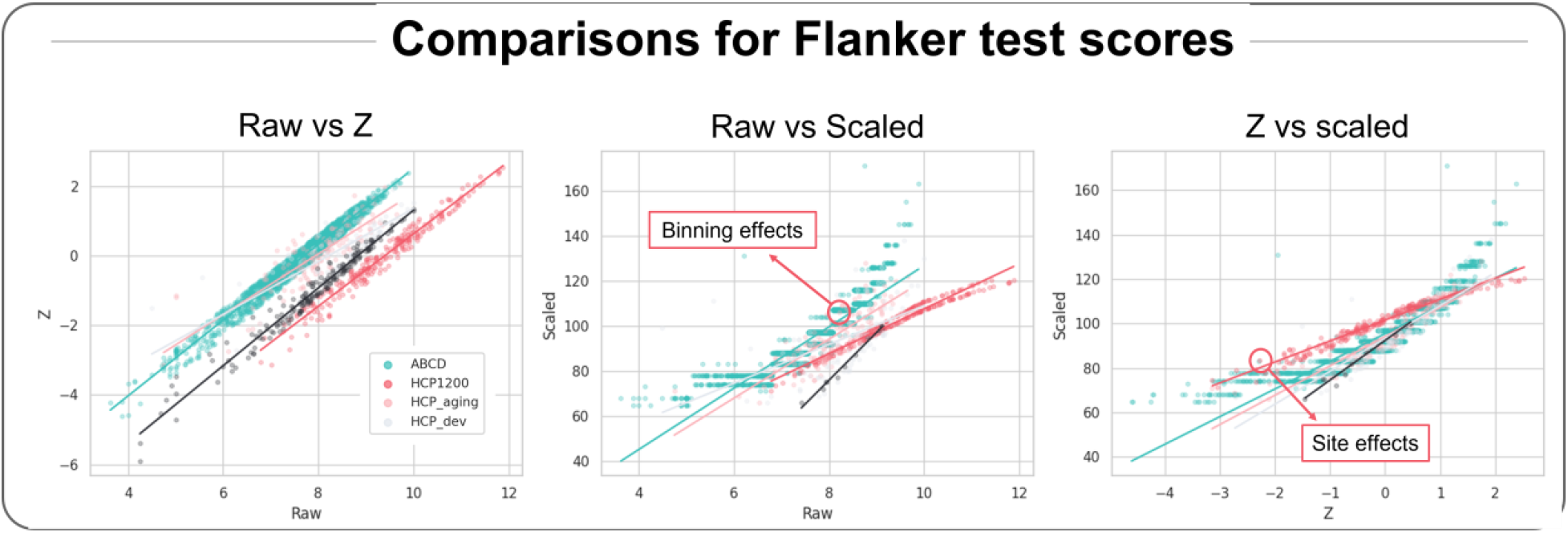
Pairwise comparisons between raw, HBR z-standardised, and scaled scores for the Flanker cognitive measures (see Supplementary Figure 31 for the other tests). Each panel shows the relationship between two scoring formats for a given cognitive measure. Pearson correlation coefficients (r) and associated p-values are reported in each panel. Binning effects can be seen by the step-like appearance of the regression line. Site effects can be observed by the shifts between the regression lines for different datasets.

### Applying Cognitive Reference Charts on Psychosis Sample

Next, we demonstrate how large-scale cognitive reference charts can be applied to a new data sample. We choose the Human Connectome Project (HCP) Early Psychosis sample to highlight future clinical application of the cognitive reference charts. Site effects were taken into account by incorporating the healthy controls from the HCP Early Psychosis dataset (the new site) into the training sample used to construct the cognitive reference charts.

The results show that across all cognitive domains, individuals with psychosis demonstrated significantly lower performance than expected based on the normative reference population, between -0.510 and -1.127 depending on the test. These results align with differences found in the literature based on other norming methods^33–35^. The top panel in Figure 6 shows the cognitive reference chart for the NIH Card Sort test and its subsequent application to the patient sample to estimate individual deviation scores. The middle panel provides an overview of the estimated HBR Z-scores across cognitive measures for patients, showing that the distributions for most cognitive domains are shifted away from the expected mean value of zero.

**Figure 6.**
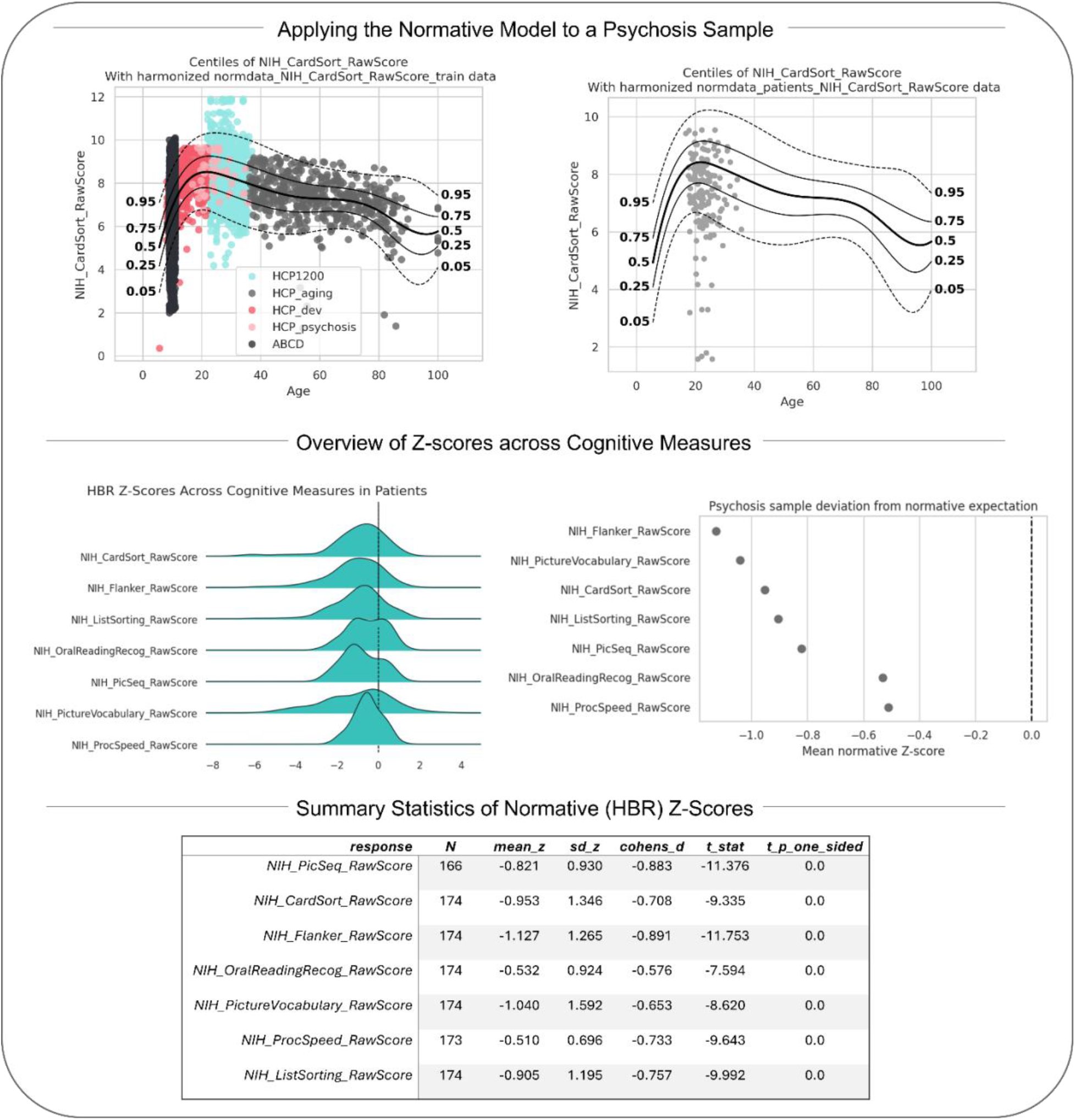
Application of the normative model to HCP Early Psychosis sample. Top: Example normative modelling results for the NIH Card Sort task. The centile plot shows patient observations projected onto normative reference trajectories across age, while the Q–Q plot assesses the distribution of normative model Z-scores. Middle: Group-level overview of normative Z-scores across cognitive domains. The ridgeline plot illustrates the distribution of patient Z-scores for each cognitive measure, and the forest plot summarises the average deviation from zero across domains. Bottom: Summary statistics of normative model results, including sample size, mean Z-score, standard deviation, effect size (Cohen’s d), and one-sample t-test statistics.

### Federated Cognitive Reference Charts

Next, we demonstrated that the cognitive reference charts can be adjusted to new data without requiring the transfer of raw data through an additional learning step: federated learning^22^ (i.e. decentralised learning). Using the HBR approach, this can be done in two ways: ‘transfer’ or ‘extend’ functions (see Methods for more information). In this work, we focused principally on the transfer function as it is the one most likely to be adopted in practice, see these papers^21,22,32,36^ for further details.

To show this, we fit two cognitive reference charts for the UK Biobank Trail Making Tests A and B. We then apply these models through federated transfer learning to the same tests in a previously unseen dataset, HCP-Aging. The Trail Making Test is an ideal choice for this because it is highly sensitive to variations such as the mode of administration (e.g. paper or electronic tablet-based) in different datasets. Accordingly, notice that the scales of the scores are not consistent across both datasets (Figure 7.2, Raw data), which means that the norms from UKB cannot be directly applied to HCP-Aging. In Figure 7, we show the fitted centiles of the transfer function for the Trail Making Test Part A, plotting the transferred chart retrained with HCP aging data. After transferring the model, we show that the model is well-calibrated, as the resulting deviation distribution matches the theoretically assumed Gaussian distribution on the Q-Q plots, and thus we can infer the HCP aging z-scores based on the transferred UKB norms. In Table 2, the model evaluation statistics are shown for the original HBR model trained on the UKB and transferred for the HCP aging datasets.

**Figure 7.**
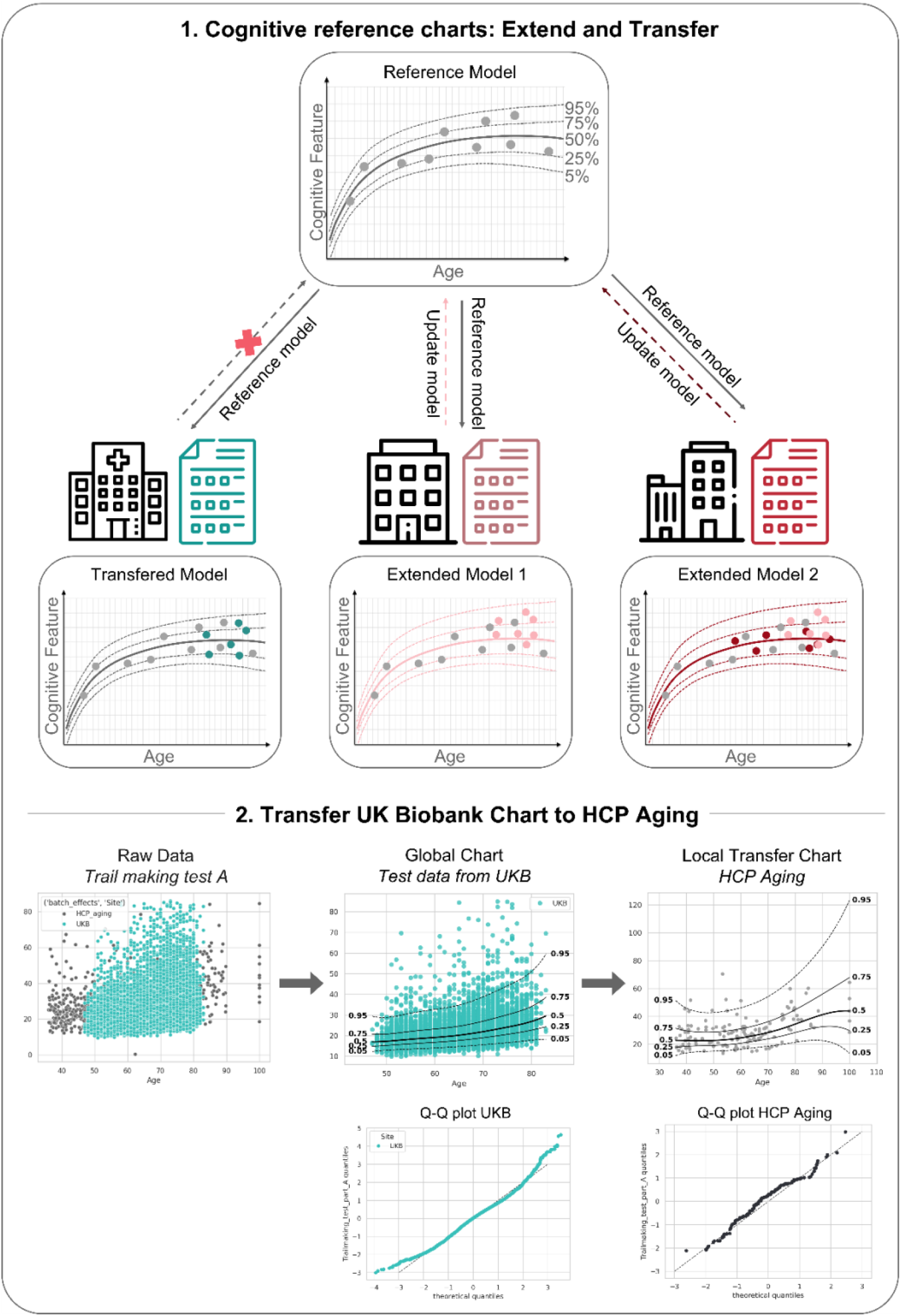
Federated learning model proof of principle. **(1)** An explanatory schematic illustrating two methods of federated learning: a transfer method, in which the original model is not updated with new data, and an extension method, in which the reference model is updated using the new data. **(2)** Results of the transferred model for the Trail Making Test A (see results for Trail Making Test B in the Supplementary Figure 31). The first panel shows scatterplot of the raw data for UKB and HCP-Aging. The second panel shows the estimated cognitive reference chart for the test based on the UKB dataset. The third panel shows the model transferred to the unseen HCP aging dataset.

**Table 1.** Model evaluation criteria for the HBR model with a normal likelihood showing the R²: variance explained (accuracy), RMSE: Root mean square error, MSLL: model fit and uncertainty, MACE: calibration of predicted uncertainty, and the Shapiro-W: distributional check (residual normality).

| ***statistic*** | ***MACE*** | ***MSLL*** | ***RMSE*** | ***R2*** | ***ShapiroW*** |
| ***variable*** |  |  |  |  |  |
|---|---|---|---|---|---|
| *NIH_PicSeq_RawScore* | 0.021 | 0.143 | 0.786 | 0.081 | 0.995 |
| *NIH_CardSort_RawScore* | 0.021 | -0.487 | 1.119 | 0.309 | 0.957 |
| *NIH_Flanker_RawScore* | 0.025 | -0.125 | 0.932 | 0.182 | 0.956 |
| *NIH_OralReadingRecog_RawScore* | 0.015 | -1.809 | 2.620 | 0.560 | 0.984 |
| *NIH_PictureVocabulary_RawScore* | 0.017 | -1.227 | 1.739 | 0.517 | 0.984 |
| *NIH_ProcSpeed_RawScore* | 0.004 | -2.542 | 10.994 | 0.099 | 0.998 |
| *NIH_ListSorting_RawScore* | 0.010 | -1.249 | 3.109 | 0.089 | 0.983 |
| *RAVLT_ShortDelay_TotalCorrect* | 0.020 | -2.669 | 14.401 | 0.029 | 0.963 |
| *RAVLT_ShortDelay_I_TotalCorrect* | 0.014 | -0.602 | 1.759 | 0.027 | 0.995 |
| *RAVLT_ShortDelay_II_TotalCorrect* | 0.009 | -0.844 | 2.257 | 0.034 | 0.998 |
| *RAVLT_ShortDelay_III_TotalCorrect* | 0.019 | -0.992 | 2.568 | 0.034 | 0.992 |
| *RAVLT_ShortDelay_IV_TotalCorrect* | 0.019 | -0.998 | 2.640 | 0.027 | 0.976 |
| *RAVLT_ShortDelay_V_TotalCorrect* | 0.032 | -0.981 | 2.564 | 0.032 | 0.954 |
| *RAVLT_ShortDelay_B_TotalCorrect* | 0.018 | -0.543 | 1.701 | 0.024 | 0.992 |
| *RAVLT_ShortDelay_VI_TotalCorrect* | 0.024 | -1.134 | 2.999 | 0.026 | 0.978 |
| *RAVLT_ShortDelay_VII_TotalCorrect* | 0.014 | -1.166 | 3.252 | 0.019 | 0.984 |

**Table 2.** Statistics for the HBR model trained on the UKB (top) and the HBR model trained on the UKB and transferred for the HCP aging dataset (bottom) with a SHASHb likelihood showing the R²: variance explained (accuracy), RMSE: Root mean square error, MSLL: model fit and uncertainty, MACE: calibration of predicted uncertainty, and the Shapiro-W: distributional check (residual normality).

| ***variable*** | ***Trail Making Test part A*** | ***Trail Making Test part B*** |
|---|---|---|
| ***statistic*** | *HBR model trained on UKB* |  |
| *MACE* | 0.013 | 0.012 |
| *MSLL* | -2.395 | -3.581 |
| *RMSE* | 7.012 | 23.717 |
| *R2* | 0.128 | 0.126 |
| *ShapiroW* | 0.992 | 0.991 |
| ***statistic*** | *HBR model trained on UKB and transferred to HCP aging* |  |
| *MACE* | 0.095 | 0.059 |
| *MSLL* | -2.055 | -3.517 |
| *RMSE* | 10.446 | 36.880 |
| *R2* | 0.124 | 0.168 |
| *ShapiroW* | 0.944 | 0.985 |

Further, we highlight the ability within this framework to allow users to choose the degree to which they expect their local sample to reflect the demographics and test parameters of the original cognitive reference charts. In other words, how close are the characteristics of the user’s dataset compared to the datasets used to compute the reference charts? This can be achieved using a so-called ‘freedom’ parameter in the HBR implementation, which allows tighter or more flexible priors (see Methods). In Supplementary Figure 31, we show the results of the transfer function under three different settings for the freedom parameter. If the datasets differ substantially, one can choose a more liberal prior, placing less weight or confidence on the original cognitive reference charts and allowing the model to learn more from the local data; vice versa in the opposite scenario.

Lastly, to highlight the benefit of using a transfer function, the model trained on the UKB dataset was applied to the HCP-Aging dataset before and after transfer learning (Supplementary Figures 32 and 33). The first scenario is similar to applying pretrained normative reference charts to a new dataset, a situation common in cognitive analyses. When the UKB-trained model was applied directly to HCP-Aging data without adaptation, the centile and QQ plots revealed expected systematic (site-) deviations between the HCP-Aging data and the cognitive reference charts. In the second scenario, in which the model was adapted through federated transfer learning using a subset of HCP-Aging participants in the training dataset, these systematic site differences were resolved, and the Q-Q and centile plots showed no apparent deviations between the transfer test data and the cognitive reference charts. These findings suggest that transfer learning successfully accounted for site- and cohort-specific differences, resulting in better calibrated normative estimates in the independent HCP-Aging sample.

## Discussion

In this work, we propose a novel approach to derive cognitive reference charts via the combination of multiple large-scale cognitive datasets and a Bayesian machine learning approach that allows data to be explicitly recalibrated to a target dataset before inference. These models provide newly estimated norms that can be readily used by the research community to estimate deviation or z-scores for a given cohort, accounting for sample characteristics such as age, sex, and site. We first highlight subtle errors that can be present in standard normed scores (e.g. binned and LOESS norms) through a simulation study. Next, we present real-world examples of how standard normed scores are often miscalibrated if used ‘out-of-the-box’ and how this can be corrected using Hierarchical Bayesian Regression methods. Third, we show that cognitive reference charts compared favourably to the standard normed scores, achieving equivalent or better calibration norms for several cognitive tests. Finally, all models discussed here are freely available online https://predictive-clinical-neuroscience.github.io/Cognitive_Charts/, along with the underlying code to derive them. Finally, we highlighted the flexibility of using these cognitive reference charts to be adapted to different populations or test versions in a federated learning framework that can be used without sharing data. Allowing users to update model parameters or adapt the models to their local datasets without sharing data or compromising data privacy.

Here, we move beyond static cognitive norms based on binning or standard regression-based methods and establish cognitive reference charts using a flexible hierarchical Bayesian regression framework, which provides several advantages. First, it provides scalability by allowing us to combine data from multiple datasets by including site-specific effects, enabling the model to account for variations in cognitive tasks in addition to language-, local noise-, and other site-related effects. This enabled us to build a much larger, representative and therefore more generalisable baseline model. Second, the model explicitly accounts for non-linearity and non-Gaussianity in the relationship between the covariates and the outcome measures, enabling a more accurate approximation of standardised z-scores compared with binned or standard regression-based scores. Finally, using the HBR methodology, we can make use of the federated transfer and extension functionalities, which allow us to adapt the model to smaller, new sites that may exhibit subtle differences from the original datasets^21,32^. These differences can explicitly be taken into account in the transfer function through the freedom parameter, which allows us to place a tighter or more liberal prior on the derived estimates. This enables users to apply the models without assuming that the cognitive features are identically estimated, as shown in the example presented here, where there were differences in the administration procedures for the cognitive tests of the UK Biobank and the HCP aging test. Another crucial point is that it is essential that the evaluation of cognitive norming strategies – regardless of what they are – is performed out of sample (i.e. on independent data). This is widely regarded as good practice within the machine learning literature but is not always implemented in the cognitive domain. In the case of cognitive reference charting, we propose to use two splits of the data, where a portion of the data is used to recalibrate the norms (‘transfer’ or ‘adaptation data’) prior to evaluation on the remaining portion (‘test data’). In practice, the amount of data used for adaptation can be minimised, leaving as much as possible for downstream inference. This has been explored in more detail in the normative modelling literature^37,38^.

Different methods are available to produce cognitive charts, yet their application depends on the characteristics of the sample. We performed a model comparison across different versions of the HBR and BLR methods available in the PCNtoolkit (Python package used in our analyses), demonstrating that an HBR model with a Gaussian likelihood for the NIH and RAVLT tests is sufficiently flexible to capture most of the non-linearity and site-related offsets present in cognitive datasets. For the Trailmaking tests, an HBR with a SHASHb likelihood was preferred (see methods), which is capable of modelling the non-Gaussianity of the distribution.

Several other approaches for creating reference charts exist, mostly methods created for height and weight reference charts^39^, as well as neuroimaging reference charts^16^. Quantile-based methods, for example, are quite common in height and weight charts, but do not prevent population centiles from crossing as a function of age, which is usually undesirable^40^. For growth charts, the “LMS method” is currently applied by the World Health Organization^17,39^, and proposed extensions such as GAM and the GAMLSS^41^ framework have also shown to have strong potential for improving cognitive norms^10^. Recently, an updated set of norms has been released for the NIH Toolbox V3, based on GAMs^42^, resulting in substantial renorming and rescoring of the cognitive measures. These models share similarities with the HBR approach presented here in that they can flexibly handle non-linearity and non-Gaussianity. For more details on the new NIH norming approach, see the V3 Technical Manual of the NIH Toolbox^42^.

One downside of both the LMS and the GAMLSS framework is that they are not truly Bayesian in nature, which may limit straightforward integration with certain Bayesian federated learning approaches, an important consideration given the often sensitive nature of cognitive data in patient populations. Therefore, mapping the original V2 scores (based on discrete age groups)^43^ to the V3 scores is not straightforward, making it currently impossible to merge older and new cognitive datasets. Furthermore, many of the other problems we highlight (e.g. site effects) may further persist when these static norms are applied to independent and local data. New algorithms are being developed to allow for federated learning in GAMLSS^44^, but they are not widely implemented yet. Additionally, the creation of large and accurate, site-specific norms can be highly costly and time-consuming; therefore, a strategy where global norms are pre-estimated once and subsequently transferred to specific sites could substantially reduce both computational and financial burdens and create the possibility of having more accurate norms, especially for local samples.

A strong argument in favour of adapting general norms for each application relates to the complexity of cognitive measurements themselves. That is, cognitive features quantify an inferred construct, such as processing speed or episodic memory, rather than a directly measurable property (e.g. height and weight). When combining different cognitive data that aim to measure the same construct across sites, we assume that, to some degree, the same underlying construct is being assessed. However, even small differences in cohorts or cognitive test administration can lead to substantial differences in the resulting population distributions. These differences could arise from language effects^45^, cultural and socio-economic differences^46^, or other cohort-specific characteristics, as well as differences in test administration (tablet vs. paper)^47^, and other specific task parameter choices. All of these features influence the individual raw scores and may misrepresent the z-scores of one specific (clinical) sample.

In principle, cognitive test batteries have been designed to minimise these differences and the NIH Toolbox now actively corrects their norms based on U.S. demographics^5^. However, certain open-source datasets still employ slightly modified versions of standardised tests, as is the case for the UK Biobank data used in this paper. Our results show that these data are not well calibrated ‘out of the box’ and cannot easily be used for different datasets. Even though the UK Biobank Trail Making Test has been shown to correlate reasonably well with the original tests assessing the same cognitive domain^48^, it remains unclear how an individual’s position within the population cohort is affected. We have demonstrated that, in theory, all of these tests and scores can be combined within a cognitive reference chart, taking into account site effects, and have done so for features that we consider reasonably overlapping, as well as testing the same constructs. The variance observed between sites is then modelled and removed as the general variance attributable to site effects. However, the question of whether tests are comparable enough to be merged remains open and should always be considered before transfer.

Another debated topic within normative modelling and cognitive reference charting is the inclusion of more specific covariates^9,49–51^. While variables such as site, age, and sex are included here, these are considered minimally corrected norms. Additional covariates, such as years of education, socioeconomic status, and race/ethnicity are also often considered; these would then provide demographically corrected norms^50^. In this study, we decided on minimally corrected norms as the standard for the cognitive reference charts for two reasons. First, retrospective harmonisation is challenging because datasets differ in their definition of certain covariates (e.g. varying definitions of race/ethnicity across sites) or may have entirely neglected to measure them. Second, our goal for the cognitive reference charts is broad applicability to various hypotheses, and adding more covariates would reduce usability if these variables were not consistently measured or of interest to the study in question.

Fundamentally, the exclusion or inclusion of a specific covariate hinges on the question of what should be regarded as a meaningful signal, and which sources of variance should be treated as noise and adjusted for in the cognitive reference charts. For example, when assessing whether an individual’s cognitive performance is typical for their age, factors such as preferred language could systematically lower scores for certain groups^45^. However, in predictive modelling settings, such as identifying individuals at risk for developing a psychiatric disorder, correcting for variables like socioeconomic status may remove meaningful signal, as these factors could correlate with both cognitive performance and the risk associated with the disorder (see also a similar argument regarding race-corrected norms in hypertension^52^). Consideration could also be given to the potential of insights from those with psychosis in patient and public involvement roles and how their perspectives could be integrated to inform norms development and what they regard as meaningful signal.

There is a consensus that general norms should, over time, represent the population in all its heterogeneity. However, here, and in most psychological and neuroimaging datasets, most of our samples are from WEIRD^4,53^ countries, reducing the potential issues that might arise from combining very diverse populations. With time, we hope that through the addition of new, more heterogeneous datasets, we can create more representative norms that then can be adapted to local datasets when needed. An individual could then be compared to a local transferred norm (within one’s state, country, or language) that also contains the information through the prior of the general cognitive reference charts or to a global norm (cross-country, global cognitive reference charts). Through this setup, more flexibility as well as nuance can be given to the now static norms. Overall, the cognitive scores both presented here and in other studies are indicative rather than fully representative of an individual and must always be interpreted with care, considering one’s historical, sociodemographic, and cultural context^54^.

With these considerations in mind, we constructed the cognitive reference charts with scalability across different contexts. We showed how the federated learning framework could be used in general to decide on a weak or strong informative prior, depending on the match between the cognitive reference sample and the local sample. The freedom parameter feature allows the model to leverage information from the more general cognitive reference models while adapting to local site effects, including potential cultural and sample-specific characteristics. This adaptation must be applied with care, as the site effects are currently unspecified and may reflect a multitude of factors, including potential meaningful local variance that could inadvertently be removed. In future work, we plan to explore this in more detail and further improve the models by taking into account longitudinal trajectories. We also plan to investigate the cognitive deviation scores of individuals with psychiatric disorders such as psychosis. Here, we have shown through a simulation that the misestimation of the norms can be detrimental for the tails, where our patients often lie. However, we have not discussed this fact using more specific samples, and how we can both under- and overclassify patients versus controls if, for example, we create a predictive algorithm on the basis of our cognitive features.

In summary, we have presented a flexible and generalisable framework for cognitive norms that involves estimating a cognitive reference chart from large scale data, then generalising this to the characteristics of a given local sample using a Bayesian normative modelling framework. This guarantees that the norms are well-calibrated for the target dataset, even if the sampling characteristics or test variant are different. This can also be achieved without sharing data, via federated learning. Finally, we provide norms suitable for a range of widely used measures via a software portal. We recommend the cognitive reference chart framework as a promising approach for generating accurate, scalable, and generalisable cognitive norms.

## Methods

### Simulation study details

For the simulation study, we generated multiple scenarios designed to capture the range of distributional shapes observed in real-world cognitive datasets. The data generation process modelled cognition as a function of age, site, and sex. In addition, three factors were modelled with varying degrees of severity, resulting in 27 distinct scenarios: site effects (none, mild, strong), heteroscedasticity (age-dependent noise: none, mild, strong), and residual distribution shape (Normal: Gaussian noise; Skewed: right-skewed via a skew-normal distribution; Kurtotic: heavy-tailed errors via a Laplace distribution; see Figure 2).

For each subject *i*, the observed cognitive parameter *Y_i_* was generated according to

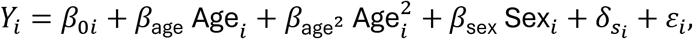

where *β*_0_*_i_* denotes the subject-specific intercept, and *δ_si_* captures site-specific effects for site *s_i_*. The coefficients were fixed to *β*_age_ = 0.4, *β*_age_2 = −0.0067, and *β*_sex_ = 0.2. The residual term *ε_i_* represents random noise and was drawn from distributions with scenario-dependent variances and shapes. For implementation details, also see the added code section in the Supplement.

For each scenario, we applied three different normative approaches, including the proposed HBR normative modelling method and two commonly used cognitive norming methods, binning and regression^55^:

- **Binning by 10**: Traditional 10-year age bins for which the mean and standard deviations are calculated.
- **Regression LOESS**: Non-parametric LOESS regression with a smoothing parameter of 0.3, producing smooth regression age–dependent functions^31^.
- **HBR**: Hierarchical Bayesian Regression with age, site, and sex as covariates, providing both mean and centile estimates (c5, c25, c50, c75, c95)^21,22,32^.

The HBR model was implemented here with standard settings^21^ as implemented in the PCNtoolkit software v1.1.2^32^. This included a Gaussian likelihood, although more flexible modelling options are available (see ‘Normative model specifications’ below for details on alternative likelihood options for the HBR method). Furthermore, the HBR model included site and sex as batch effects and modelled non-linearity using a B-spline function. The models were fitted through posterior sampling via MCMC with 4 chains, 1500 draws, and 500 tuning iterations. Finally, to assess how errors in norming approaches propagate to clinical populations, we simulated an additional subgroup of “patients” with a 1% prevalence rate in the population (indicated with an X in Figure 2). These data points had the same age, sex, site, and noise characteristics as the controls, but were sampled to show a mild cognitive deficit by introducing a slight reduction in their cognitive scores sampled from a normal distribution (*μ* = 0.5, 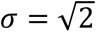). This data generation approach approximates the cognitive impairments observed in psychiatric conditions such as psychosis^56^, and highlights how small misestimations in norming approaches can either exaggerate or diminish deviation scores.

### Data selection and preprocessing

We assembled two large cognitive data samples that had overlapping cognitive tests, one to establish large cognitive reference norms and the second to show the federated learning aspect of normative modelling. The first dataset contained 14,344 individuals with cognitive data from five datasets spanning the human lifespan (5.6–100 years), including data from the following sources: Human Connectome Project 1200 subjects release (HCP1200)^23,24^ (N = 1,206, aged 22–37 years, 54% female), HCP Aging^26^ (N = 720, aged 36–100 years, 56% female), HCP Development^25^ (N = 468, aged 5.6–22years, 54% female), HCP Early Psychosis^29^ (N = 234, aged 16-35 years, 38% female) and ABCD^30^ (N = 11,716, aged 8–11 years, 48% female). For the second dataset, we included data from over 35,576 healthy individuals; we combined data from the UK Biobank^27,28^ dataset (age 47–83 years, N= 34,003, sex 54% female) and HCP Aging (N = 717, aged 36–100 years, 56% female). Table 3 provides the number of participants included in each of the cognitive tests analysed. Summary statistics for each test, including minimum, maximum, mean, standard deviation, skewness, kurtosis, number of extreme outliers removed (z-scores>7), and total N, are provided. For each cognitive variable, the number of participants (N) was slightly lower than the total number of participants, as shown in Table 3, because not every participant completed every cognitive test. Further information on the assessments and inclusion of tests is described in the Supplement. Protocol-specific details for each cohort are available in the original cohort publications^25,26,29,57–59^.

**Table 3.**
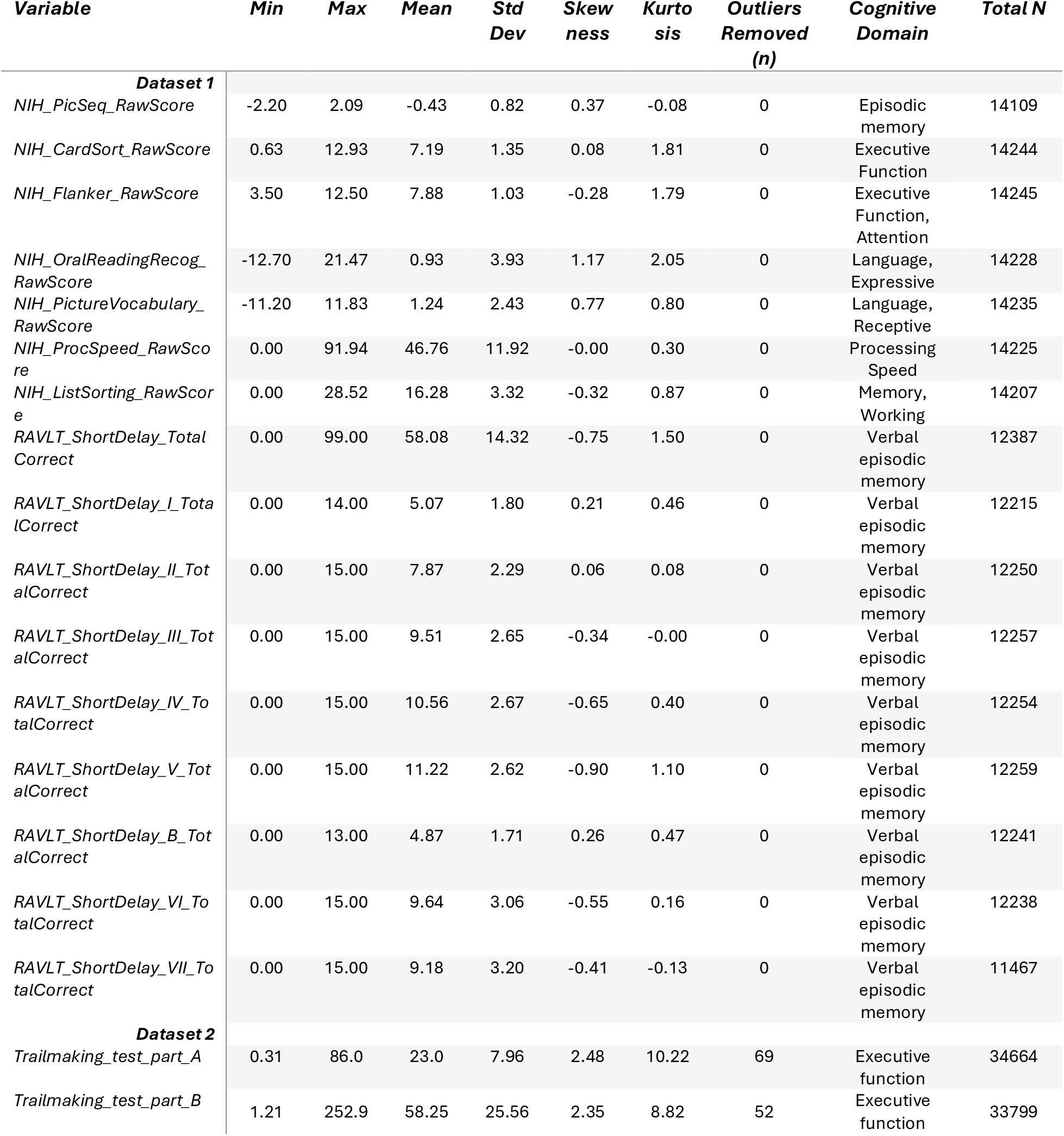
showing the summary statistics for the cognitive tests of dataset 1, combining all the NIH and RAVLT measures, and dataset 2 from the UK Biobank and HCP aging. For each variable, we present the minimum, maximum, mean, and standard deviation (SD), along with the skewness and kurtosis, which describe the shape of the distribution, the total number of outliers removed (n, |z| > 7), and the total number of observations (N). For more information on the different assessments, see: https://nihtoolbox.org/.

| ***Variable*** | ***Min*** | ***Max*** | ***Mean*** | ***Std Dev*** | ***Skewness*** | ***Kurtosis*** | ***Outliers Removed (n)*** | ***Cognitive Domain*** | ***Total N*** |
|---|---|---|---|---|---|---|---|---|---|
| ***Dataset 1*** |  |  |  |  |  |  |  |  |  |
| *NIH_PicSeq_RawScore* | -2.20 | 2.09 | -0.43 | 0.82 | 0.37 | -0.08 | 0 | Episodic memory | 14109 |
| *NIH_CardSort_RawScore* | 0.63 | 12.93 | 7.19 | 1.35 | 0.08 | 1.81 | 0 | Executive Function | 14244 |
| *NIH_Flanker_RawScore* | 3.50 | 12.50 | 7.88 | 1.03 | -0.28 | 1.79 | 0 | Executive Function, Attention | 14245 |
| *NIH_OralReadingRecog_RawScore* | -12.70 | 21.47 | 0.93 | 3.93 | 1.17 | 2.05 | 0 | Language, Expressive | 14228 |
| *NIH_PictureVocabulary_RawScore* | -11.20 | 11.83 | 1.24 | 2.43 | 0.77 | 0.80 | 0 | Language, Receptive | 14235 |
| *NIH_ProcSpeed_RawScore* | 0.00 | 91.94 | 46.76 | 11.92 | -0.00 | 0.30 | 0 | Processing Speed | 14225 |
| *NIH_ListSorting_RawScore* | 0.00 | 28.52 | 16.28 | 3.32 | -0.32 | 0.87 | 0 | Memory, Working | 14207 |
| *RAVLT_ShortDelay_Total Correct* | 0.00 | 99.00 | 58.08 | 14.32 | -0.75 | 1.50 | 0 | Verbal episodic memory | 12387 |
| *RAVLT_ShortDelay_I_TotalCorrect* | 0.00 | 14.00 | 5.07 | 1.80 | 0.21 | 0.46 | 0 | Verbal episodic memory | 12215 |
| *RAVLT_ShortDelay_II_TotalCorrect* | 0.00 | 15.00 | 7.87 | 2.29 | 0.06 | 0.08 | 0 | Verbal episodic memory | 12250 |
| *RAVLT_ShortDelay_III_TotalCorrect* | 0.00 | 15.00 | 9.51 | 2.65 | -0.34 | -0.00 | 0 | Verbal episodic memory | 12257 |
| *RAVLT_ShortDelay_IV_TotalCorrect* | 0.00 | 15.00 | 10.56 | 2.67 | -0.65 | 0.40 | 0 | Verbal episodic memory | 12254 |
| *RAVLT_ShortDelay_V_TotalCorrect* | 0.00 | 15.00 | 11.22 | 2.62 | -0.90 | 1.10 | 0 | Verbal episodic memory | 12259 |
| *RAVLT_ShortDelay_B_TotalCorrect* | 0.00 | 13.00 | 4.87 | 1.71 | 0.26 | 0.47 | 0 | Verbal episodic memory | 12241 |
| *RAVLT_ShortDelay_VI_TotalCorrect* | 0.00 | 15.00 | 9.64 | 3.06 | -0.55 | 0.16 | 0 | Verbal episodic memory | 12238 |
| *RAVLT_ShortDelay_VII_TotalCorrect* | 0.00 | 15.00 | 9.18 | 3.20 | -0.41 | -0.13 | 0 | Verbal episodic memory | 11467 |
| ***Dataset 2*** |  |  |  |  |  |  |  |  |  |
| *Trailmaking_test_part_A* | 0.31 | 86.0 | 23.0 | 7.96 | 2.48 | 10.22 | 69 | Executive function | 34664 |
| *Trailmaking_test_part_B* | 1.21 | 252.9 | 58.25 | 25.56 | 2.35 | 8.82 | 52 | Executive function | 33799 |

To fit the cognitive reference charts, we only included a cognitive test in the analysis if it was measured in more than two cohorts (see Table 3). For the UK Biobank, this criterion excluded most tests, as they were designed specifically for the UK Biobank and showed little direct overlap with other tests, except for the Trailmaking_test_part_A and Trailmaking_test_part_B. In general, it is possible to include slight differences in tests between cohorts, as we will demonstrate in the federated learning section. However, when fundamental differences in test design exist, it is not recommended to combine these tests^60^. In the discussion, we expand on the nuances of combining different cognitive tests and cohorts, as often they intend to measure the same underlying cognitive construct, but small changes in administration, language, and test details can impact cognitive scores significantly. We also demonstrate the added benefit of HBR, which allows for the incorporation of these test differences through an upweighting or downweighting of the prior information (see ‘Federated Cognitive Reference Charts’ section in the results).

### Normative model specifications

The normative modelling approaches tested in this paper were primarily based on the HBR framework implemented in the PCNtoolkit (version 1.1.2^32^). Across all the models, we estimated the cognitive variables as a nonlinear function of age using a B-spline basis function with five equally spaced knots and degree three. Sex and site were included as batch effects and implemented as random intercept offsets for the mean of the distribution. Input covariates were scaled prior to the model fit, using the standard scaler in the PCNtoolkit, 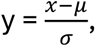 with *μ* the mean and *σ* the standard deviation estimated from the data. For each cognitive measure, a separate model was fit, in which a train–test split was performed with 80% of the data assigned to training and 20% to testing. Importantly, batch effects (site and sex) were conserved across the split, such that no new site appeared exclusively in either train or test. The patient data of the HCP psychosis sample were placed in a separate patient test sample; thus, only the healthy controls were used for model estimation.

For each cognitive variable, a normative HBR curve was fit independently using four chains with 1,500 posterior samples and 500 tuning samples per chain. The sampling was performed using the standard implemented PyMC library with the No-U-Turn Sampler (NUTS)^21,61^.

Different settings and model designs of the normative methods available in the PCNtoolkit were tested, including a Bayesian Linear Regression (BLR) and Hierarchical Bayesian regression (HBR). We performed a systematic model comparison across all model settings described in the Supplement, and the detailed results of these analyses can be found in the Supplementary Figures 19-30. In general, it was found that a simple HBR model with a fixed variance and a fixed kurtosis for the NIH and RAVLT tests (dataset 1) and an SHASHb likelihood HBR model for the Trail Making tests (dataset 2) were preferred. As shown in the Supplementary results, a higher flexibility for some variables meant that the models did not converge well, and thus a less flexible model was often preferred.

For every final fitted model, we generated centile plots and Q-Q plots (split by site) to assess the model assumptions. Model-specific evaluation metrics (Explained variance (R²), RMSE, MAE, as well as distribution shape-sensitive metrics such as Q-Q plots, skew, and kurtosis) were extracted for both train and test sets and are reported in the results section. Finally, the deviation scores of the cognitive reference charts were compared with the standardised and raw scores of each dataset, highlighting potentially misestimated site effects and non-normality in the standardised dataset. For further details and recommendations on how to apply and test the normative models, we refer readers to two protocol papers^32,62^.

### Local norms through federated learning

We demonstrate the applicability of constructing local and transferred cognitive reference charts through the federated learning function within the HBR framework. As noted, the PCNtoolkit package, that we use here, allows global cognitive reference charts to be adapted to local new sites, where the sample sizes are limited, or data sharing is not possible (see Figure 7). When applying the large reference models to new (clinical) datasets, two common challenges arise: site effects (e.g. cultural, language, and demographic differences) and differences in test administration and versions that can lead to downstream variation in the estimated cognitive variable. To solve these problems without training a new local model from scratch, two functionalities are present within federated learning using the PCNtoolkit: transfer and extend.

Both the transfer and extend approaches allow for the combination of the existing cognitive reference chart with a smaller local dataset from a different cohort^32^. The extension method updates the parameters of the original reference model through re-estimating the model, effectively building an expanded reference model. In contrast, the transfer method derives a local model from the larger reference model without refitting the parameters for the existing sites, but using them as informative priors. This provides a model that is well calibrated to the target dataset (see Figure 7).

One key parameter controlling the influence of the prior information (in this case, the larger cognitive reference chart) is the freedom parameter. We set this parameter to 1 by default, corresponding to using the original posterior as the full prior. However, when the distributions or cohorts are expected to differ a lot due to site or other effects, the freedom parameter can be adjusted: freedom < 1 enforces a tighter prior, and freedom > 1 allows greater deviation from the reference model. The transfer approach can be useful when we want to create local norms that can differ in various ways from the global reference charts.

Here, we demonstrate how the transfer function can be used to create local normative models that learn, via informative priors, from general cognitive reference charts. For this analysis, we first estimate a cognitive reference chart for the Trail Making Tests Part A and B using data from the UK Biobank and subsequently transfer this model using one held-out dataset: HCP Aging. Prior to constructing the model, both the reference data and the held-out transfer data were split into training and test sets with an 80-20 split. As before, an HBR model was fit with a B-spline basis using the training set of the UK Biobank data. In this case, as the data are more skewed, a SHASHb likelihood was preferred, with age as a covariate and site and sex as batch effects. The sampling strategy was equal to that described earlier. After obtaining a fully trained normative model as the reference model, model fit-statistics, centile curves, and Q-Q plots were derived and visualised for every stage (that is testing set of the UK Biobank to assess the quality of the reference model, and the training and testing set of the HCP Aging) to characterise the distribution as well as the model fit.

## Data Availability

All data produced are available online. The ABCD cohort is available through the NIH Brain Development Cohorts (NBDC) Data Sharing Platform (https://www.nbdc-datahub.org/). The UK Biobank can be accessed following an application at https://www.ukbiobank.ac.uk/. Fully de-identified data of the Human Connectome Project data are openly available in the ConnectomeDB website (https://www.humanconnectome.org/software/connectomedb).

https://www.ukbiobank.ac.uk/

https://www.humanconnectome.org/software/connectomedb

## Acknowledgements

For the ABCD sample: Data used in the preparation of this article were obtained from the Adolescent Brain Cognitive Development™ (ABCD) Study, held in the NIH Brain Development Cohorts Data Sharing Platform. This is a multisite, longitudinal study designed to recruit more than 10,000 children aged 9–10 and follow them over 10 years into early adulthood. The ABCD Study® is supported by the National Institutes of Health and additional federal partners under award numbers: U01DA041048, U01DA050989, U01DA051016, U01DA041022, U01DA051018, U01DA051037, U01DA050987, U01DA041174, U01DA041106, U01DA041117, U01DA041028, U01DA041134, U01DA050988, U01DA051039, U01DA041156, U01DA041025, U01DA041120, U01DA051038, U01DA041148, U01DA041093, U01DA041089, U24DA041123, U24DA041147. A full list of supporters is available at Federal Partners – ABCD Study. ABCD Consortium investigators designed and implemented the study and/or provided data but did not necessarily participate in the analysis or writing of this report. This manuscript reflects the views of the authors and may not reflect the opinions or views of the NIH or ABCD Consortium investigators.

For the UK Biobank sample: This research has been conducted using the UK Biobank Resource under Application Number 23668 (PI: Andre Marquand).

For the HCP1200 dataset: Data were provided in part by the Human Connectome Project, WU-Minn Consortium (Principal Investigators: David Van Essen and Kamil Ugurbil; 1U54MH091657) funded by the 16 NIH Institutes and Centers that support the NIH Blueprint for Neuroscience Research; and by the McDonnell Center for Systems Neuroscience at Washington University.

For the HCP-aging dataset: Research reported in this publication was supported by the National Institute On Aging of the National Institutes of Health under Award Number U01AG052564 and by funds provided by the McDonnell Center for Systems Neuroscience at Washington University in St. Louis.

For the HCP-dev dataset: Research reported in this publication was supported by the National Institute Of Mental Health of the National Institutes of Health under Award Number U01MH109589 and by funds provided by the McDonnell Center for Systems Neuroscience at Washington University in St. Louis. The HCP-Development 2.0 Release data used in this report came from DOI: 10.15154/1520708.

For the HCP-psychosis dataset: Research using Human Connectome Project for Early Psychosis (HCP-EP) data reported in this publication was supported by the National Institute of Mental Health of the National Institutes of Health under Award Number U01MH109977. The HCP-EP 1.1 Release data used in this report came from DOI: 10.15154/1522899.

## Data Availability declaration

The ABCD cohort is available through the NIH Brain Development Cohorts (NBDC) Data Sharing Platform (https://www.nbdc-datahub.org/). The UK Biobank can be accessed following an application at https://www.ukbiobank.ac.uk/. Fully de-identified data of the Human Connectome Project data are openly available in the ConnectomeDB website (https://www.humanconnectome.org/software/connectomedb).

## Author Contribution declaration

Conceptualization: CF, BRB, CBF, LSS, GC, DS, VG, OAA, LTW, TU, PD, AFM; Data curation: BRB; Formal analysis: CF; Funding acquisition: OAA, LTW, TU, PD, AFM; Investigation: CF, BRB; Methodology: CF, BRB, OAA, LTW, TU, PD, AFM; Project administration: TU, PD, AFM; Software: SB, AFM; Supervision: VG, OAA, LTW, TU, PD, AFM; Validation: CF, BRB; Visualization: CF, LCM; Writing - original draft: CF, BRB, LCM, SB, CBF, LSS, VB, GC, DS, VG, OAA, LTW, TU, PD, AFM; Writing - review & editing: CF, BRB, LCM, LSS, VB, GC, DS, VG, OAA, LTW, TU, PD, AFM.

