## Supplementary Information for "Cognitive Reference Charts for Precision Psychiatry"

### Supplement

### Simulation Supplementary Results

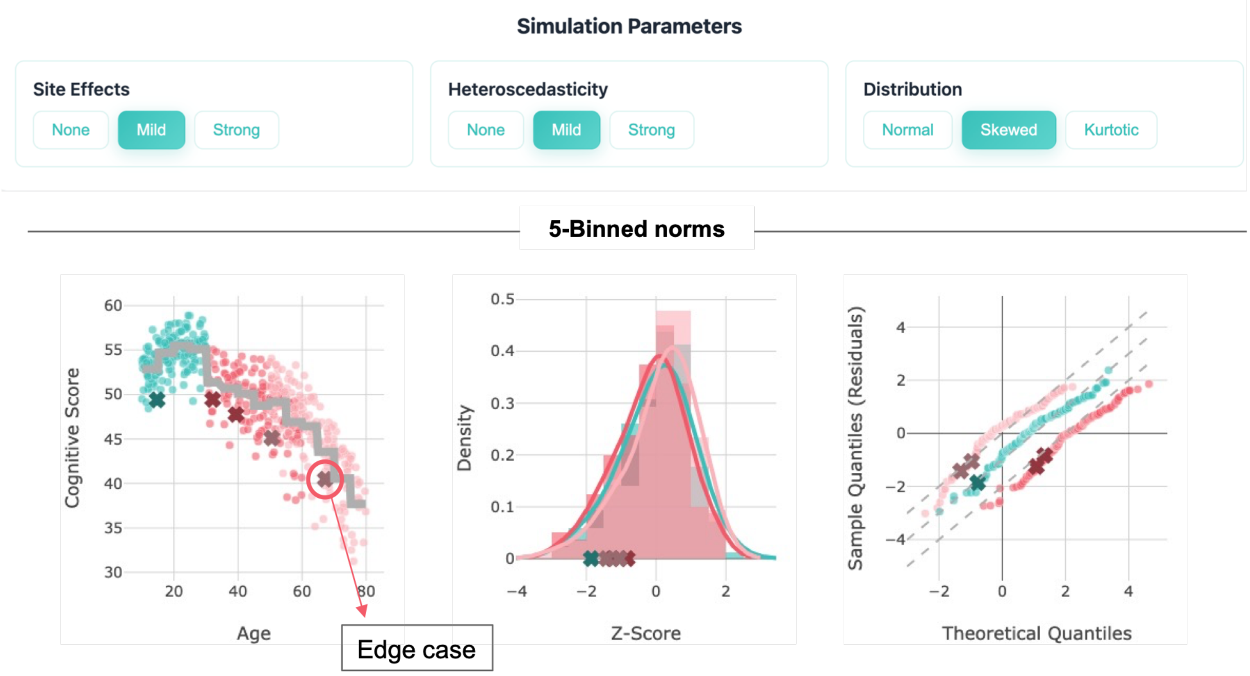


*Figure 1 - Highlighting an edge case observed for specific subjects when applying the binning method. The left panel shows the scatter plot, the middle panel displays the histogram of deviation scores, and the right panel presents the QQ-plot.*

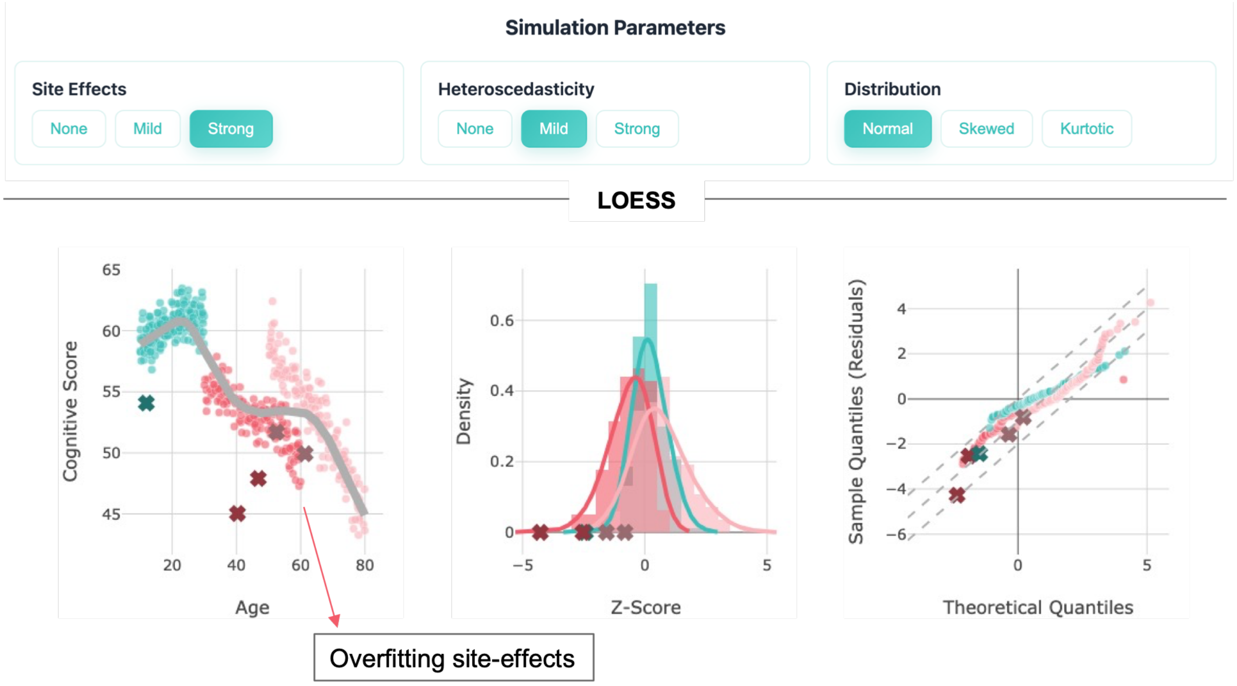


*Figure 2 - Highlighting an edge case observed for specific subjects when applying the LOESS method with strong site effects. The left panel shows the scatter plot, the middle panel displays the histogram of deviation scores, and the right panel presents the QQ-plot.*

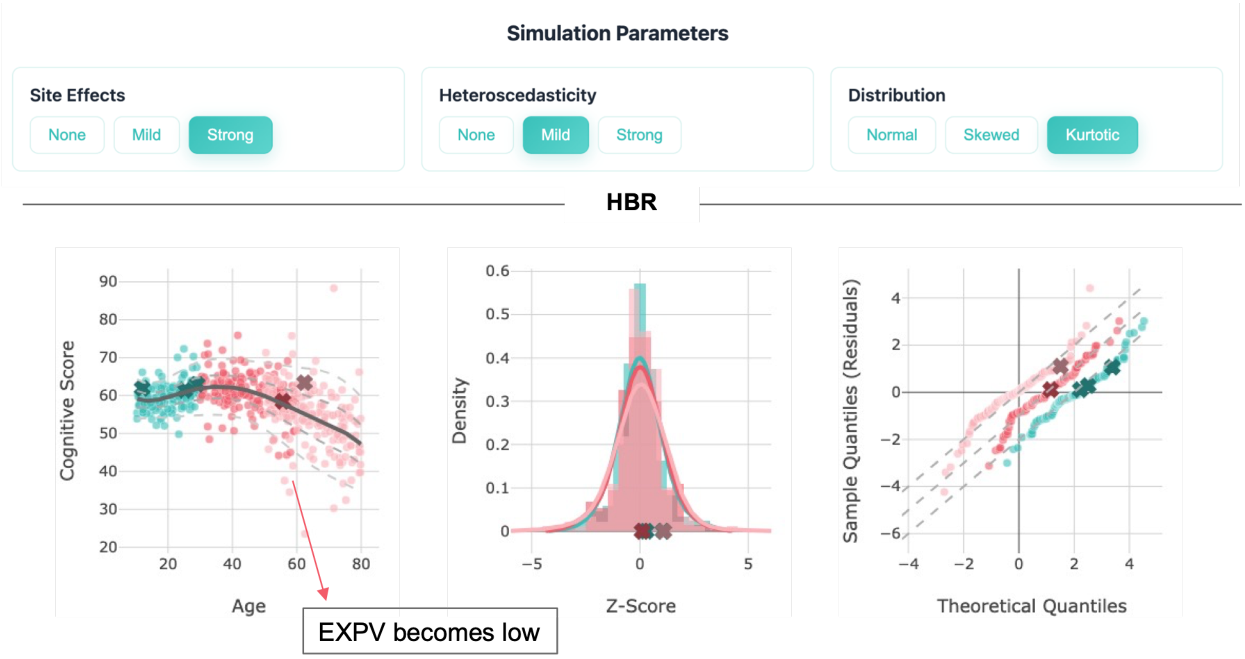


*Figure 3 - Highlighting an edge case observed for when mild heteroscedasticity and a kurtotic distribution are present, lowering the estimated EXPV. The left panel shows the scatter plot, the middle panel displays the histogram of deviation scores, and the right panel presents the QQ-plot.*

| | Site effects | heteroscedasticity | distribution | MACE | MSLL | $R^2$ | RMSE | ShapiroW |
|---|---|---|---|---|---|---|---|---|
| 1 | none | none | normal | 0.02 | 1.47 | 0.95 | 0.23 | 0.99 |
| 2 | none | none | skewed | 0.01 | 1.14 | 0.9 | 0.32 | 0.99 |
| 3 | none | none | kurtotic | 0.02 | 0.6 | 0.7 | 0.55 | 0.98 |
| 4 | none | mild | normal | 0.01 | 1.26 | 0.92 | 0.28 | 0.98 |
| 5 | none | mild | skewed | 0.03 | 0.87 | 0.8 | 0.44 | 0.97 |
| 6 | none | mild | kurtotic | 0.03 | 0.35 | 0.41 | 0.77 | 0.97 |
| 7 | none | strong | normal | 0.01 | 1.12 | 0.89 | 0.34 | 0.99 |
| 8 | none | strong | skewed | 0.03 | 0.86 | 0.8 | 0.45 | 0.97 |
| 9 | none | strong | kurtotic | 0.02 | 0.27 | 0.24 | 0.87 | 0.98 |
| 10 | mild | none | normal | 0.02 | 1.29 | 0.93 | 0.27 | 0.97 |
| 11 | mild | none | skewed | 0.02 | 1.13 | 0.9 | 0.32 | 0.98 |
| 12 | mild | none | kurtotic | 0.03 | 0.65 | 0.73 | 0.52 | 0.97 |
| 13 | mild | mild | normal | 0.01 | 1.19 | 0.91 | 0.31 | 0.98 |
| 14 | mild | mild | skewed | 0.03 | 0.79 | 0.78 | 0.47 | 0.97 |
| 15 | mild | mild | kurtotic | 0.02 | 0.38 | 0.45 | 0.74 | 0.97 |
| 16 | mild | strong | normal | 0.02 | 0.91 | 0.84 | 0.4 | 0.99 |
| 17 | mild | strong | skewed | 0.03 | 0.63 | 0.66 | 0.58 | 0.97 |
| 18 | mild | strong | kurtotic | 0.03 | 0.28 | 0.21 | 0.89 | 0.98 |
| 19 | strong | none | normal | 0.02 | 1.14 | 0.89 | 0.33 | 0.96 |
| 20 | strong | none | skewed | 0.01 | 1.19 | 0.91 | 0.3 | 0.99 |

| | | | | | | | | |
|---|---|---|---|---|---|---|---|---|
| **21** | strong | none | kurtotic | 0.02 | 0.45 | 0.6 | 0.63 | 0.98 |
| **22** | strong | mild | normal | 0.02 | 1.24 | 0.92 | 0.29 | 0.97 |
| **23** | strong | mild | skewed | 0.03 | 0.77 | 0.77 | 0.48 | 0.97 |
| **24** | strong | mild | kurtotic | 0.03 | 0.42 | 0.45 | 0.74 | 0.97 |
| **25** | strong | strong | normal | 0.01 | 0.77 | 0.77 | 0.48 | 0.99 |
| **26** | strong | strong | skewed | 0.03 | 0.8 | 0.76 | 0.49 | 0.96 |
| **27** | strong | strong | kurtotic | 0.04 | 0.3 | 0.23 | 0.88 | 0.94 |

*Table 1: Model evaluation statistics for the different simulation scenarios; the models were estimated using the HBR model with a normal likelihood. Statistics displayed are: MACE: calibration of predicted uncertainty, MSLL: model fit and uncertainty, $R^2$: variance explained (accuracy), RMSE: Root mean square error, and the Shapiro-W: distributional check (residual normality).*

### Data Description Dataset 1

For the first large cognitve dataset, as described in the main text, we added 14,344 individuals from four datasets spanning the human lifespan (5.6–100 years), including data from the following sources: HCP1200[1,2] (N = 1,206, aged 22–37 years, 54% female), HCP Aging[3] (N = 720, aged 36–100 years, 56% female), HCP Development[4] (N = 468, aged 5.6–22years, 54% female), HCP Early Psychosis[5] (N = 234, aged 16-35 years, 38% female) and ABCD[6] (N = 11,716, aged 8–11 years, 48% female) using data from the NIH toolbox[7,8] and the RAVLT[9]. Here we will give some details on the datasets and used tests, but for further information, also see the original publications and release manuals:
https://www.humanconnectome.org/storage/app/media/documentation/s1200/HCP_S1200_Release_Reference_Manual.pdf

https://www.humanconnectome.org/storage/app/media/documentation/HCP-EP1.1/HCP-EP_Release_1.1_Manual.pdf

https://wiki.humanconnectome.org/

As well as the NIH toolbox information[7,8]. The NIH toolbox configuration measured consisted of seven tasks that together also generate three composite scores. In our sample, we used the following measures: the Dimensional Change Card Sort Test (NIH_CardSort), Flanker Inhibitory Control and Attention Test (NIH_Flanker), List Sorting Working Memory Test (NIH_ListSorting), Oral Reading Recognition Test (NIH_OralReading), Pattern Comparison Processing Speed Test (NIH_ProcSpeed), Picture Sequence Memory Test (NIH_PicSeq), Picture Vocabulary Test (NIH_PictureVocabulary), and the Rey Auditory Verbal Learning Test (RAVLT), for details see https://nihtoolbox.org/domain/cognition/ as well as the technical manual: https://nihbabytoolbox.org/wp-content/uploads/2024/04/NIHTB-V3-Technical-Manual_040524.pdf.

There are several “families” of HCP studies[10], and most follow the behavioural protocol laid out by the original HCP1200 sample[1] applying the NIH Toolbox for cognitive testing using version 1 or 2, depending on the year the data were collected. In general, the NIH Toolbox is a standardized test, although some updates were made to the cognitive tests to accommodate different participant ages and abilities[11]. As the test is (mostly) standardized, it allows for the comparison between different samples and overall harmonization, although some site effects can occur depending on how and in which language the test was administered. Every version of the NIH Toolbox also includes slight changes and should be assessed before deciding to harmonize different cognitive samples. One notable feature between v1 and v2 is that the administration of the test differed between sites, with ABCD and the later HCP samples using an iPad to perform the cognitive tests and the HCP1200 using the desktop version. This could lead to slight discrepancies between results, especially for reaction-time tasks[12]; see table. This makes site adaptation necessary when harmonizing the different datasets.

| COHORT / STUDY | NIH TOOLBOX VERSION USED | POPULATION | KEY REFERENCES |
|---|---|---|---|
| **HUMAN CONNECTOME PROJECT YOUNG ADULT (HCPS1200)** | NIH Toolbox Version 1[8] (desktop implementation | Healthy adults 22–37 | Barch et al., 2013[13] |
| **ADOLESCENT BRAIN COGNITIVE DEVELOPMENT STUDY (ABCD)** | NIH Toolbox Version 2 (iPad implementation) | Children 8-11 | Luciana et al., 2018[14] |
| **HUMAN CONNECTOME PROJECT IN DEVELOPMENT (HCP-DEVELOPMENT)** | NIH Toolbox Version 2.0 | Children & adolescents ages 5–22 | Somerville et al., 2018[4] |
| **HUMAN CONNECTOME PROJECT IN AGING (HCP-AGING)** | NIH Toolbox Version 2.0 | Adults ages 36–100 | Bookheimer et al., 2019[3] |
| **HUMAN CONNECTOME PROJECT EARLY PSYCHOSIS (HCP EARLY PSYCHOSIS)** | NIH Toolbox Version 2.1 | Healthy controls and participants with Schizophrenia, bipolar disorder or early psychosis | Jacobs et al., 2024[5]; |

*Table 2: Overview of the NIH toolbox versions used across dataset 1.*

#### Data Description Dataset 2

For the second dataset, we combined the UK Biobank and the HCP Aging datasets (N = 35,576) for the two Trail Making Tests (TMT across an age range of 36–100 years. The UK Biobank uses a web-based adaptation of the Trail Making Test (see also: https://biobank.ndph.ox.ac.uk/ukb/ukb/docs/trailmaking_doc.pdf). The concordance with the original tests has been discussed previously[15] and showed a correlation of 0.439 for the UKB TMT Part A and 0.663 for the UKB TMT Part B with the chosen reference test. The HCP Aging dataset uses the classic TMT A and B, as described here: https://www.humanconnectome.org/storage/app/media/documentation/LS2.0/LS_2.0_Release_Appendix_2.pdf. Although these tests are conceptually similar, they differ in administration format (computerized vs. paper-and-pencil), which may lead to slight differences in the cognitive processes being assessed. Thus, as presented in this paper, they can theoretically be harmonized, for example, through an extend or transfer function; however, in practice, it remains an open question whether the exact same cognitive features are being tested.

### Overview Figures NIH Cognitive Reference Charts

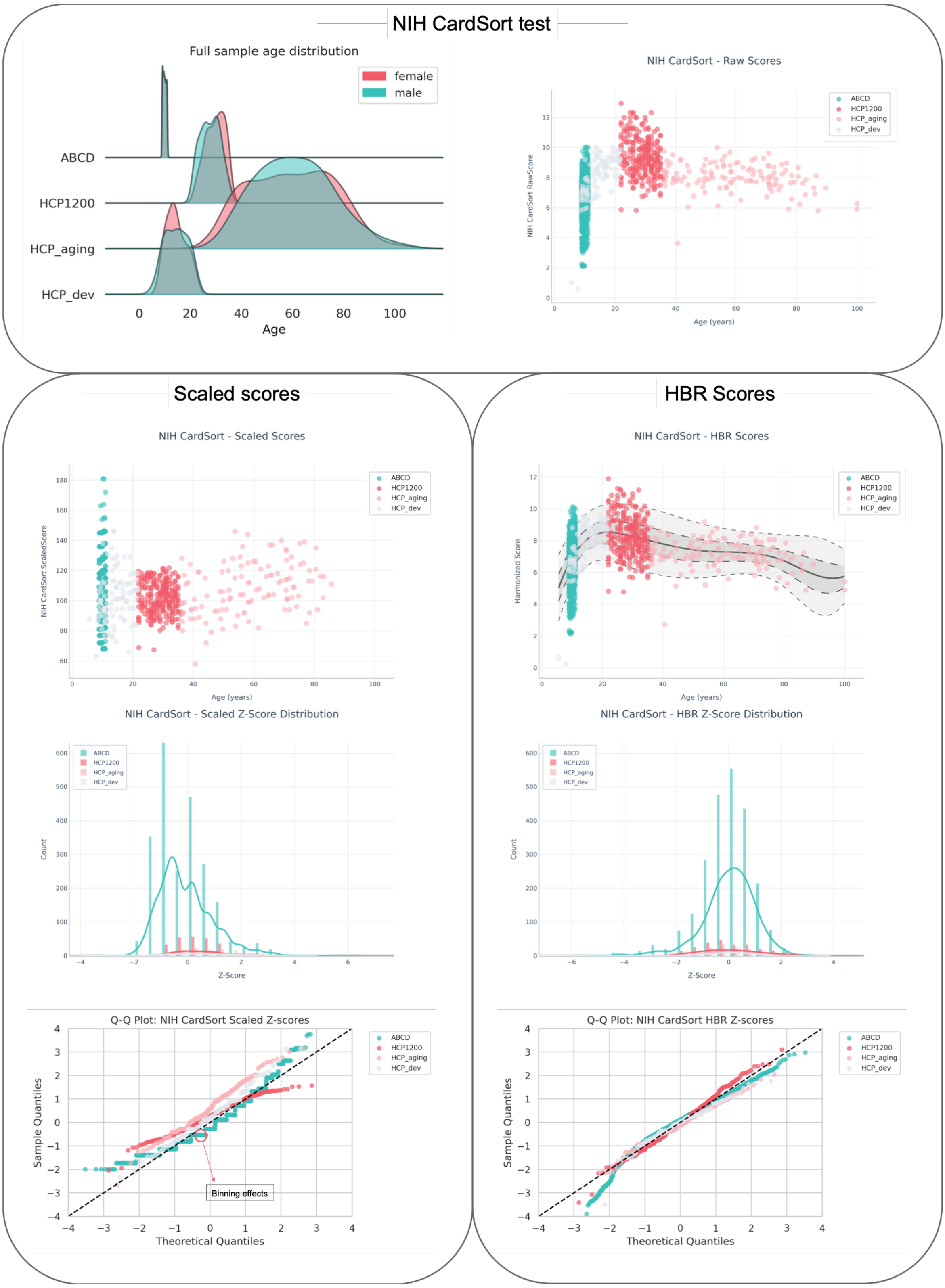


*Figure 4 Cognitive reference chart for the NIH Card Sort test. The top panel shows the sample age distribution and the scatter plot of the raw scores by age. The left panel presents the scaled scores, including a scatter plot, a distributional plot, and Q-Q plot. The right panel shows the HBR deviation scores, also with a scatter plot, a distributional plot, and Q-Q plot.*

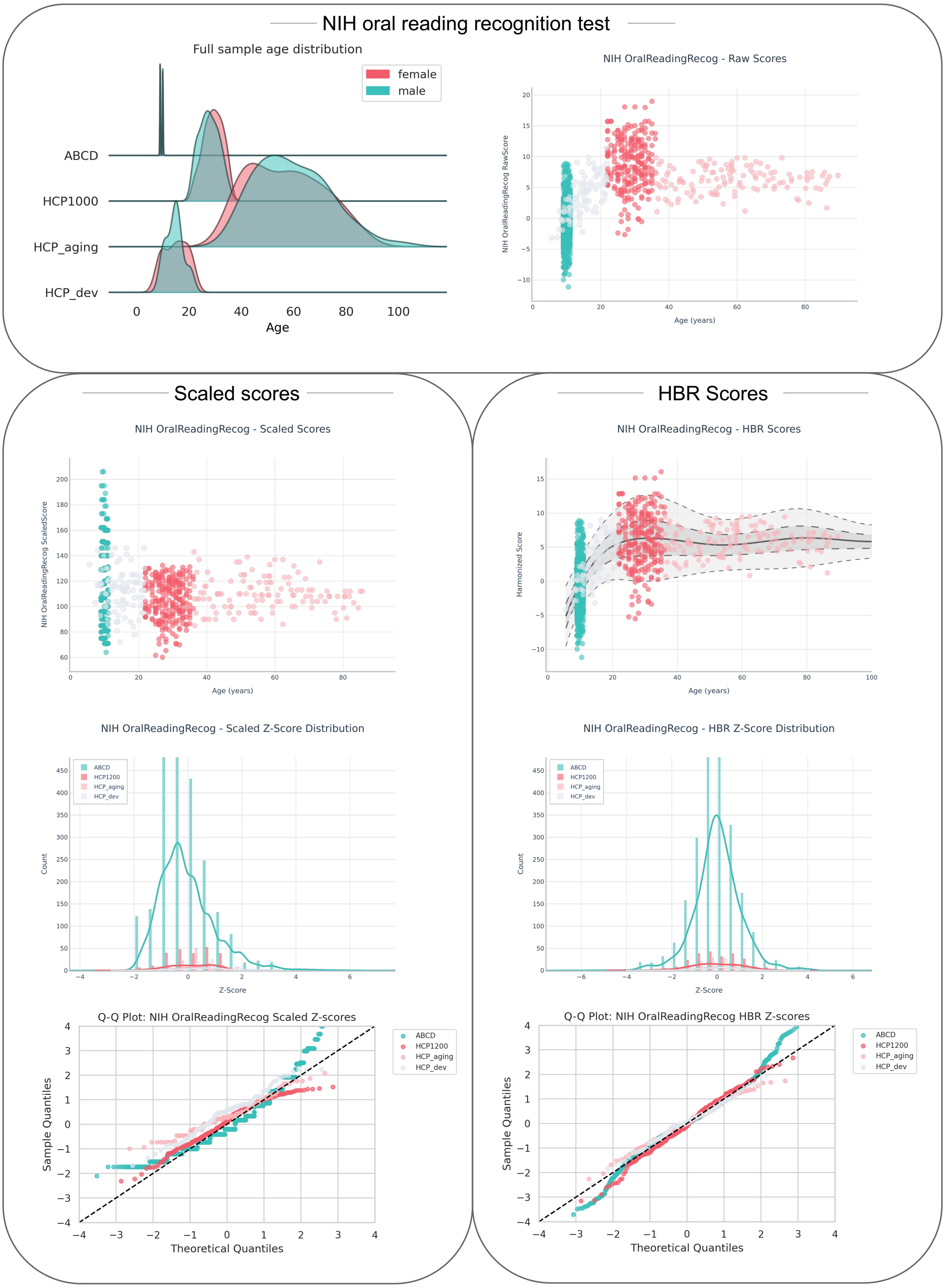


*Figure 5 Cognitive reference chart for the NIH oral reading recognition test. The top panel shows the sample age distribution and the scatter plot of the raw scores by age. The left panel presents the scaled scores, including a scatter plot, distributional plot, and Q-Q plot. The right panel shows the HBR deviation scores, also with a scatter plot, distributional plot, and Q-Q plot.*

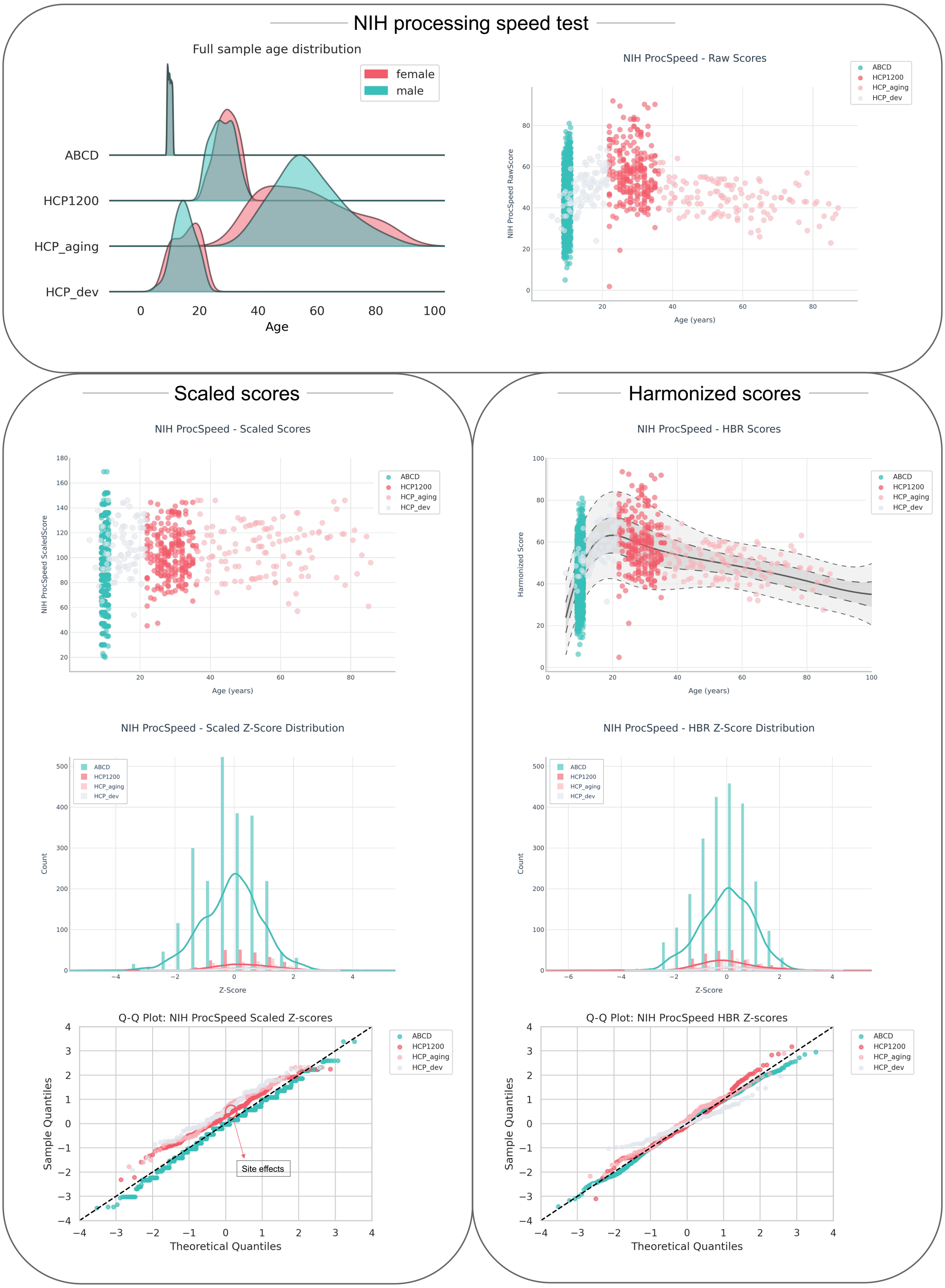


*Figure 6 Cognitive reference charts for the NIH processing speed test. The top panel shows the sample age distribution and the scatter plot of the raw scores by age. The left panel presents the scaled scores, including a scatter plot, distributional plot, and Q-Q plot. The right panel shows the HBR deviation scores, also with a scatter plot, a distributional plot, and Q-Q plot.*

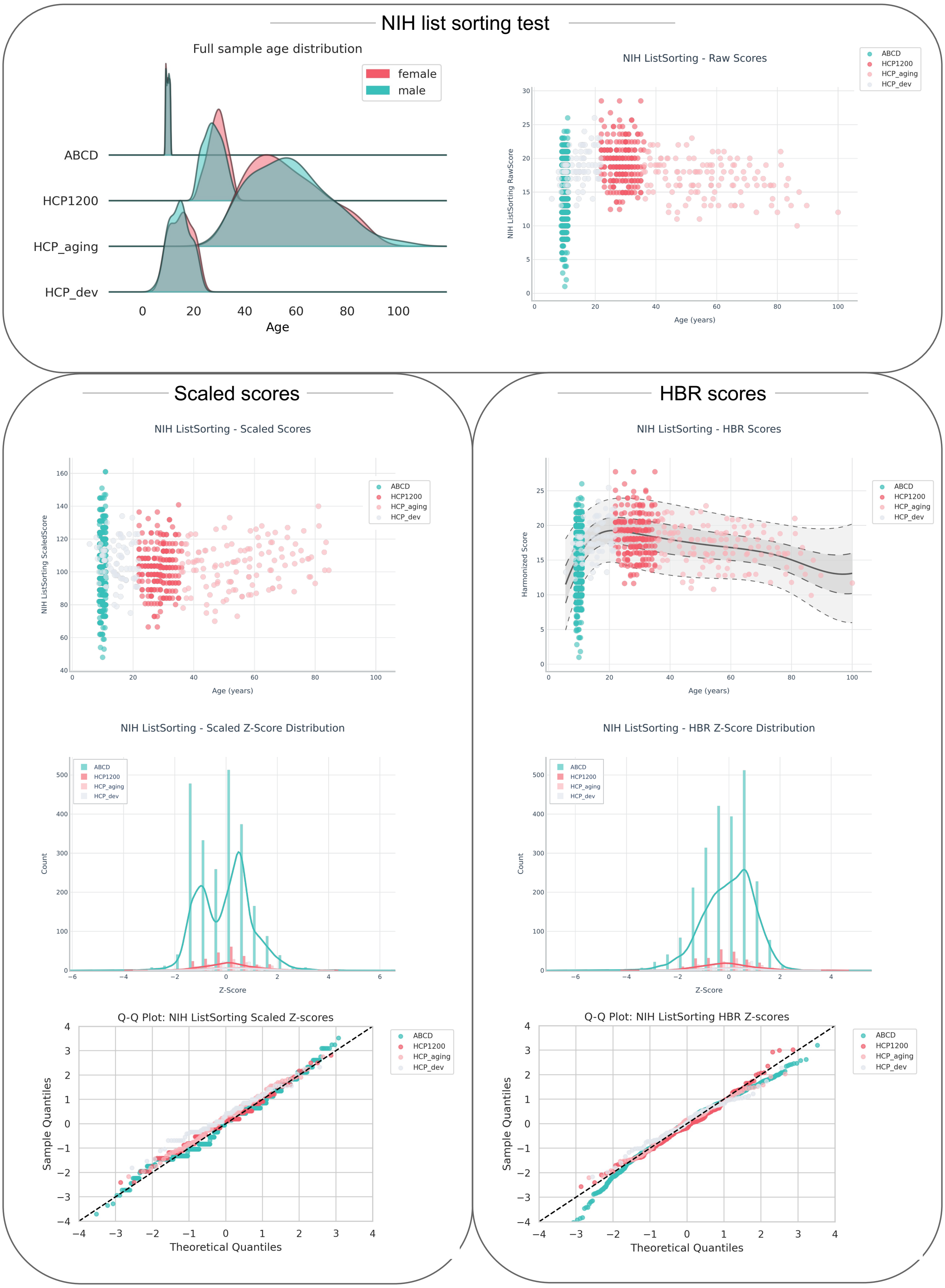


*Figure 7 Cognitive reference charts for the NIH list sorting test. The top panel shows the sample age distribution and the scatter plot of the raw scores by age. The left panel presents the scaled scores, including a scatter plot, distributional plot, and Q-Q plot. The right panel shows the HBR deviation scores, also with a scatter plot, distributional plot, and Q-Q plot.*

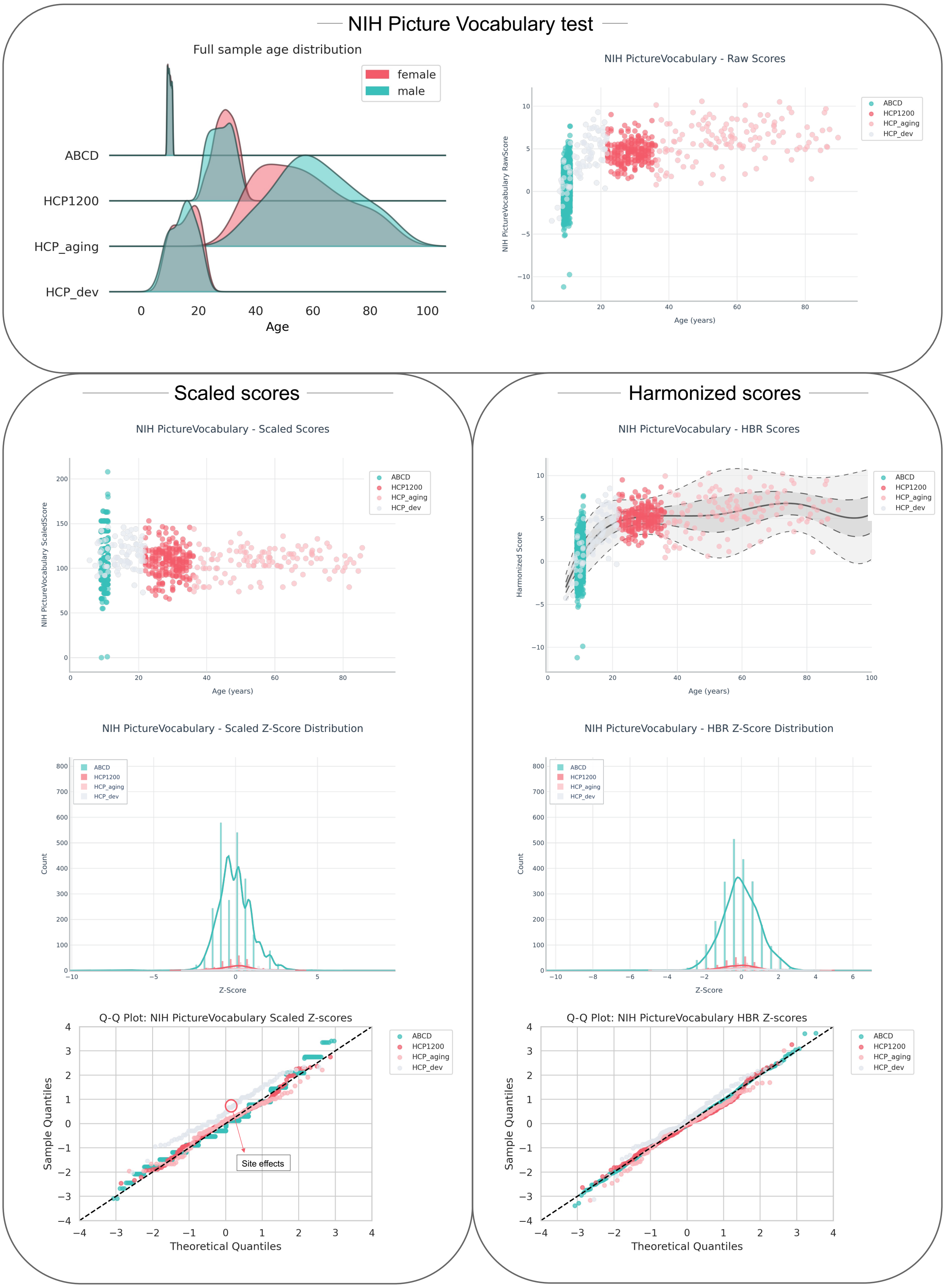


*Figure 8 Cognitive reference chart for the NIH Picture Vocabulary test. The top panel shows the sample age distribution and the scatter plot of the raw scores by age. The left panel presents the scaled scores, including a scatter plot, a distributional plot, and Q-Q plot. The right panel shows the HBR deviation scores, also with a scatter plot, a distributional plot, and Q-Q plot.*

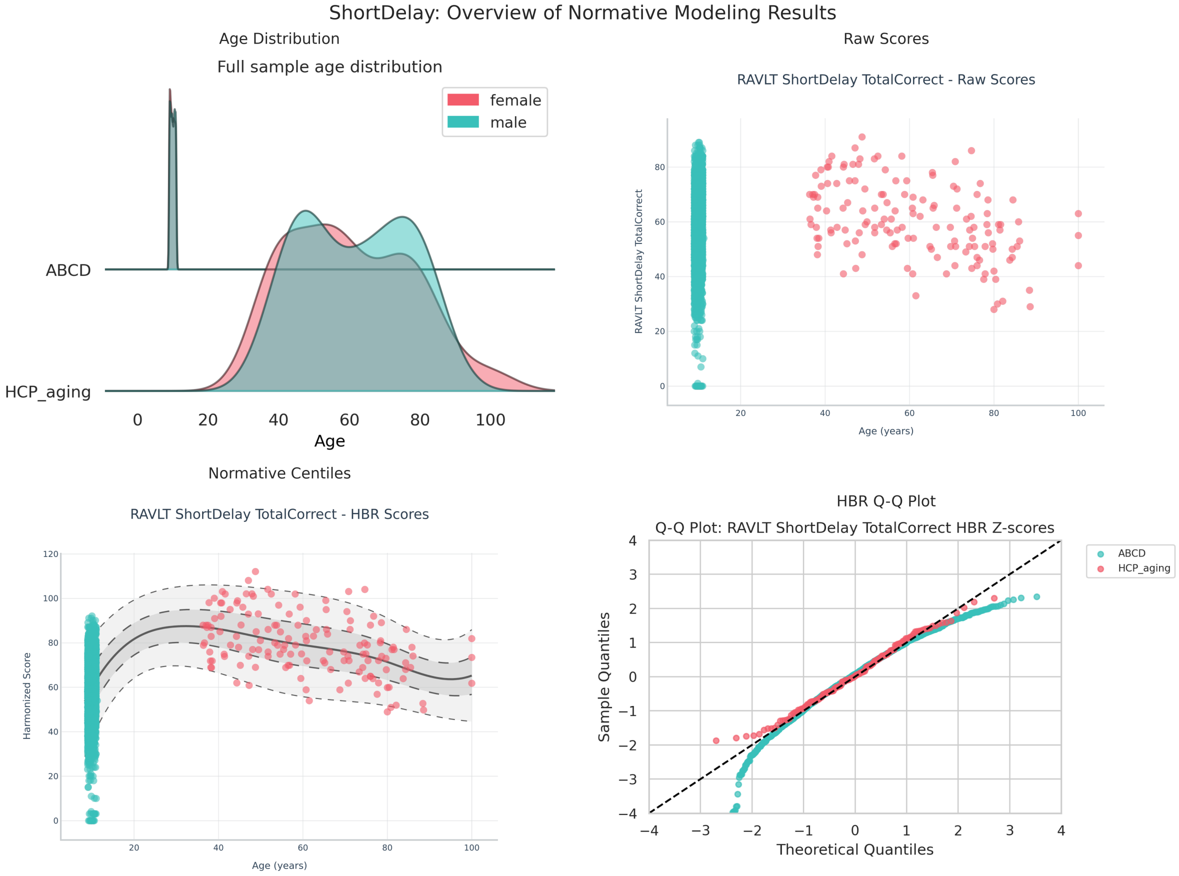


*Figure 9 Cognitive reference charts for the RAVLT Short Delay test total score of all the trials. The top panel shows the sample age distribution and the scatter plot of the raw scores by age. The bottom panel shows the HBR deviation scores, also with a scatter plot and the Q-Q plot.*

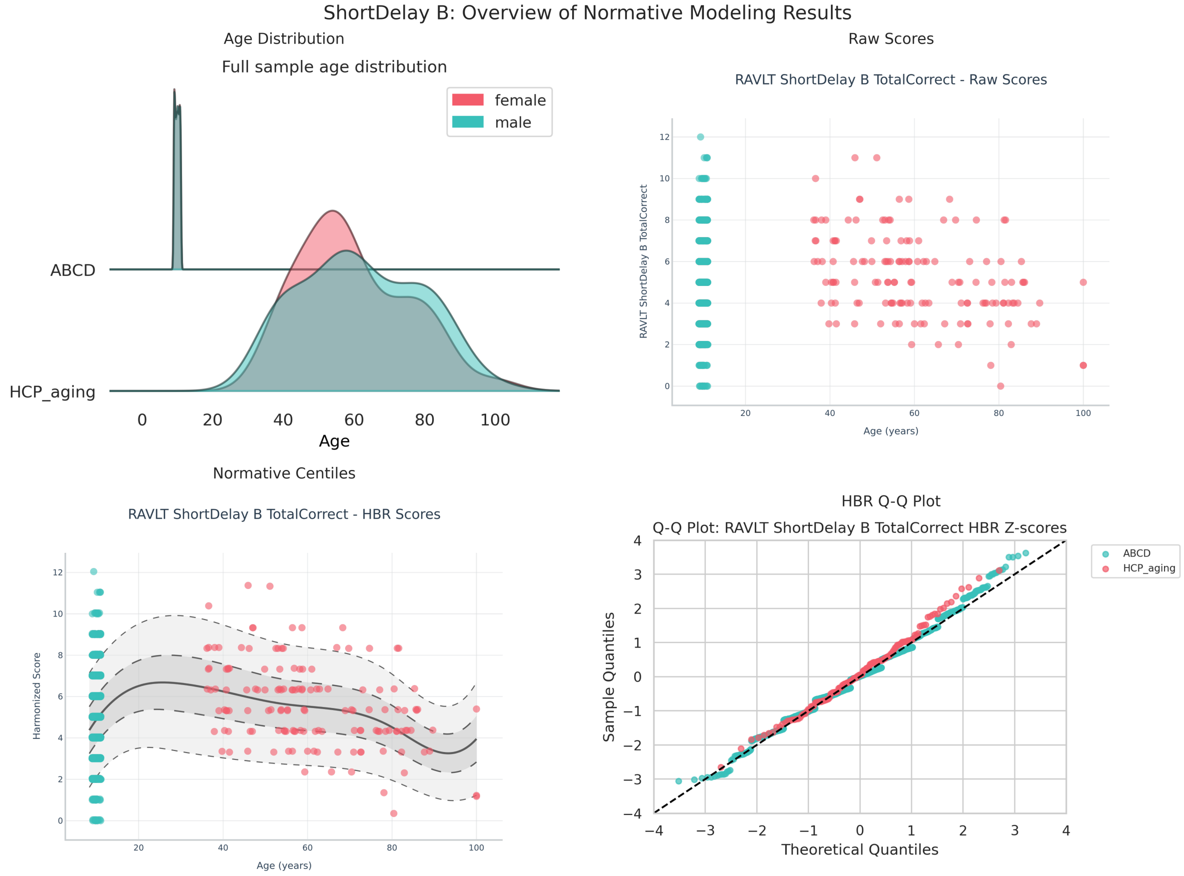


*Figure 10 Cognitive reference charts for the RAVLT Short Delay test, trial B. The top panel shows the sample age distribution and the scatter plot of the raw scores by age. The bottom panel shows the HBR deviation scores, also with a scatter plot and the Q-Q plot.*

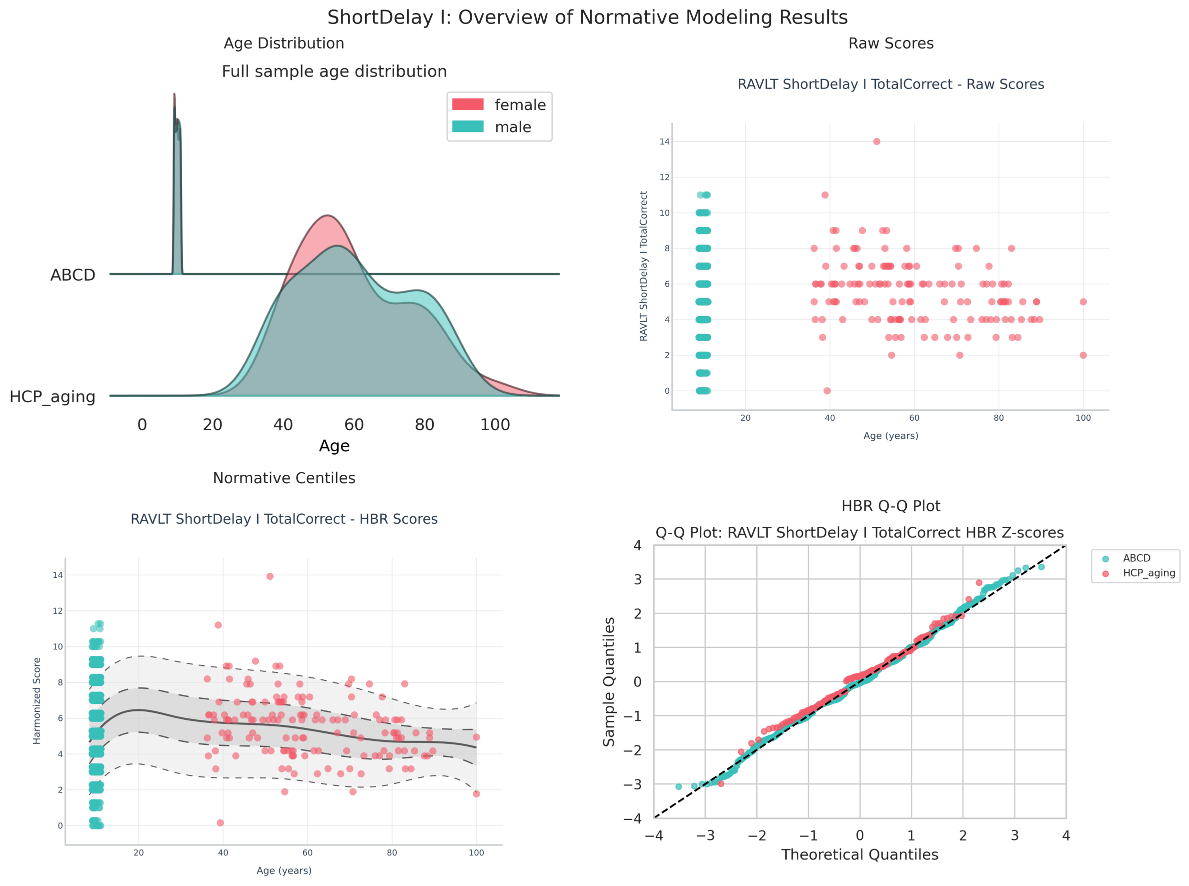


*Figure 11 Cognitive reference charts for the RAVLT Short Delay test trial I. The top panel shows the sample age distribution and the scatter plot of the raw scores by age. The bottom panel shows the HBR deviation scores, also with a scatter plot and the Q-Q plot.*

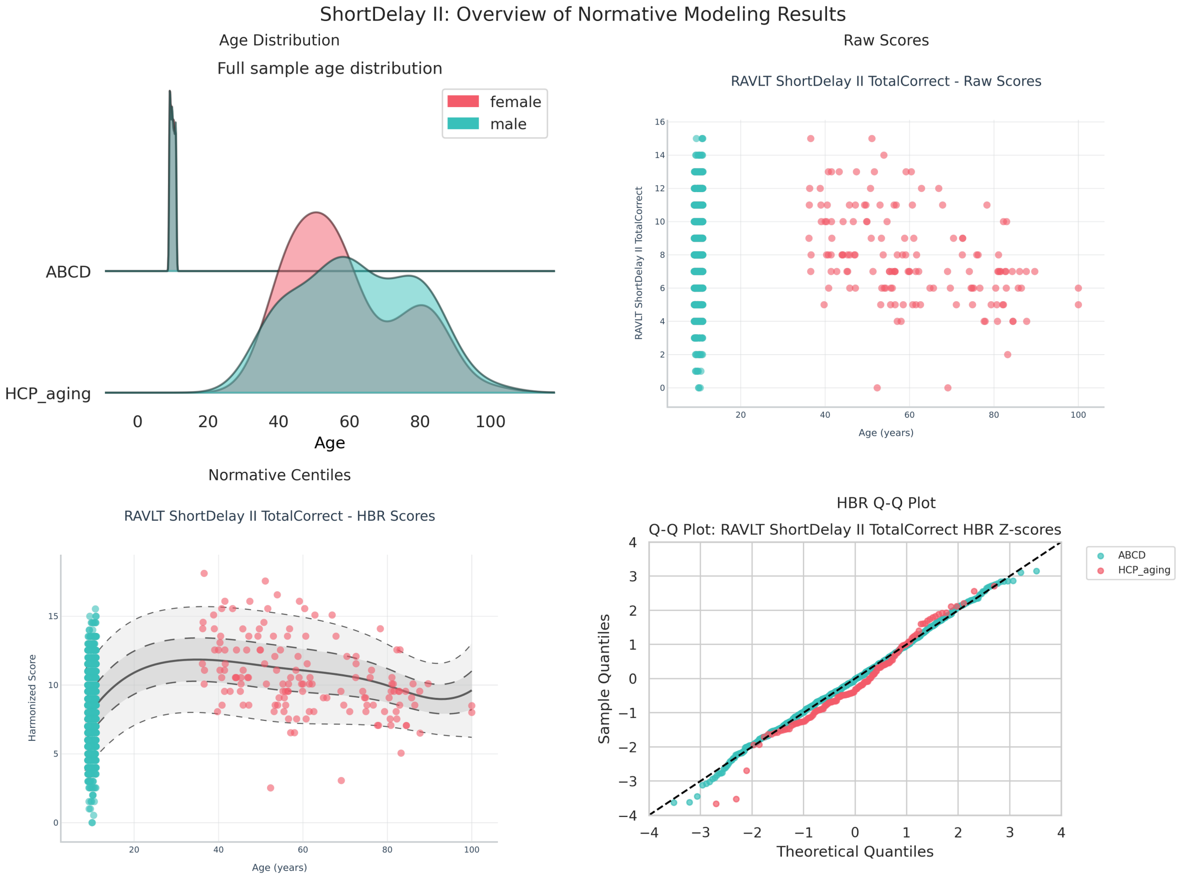


*Figure 12 Cognitive reference charts for the RAVLT Short Delay test trial II. The top panel shows the sample age distribution and the scatter plot of the raw scores by age. The bottom panel shows the HBR deviation scores, also with a scatter plot and the Q-Q plot.*

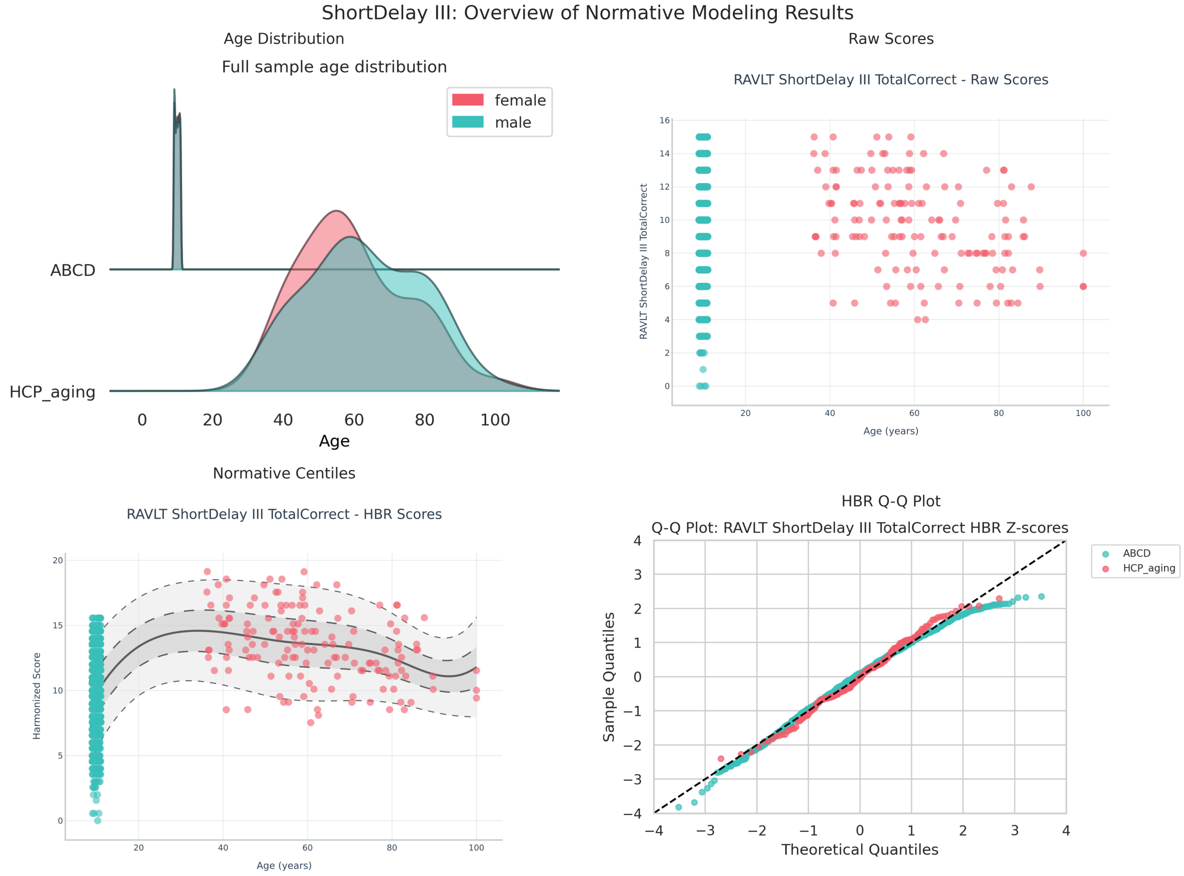


*Figure 13 Cognitive reference charts for the RAVLT Short Delay test trial III. The top panel shows the sample age distribution and the scatter plot of the raw scores by age. The bottom panel shows the HBR deviation scores, also with a scatter plot and the Q-Q plot.*

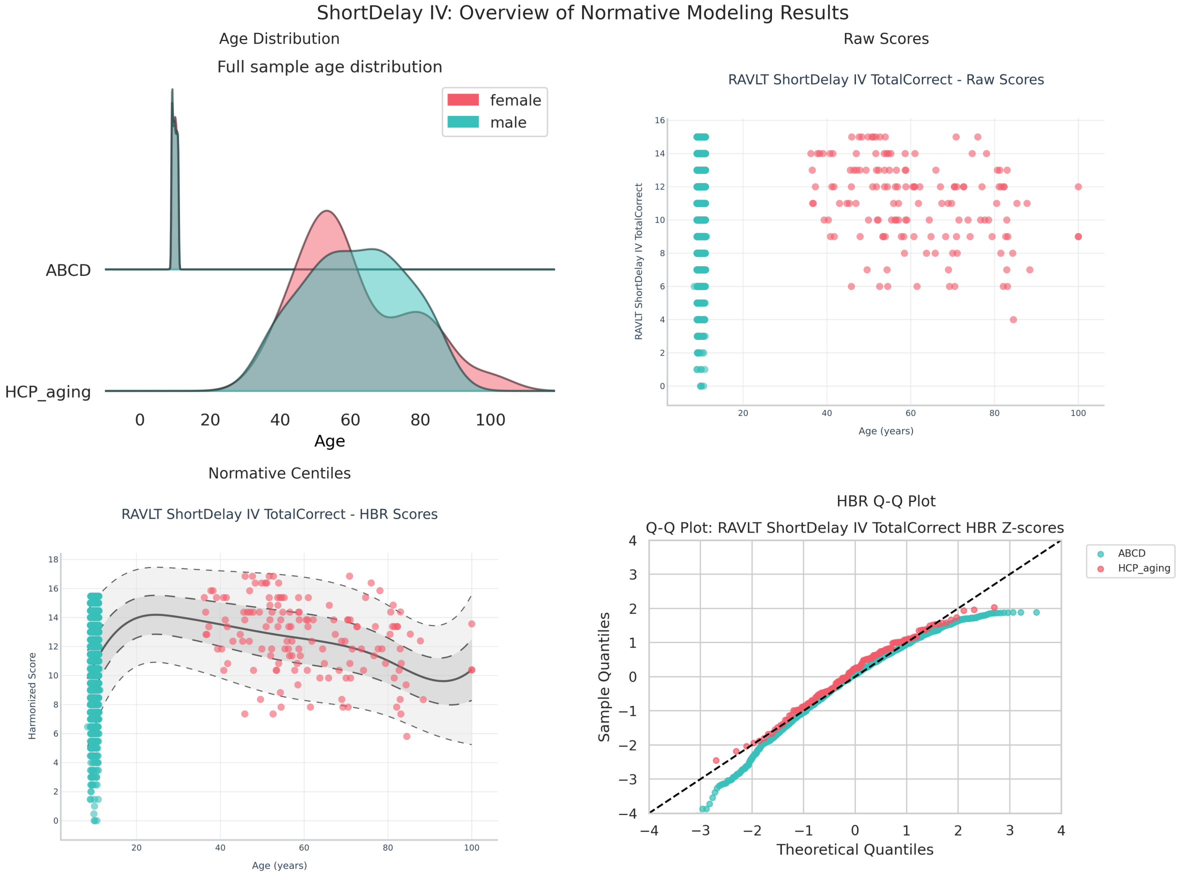


*Figure 14 Cognitive reference charts for the RAVLT Short Delay test trial IV. The top panel shows the sample age distribution and the scatter plot of the raw scores by age. The bottom panel shows the HBR deviation scores, also with a scatter plot and the Q-Q plot.*

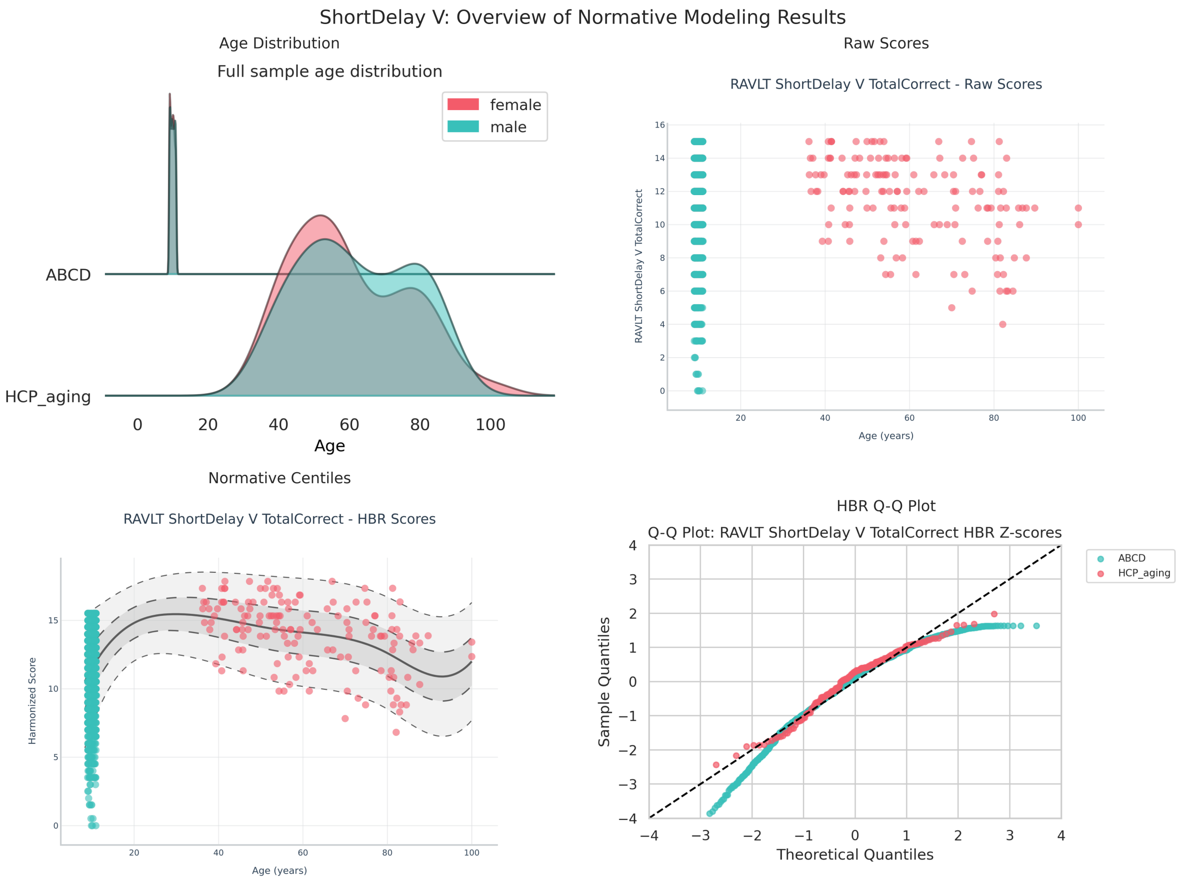


*Figure 15 Cognitive reference charts for the RAVLT Short Delay test trial V. The top panel shows the sample age distribution and the scatter plot of the raw scores by age. The bottom panel shows the HBR deviation scores, also with a scatter plot and the Q-Q plot.*

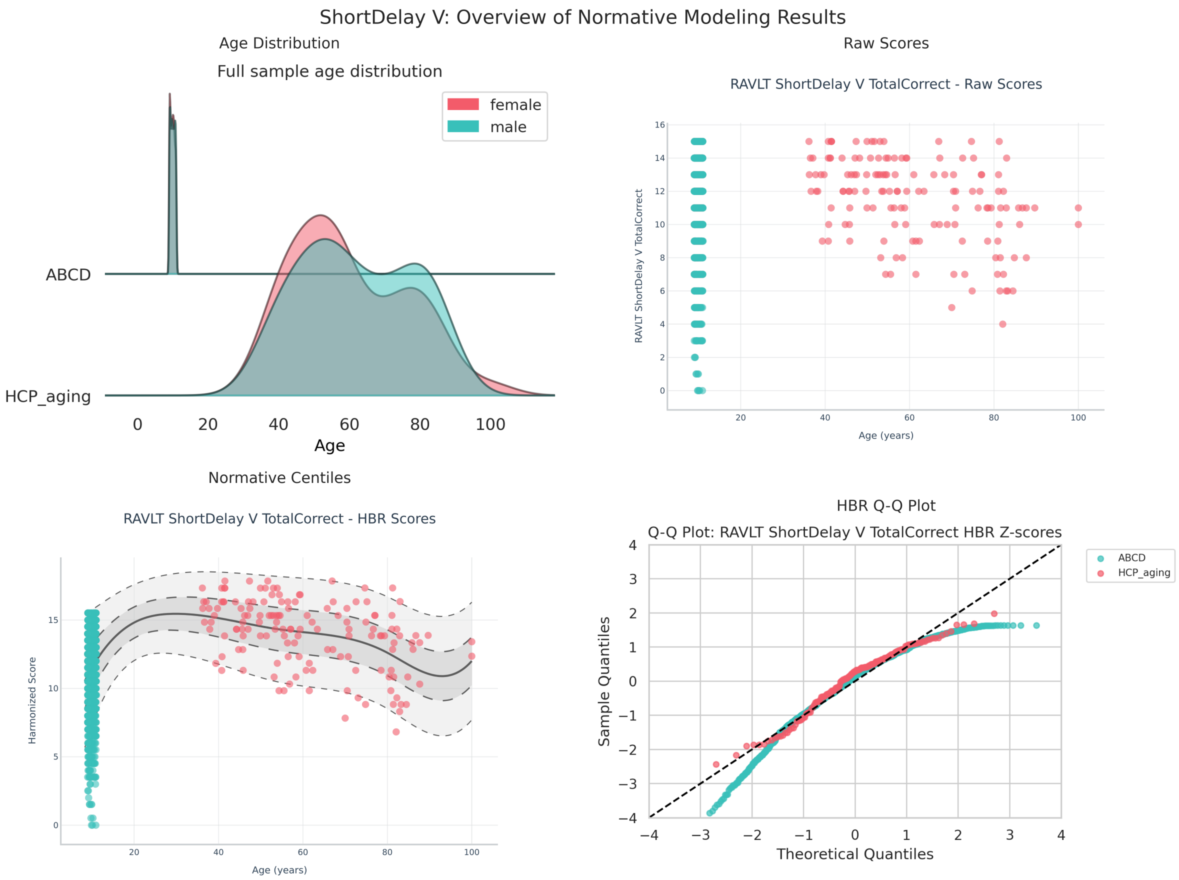


*Figure 16 Cognitive reference charts for the RAVLT Short Delay test trial VI. The top panel shows the sample age distribution and the scatter plot of the raw scores by age. The bottom panel shows the HBR deviation scores, also with a scatter plot and the Q-Q plot.*

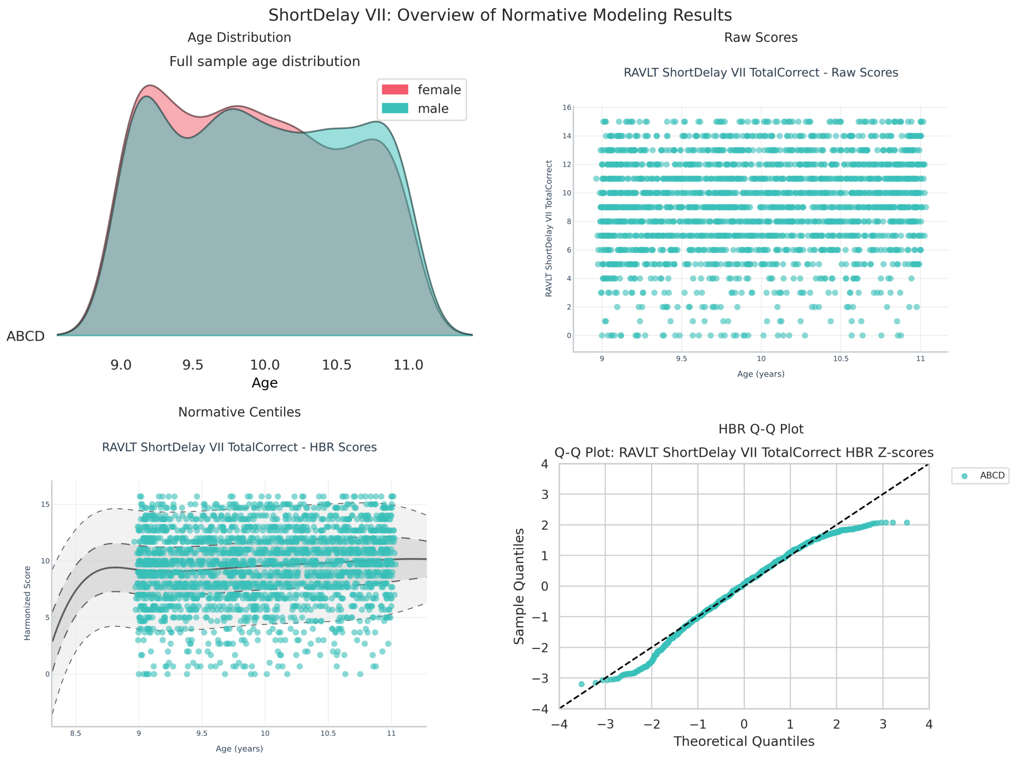


*Figure 17 Cognitive reference charts for the RAVLT Short Delay test trial VII. The top panel shows the sample age distribution and the scatter plot of the raw scores by age. The bottom panel shows the HBR deviation scores, also with a scatter plot and the Q-Q plot.*

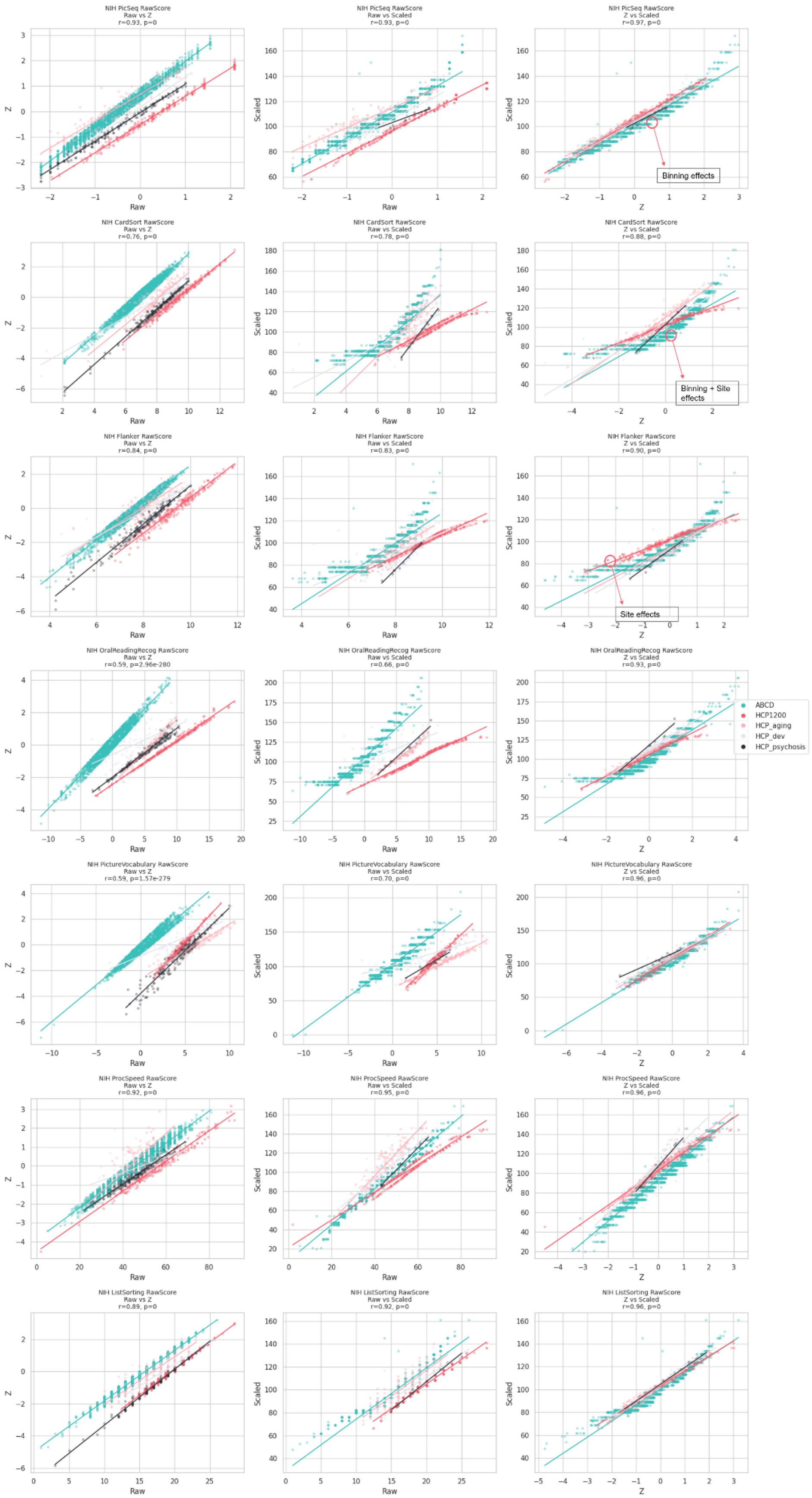


*Figure 18 Pairwise comparisons between raw, HBR z-standardised, and scaled scores across all cognitive measures included in the dataset. Each panel shows the relationship between two scoring formats for a given cognitive measure. Pearson correlation coefficients (r) and associated p-values are reported in each panel.*

**Model comparison results**

To create the cognitive reference charts, a wide range of normative model settings can be selected and optimized to achieve the best model fit, balancing model fit with model complexity. In general, we aim to select the simplest model that can still adequately capture the shape of the data distribution, including skewness, kurtosis, and site effects. Here, we compare seven different modeling settings: a Bayesian Linear Regression method[16], which is preferred for fast optimization; a Hierarchical Bayesian Regression (HBR) model using a Normal likelihood, which is appropriate for modeling data that are approximately normally distributed; an HBR model using a SHASHb likelihood, which is suitable for modeling data that are skewed or heavy-tailed; and an HBR model using a Beta likelihood, which is appropriate for modeling bounded data (e.g., between 0 and 1). For more details and the preferred hyperparameter starting values, see the accompanying reference papers[17,18].

In general, the likelihood parameters must be configured; here, we used the default settings[17]. Briefly, the hierarchical Bayesian regression with a Normal likelihood used linear hierarchical priors on the mean and variance, including random intercepts for batch effects (site) and a B-spline basis expansion to capture nonlinear age effects. The variance parameters were mapped via a softplus function to ensure positivity. The HBR model with a SHASHb likelihood extended the Normal HBR model by allowing skewness and heavy tails through the additional $\epsilon$ and $\delta$ parameters. The final setting we evaluated was the Beta-HBR model for bounded responses, in which $\alpha$ and $\beta$ parameters are introduced. All HBR models were sampled using the NUTS algorithm with four chains, 1,500 draws, 500 tuning steps, and 16 CPU cores.

In the figures below, we show the results of the different model comparisons with the settings. The BLR model included fixed effects for the batch variables, heteroskedastic noise, and a warping for the likelihood using a sin-arcsinh transformation accounting for non-Gaussian distribution[16]. The HBR models were designed to account for batch effects across sites. Specifically, we estimated a Gaussian HBR model, a Gaussian HBR model with hierarchical variance (random σ), a sinh–arcsinh (SHASHb) likelihood HBR model that allows flexible modelling of skewness and kurtosis, and a Beta HBR model for bounded outcome variables. Full details on these likelihood formulations, their parameterizations, and their advantages can be found in the accompanying papers[16–19] , and details of the exact implementation of these methods can also be found in the accompanying code.

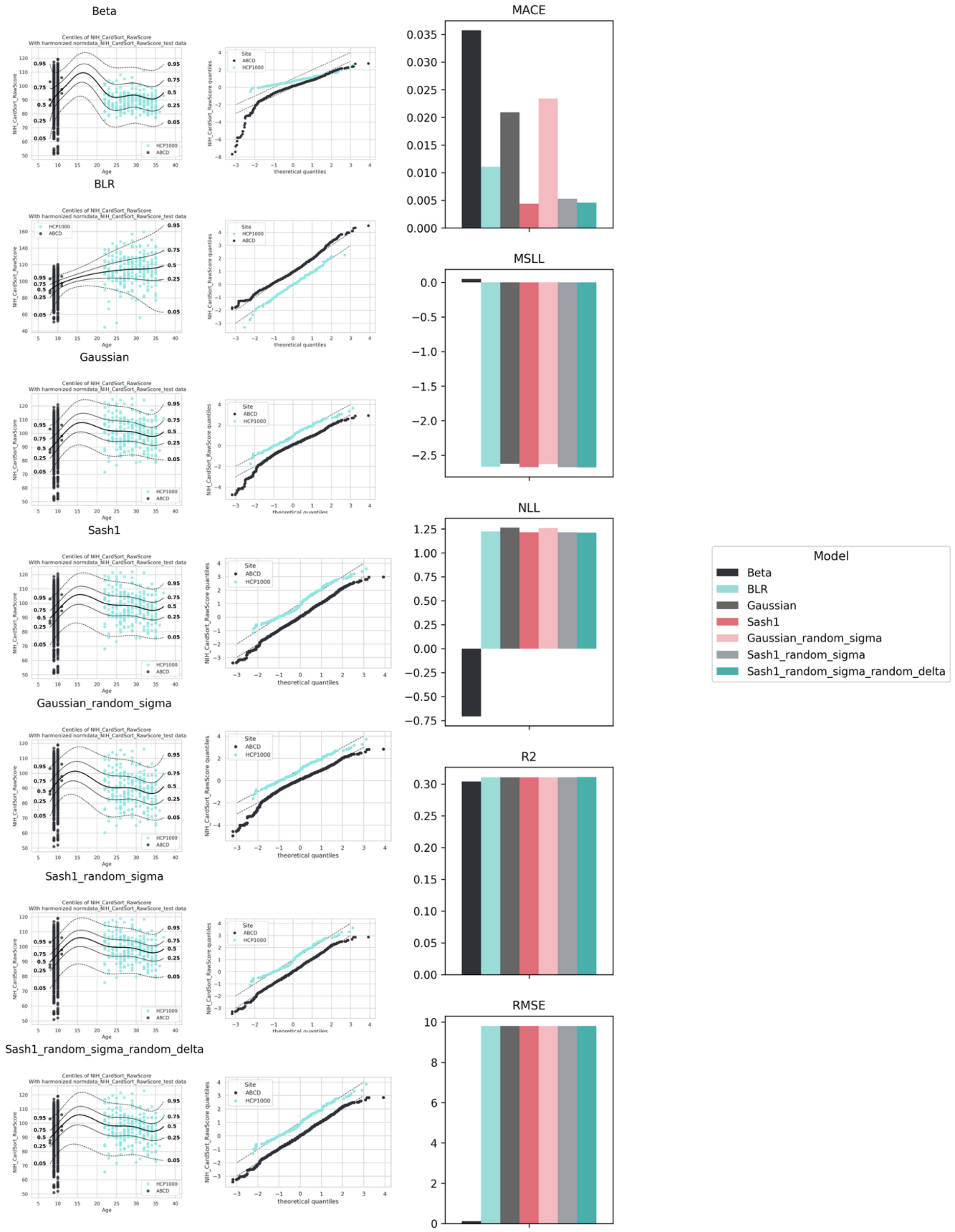


*Figure 18 Model comparison for the NIH Cardsort test. For each model in the left column, the centile plot is shown. In the middle column, the corresponding QQ plot is demonstrated, assessing the distributional fit. The rightmost column summarizes the predictive performance metrics across all models using (MACE, MSLL, NLL, $R^{2,}$ and the RMSE). Bars represent metric values for each model, enabling direct comparison of calibration, goodness-of-fit, and predictive accuracy.*

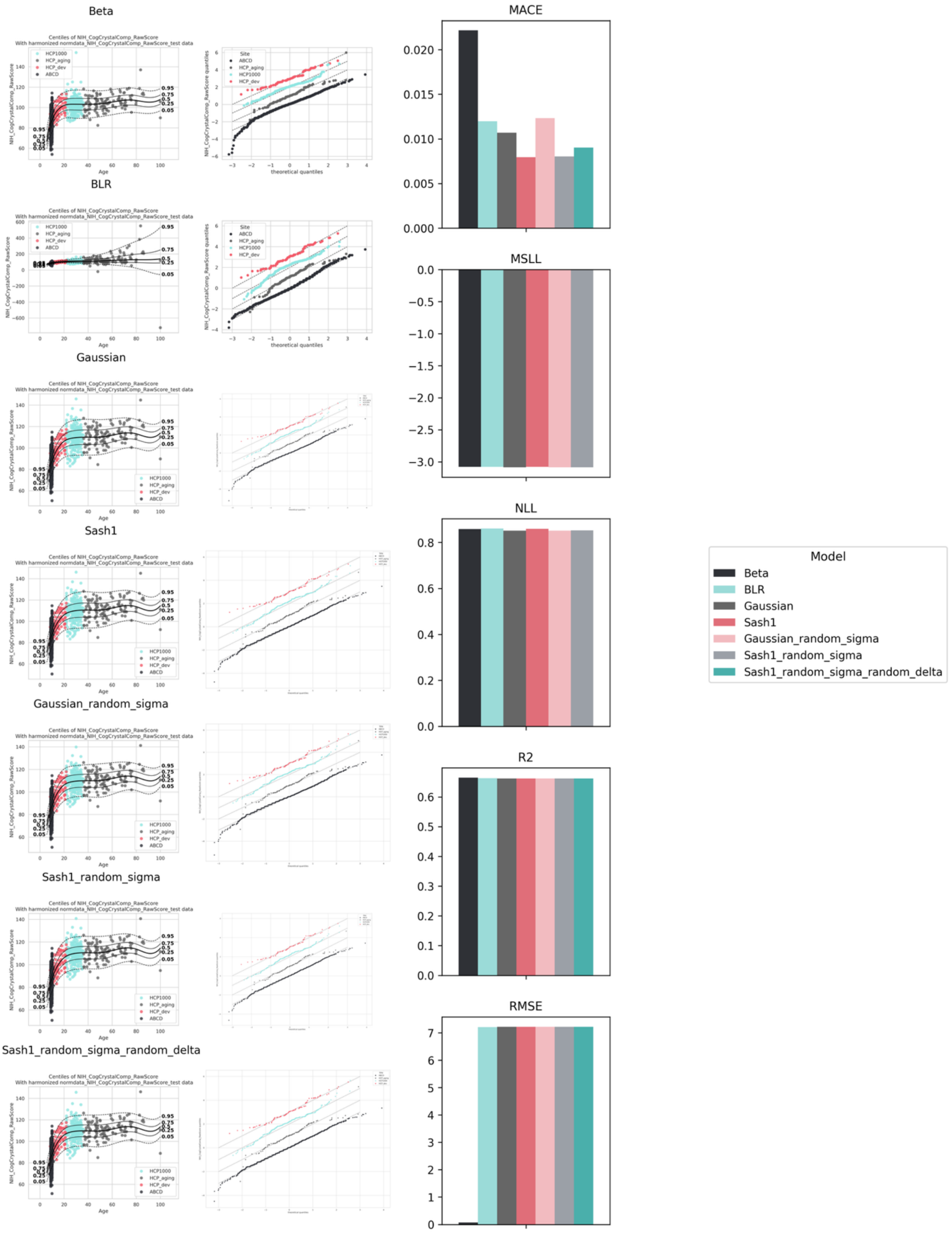


*Figure 20 Model comparison for the NIH Crystallized Cognitive Composite score. For each model in the left column, the centile plot is shown. In the middle column, the corresponding QQ plot is demonstrated, assessing the distributional fit. The rightmost column summarizes the predictive performance metrics across all models using (MACE, MSLL, NLL, $R^{2,}$ and the RMSE). Bars represent metric values for each model, enabling direct comparison of calibration, goodness-of-fit, and predictive accuracy.*

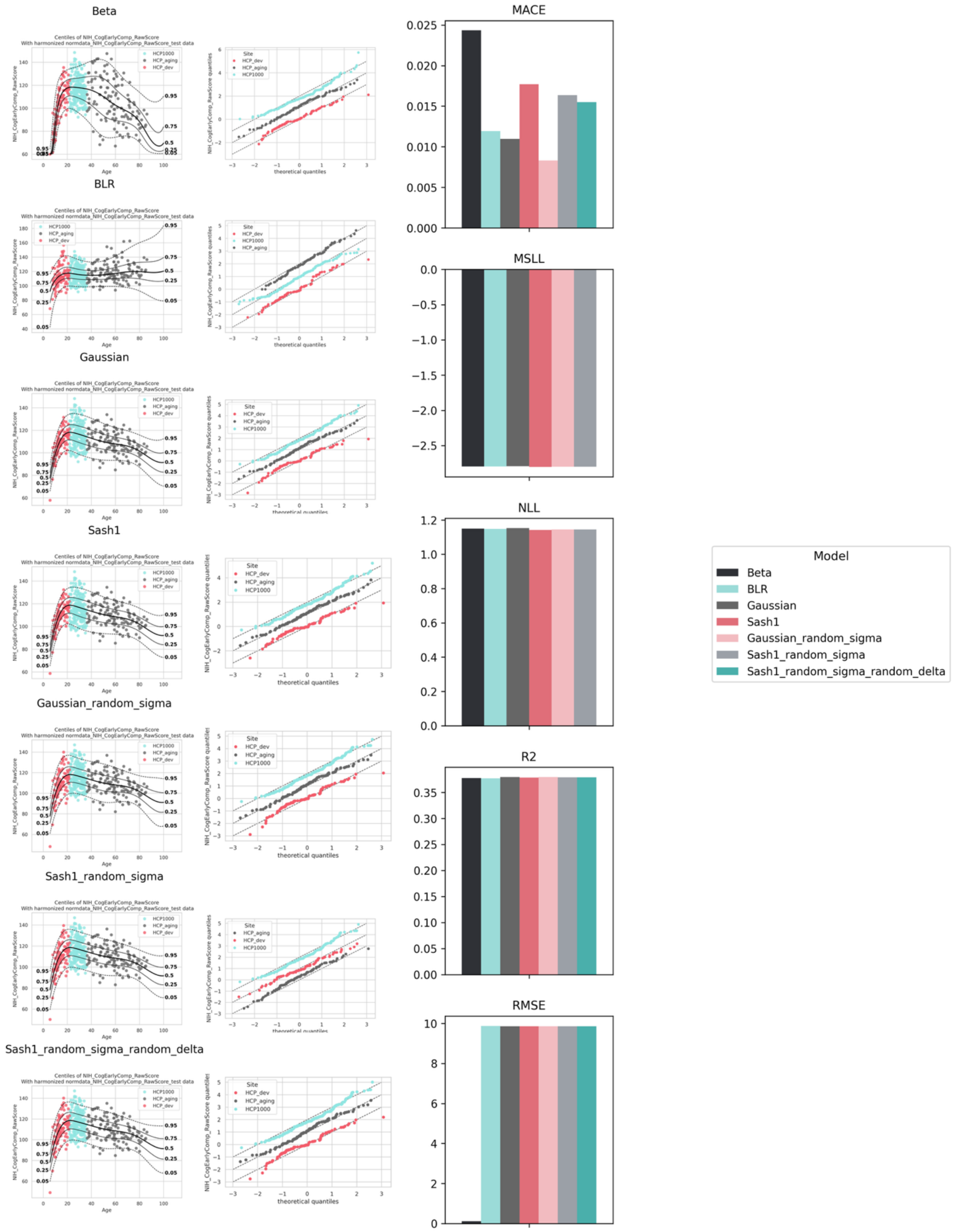


*Figure 19 Model comparison for the NIH Early Cognitive Composite Scores. For each model in the left column, the centile plot is shown. In the middle column, the corresponding QQ plot is demonstrated, assessing the distributional fit. The rightmost column summarizes the predictive performance metrics across all models using (MACE, MSLL, NLL, $R^{2,}$ and the RMSE). Bars represent metric values for each model, enabling direct comparison of calibration, goodness-of-fit, and predictive accuracy.*

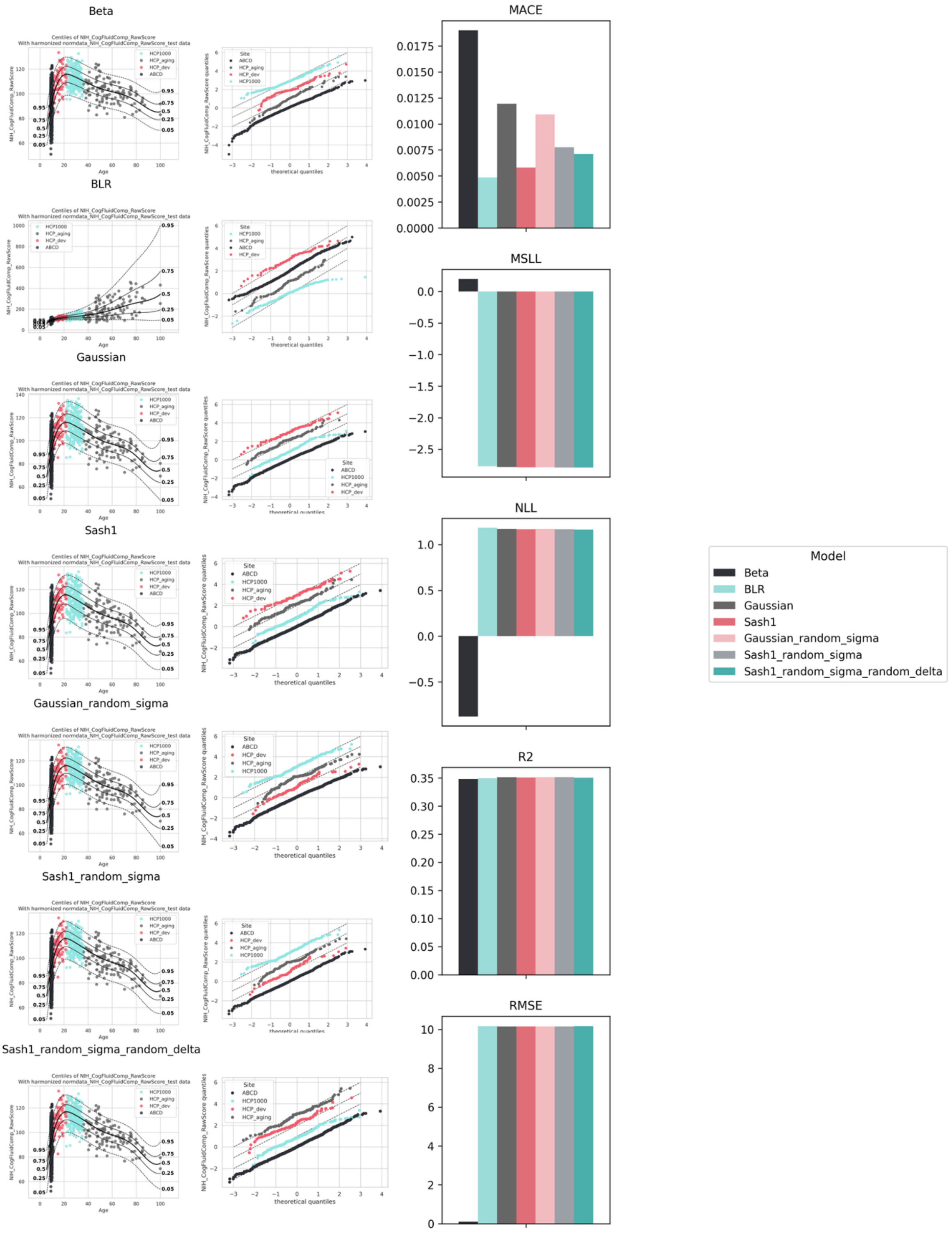


*Figure 20 Model comparison for the NIH Fluid Cognitive Composite Scores. For each model in the left column, the centile plot is shown. In the middle column, the corresponding QQ plot is demonstrated, assessing the distributional fit. The rightmost column summarizes the predictive performance metrics across all models using (MACE, MSLL, NLL, $R^{2}$, and the RMSE). Bars represent metric values for each model, enabling direct comparison of calibration, goodness-of-fit, and predictive accuracy.*

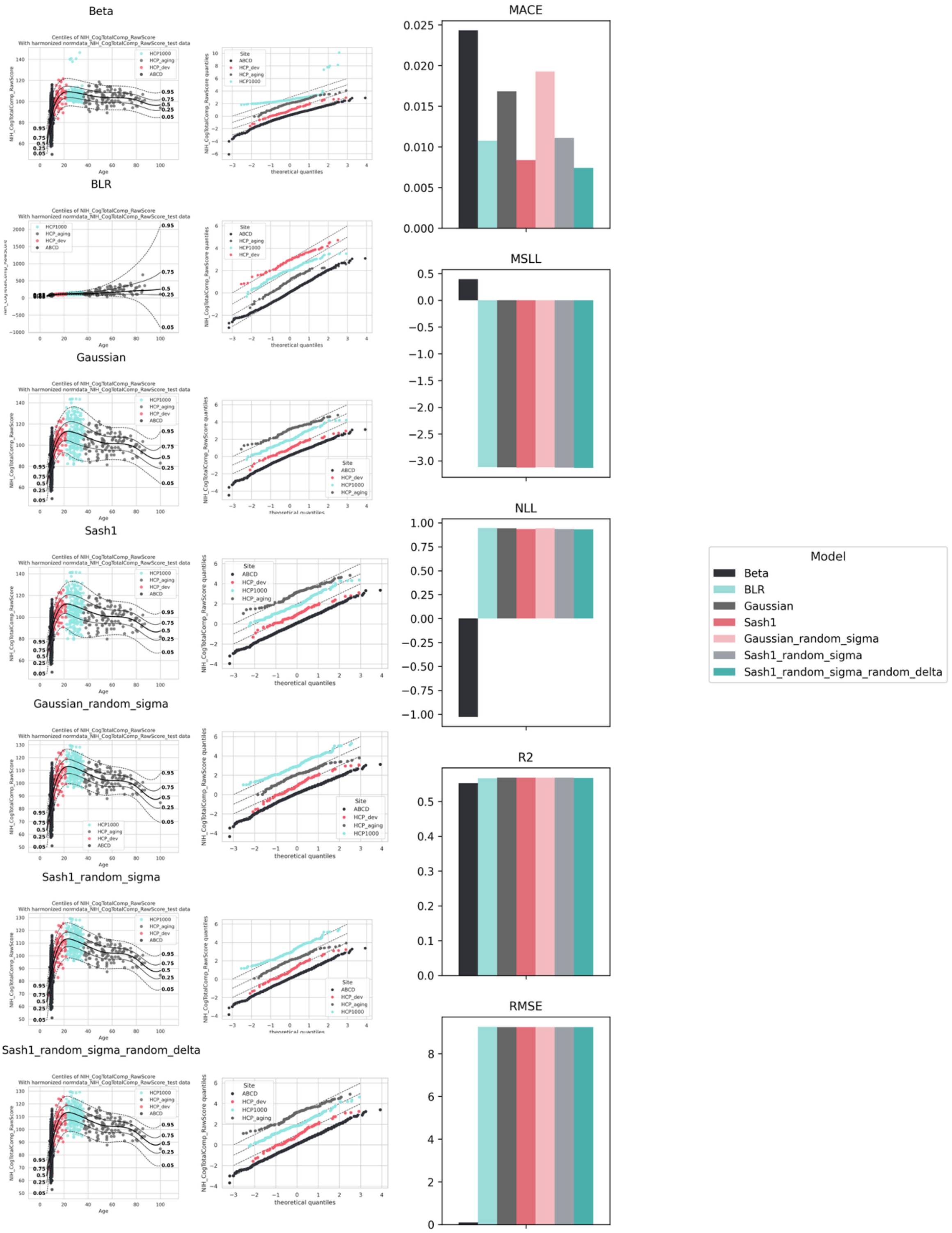


*Figure 21 Model comparison for the NIH Total Cognitive Composite Scores. For each model in the left column, the centile plot is shown. In the middle column, the corresponding QQ plot is demonstrated, assessing the distributional fit. The rightmost column summarizes the predictive performance metrics across all models using (MACE, MSLL, NLL, $R^{2,}$ and the RMSE). Bars represent metric values for each model, enabling direct comparison of calibration, goodness-of-fit, and predictive accuracy.*

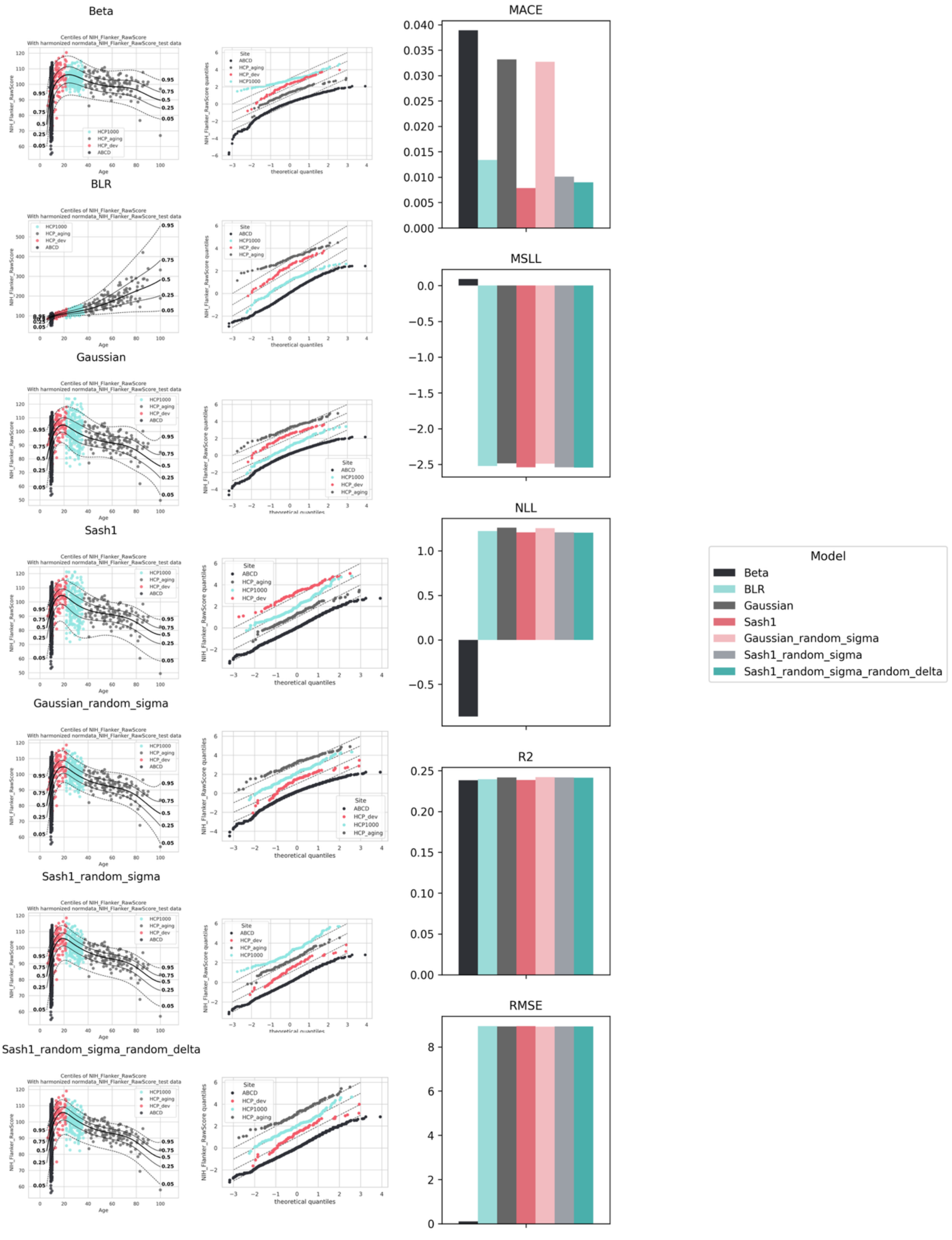


*Figure 22 Model comparison for the NIH Flanker Scores. For each model in the left column, the centile plot is shown. In the middle column, the corresponding QQ plot is demonstrated, assessing the distributional fit. The rightmost column summarizes the predictive performance metrics across all models using (MACE, MSLL, NLL, $R^{2,}$ and the RMSE). Bars represent metric values for each model, enabling direct comparison of calibration, goodness-of-fit, and predictive accuracy.*

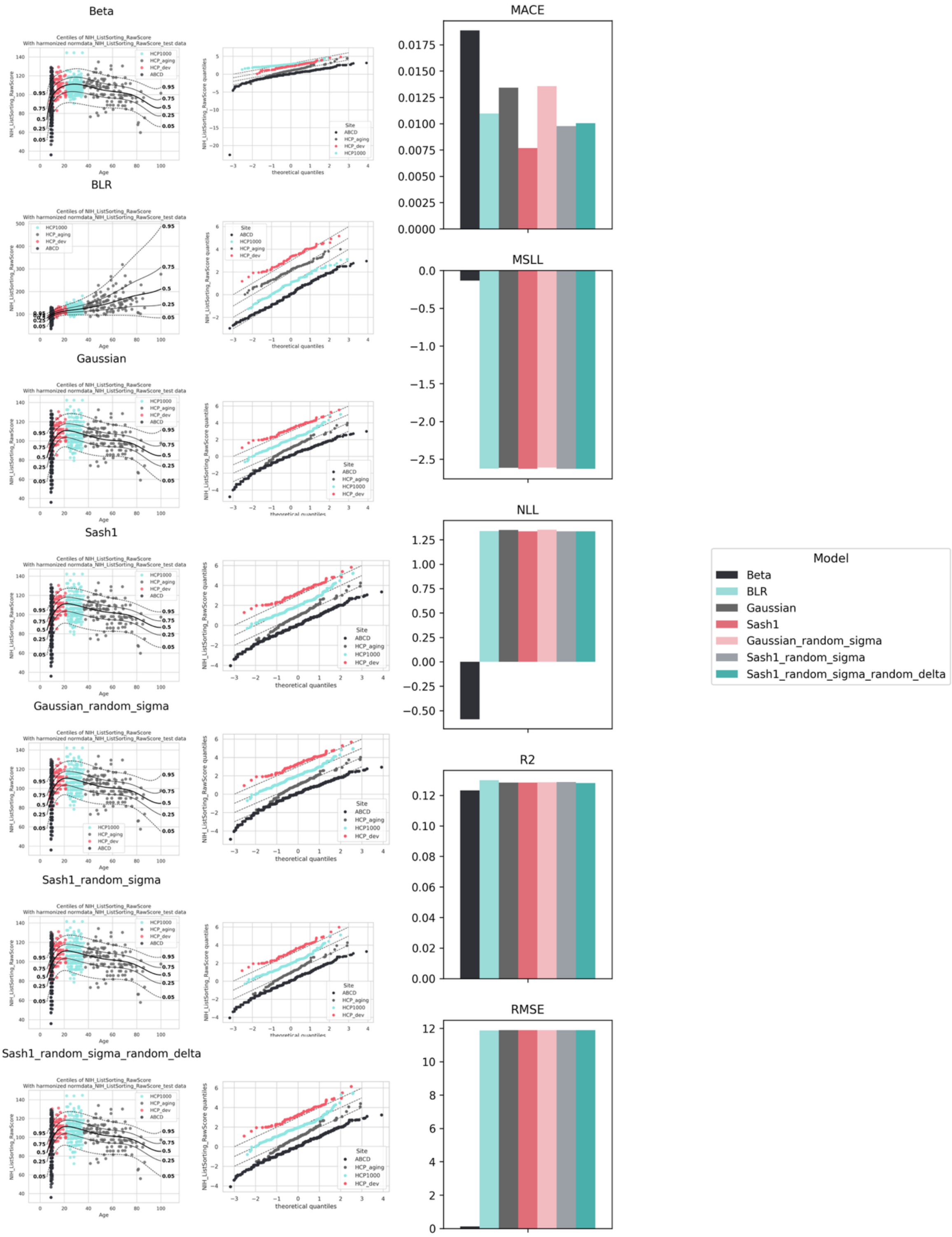


*Figure 23 Model comparison for the NIH List Sorting Scores. For each model in the left column, the centile plot is shown. In the middle column, the corresponding QQ plot is demonstrated, assessing the distributional fit. The rightmost column summarizes the predictive performance metrics across all models using (MACE, MSLL, NLL, $R^2$, and the RMSE). Bars represent metric values for each model, enabling direct comparison of calibration, goodness-of-fit, and predictive accuracy.*

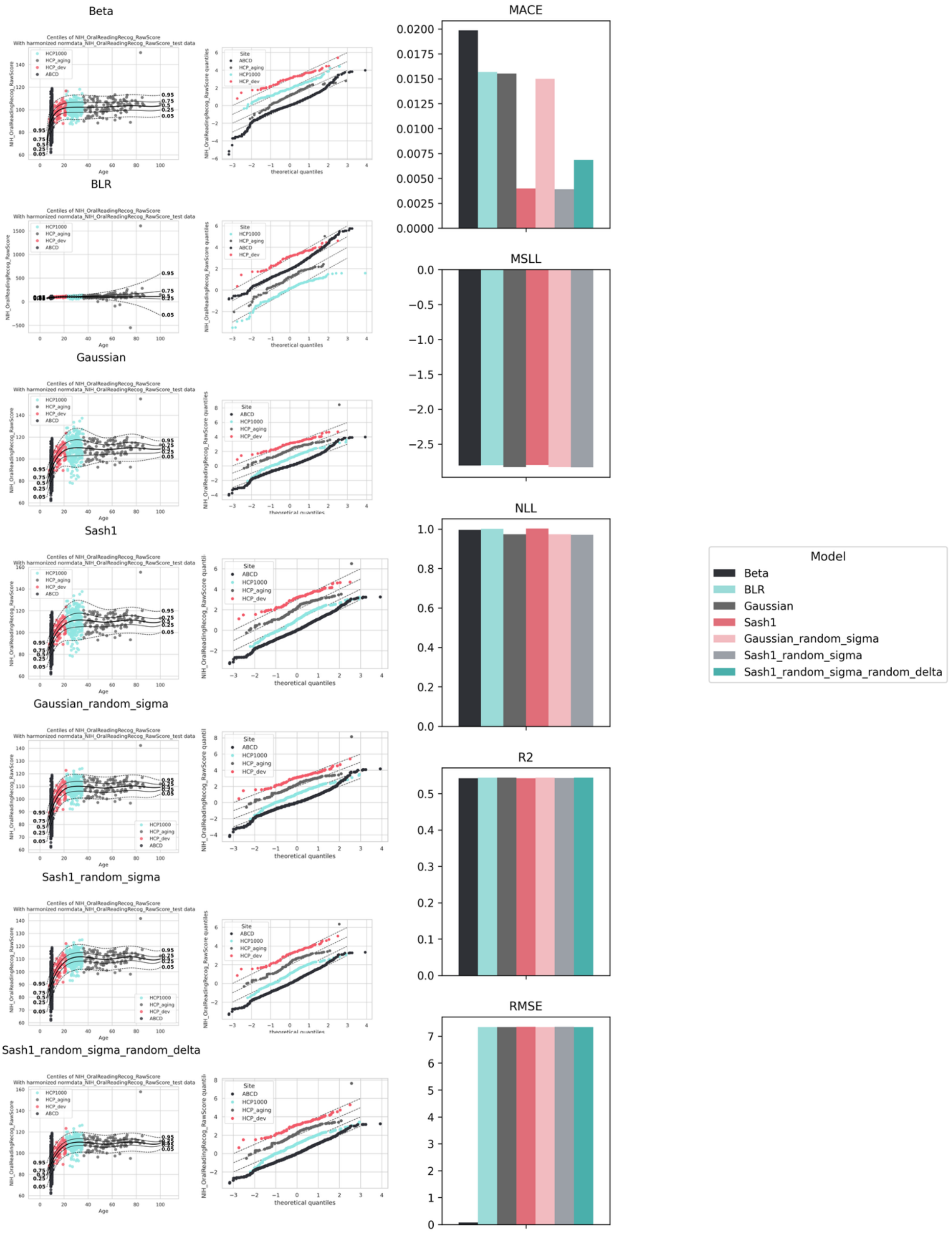


*Figure 24 Model comparison for the NIH Oral Reading Recognition Scores. For each model in the left column, the centile plot is shown. In the middle column, the corresponding QQ plot is demonstrated, assessing the distributional fit. The rightmost column summarizes the predictive performance metrics across all models using (MACE, MSLL, NLL, $R^{2,}$ and the RMSE). Bars represent metric values for each model, enabling direct comparison of calibration, goodness-of-fit, and predictive accuracy.*

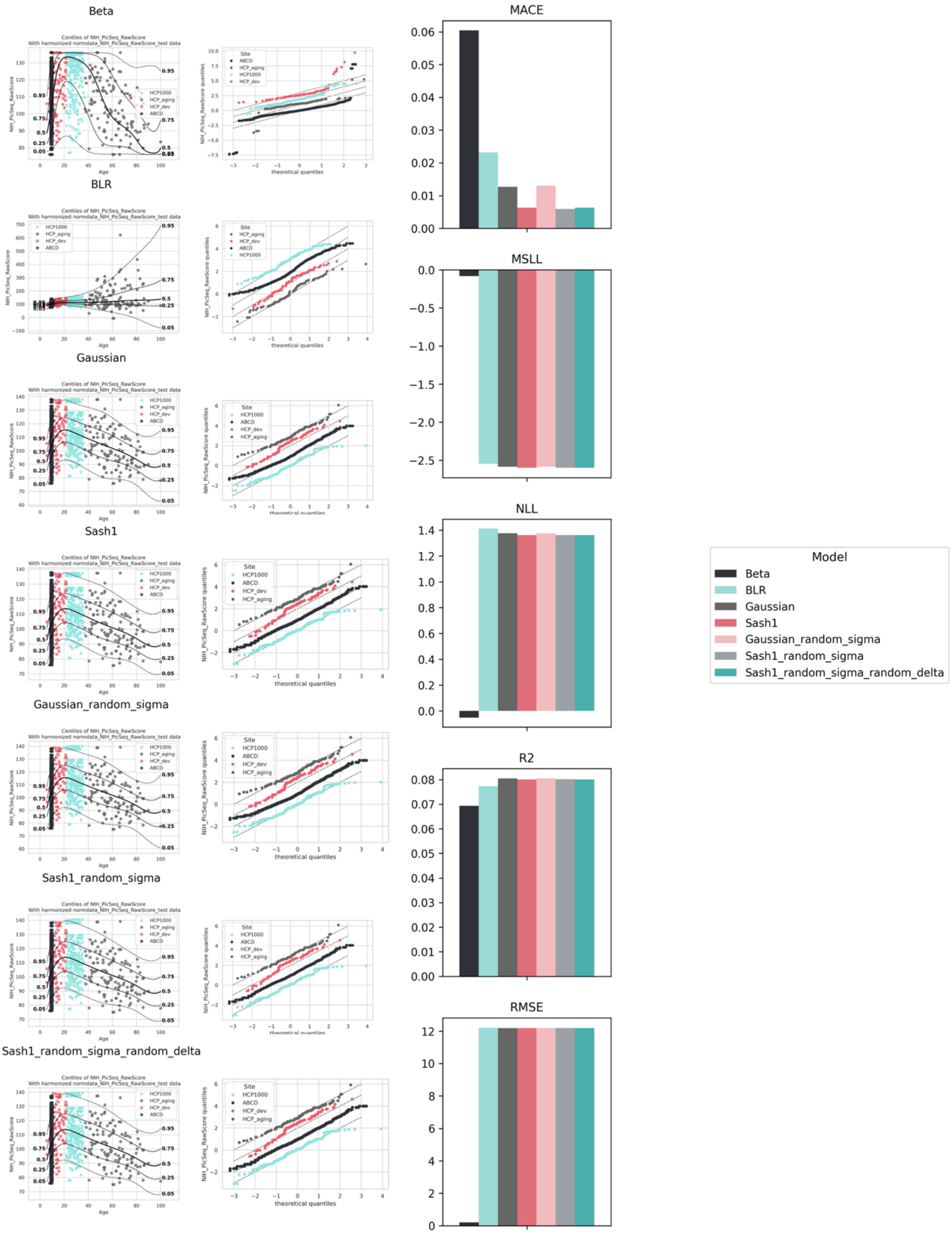


*Figure 25 Model comparison for the NIH Picture Sequence Scores. For each model in the left column, the centile plot is shown. In the middle column, the corresponding QQ plot is demonstrated, assessing the distributional fit. The rightmost column summarizes the predictive performance metrics across all models using (MACE, MSLL, NLL, $R^2$, and the RMSE). Bars represent metric values for each model, enabling direct comparison of calibration, goodness-of-fit, and predictive accuracy.*

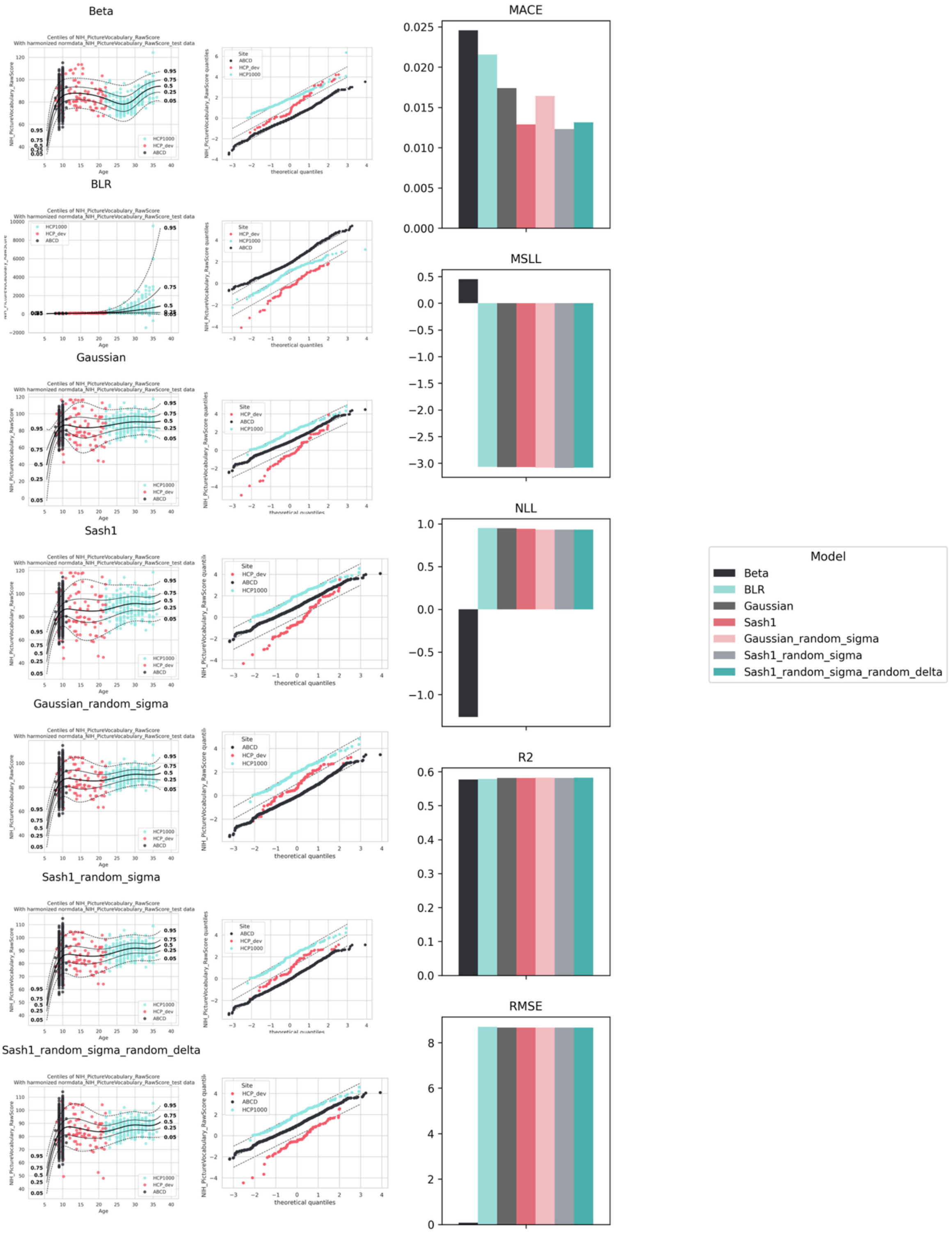


*Figure 26 Model comparison for the NIH Picture Vocabulary Scores. For each model in the left column, the centile plot is shown. In the middle column, the corresponding QQ plot is demonstrated, assessing the distributional fit. The rightmost column summarizes the predictive performance metrics across all models using (MACE, MSLL, NLL, $R^2$, and the RMSE). Bars represent metric values for each model, enabling direct comparison of calibration, goodness-of-fit, and predictive accuracy.*

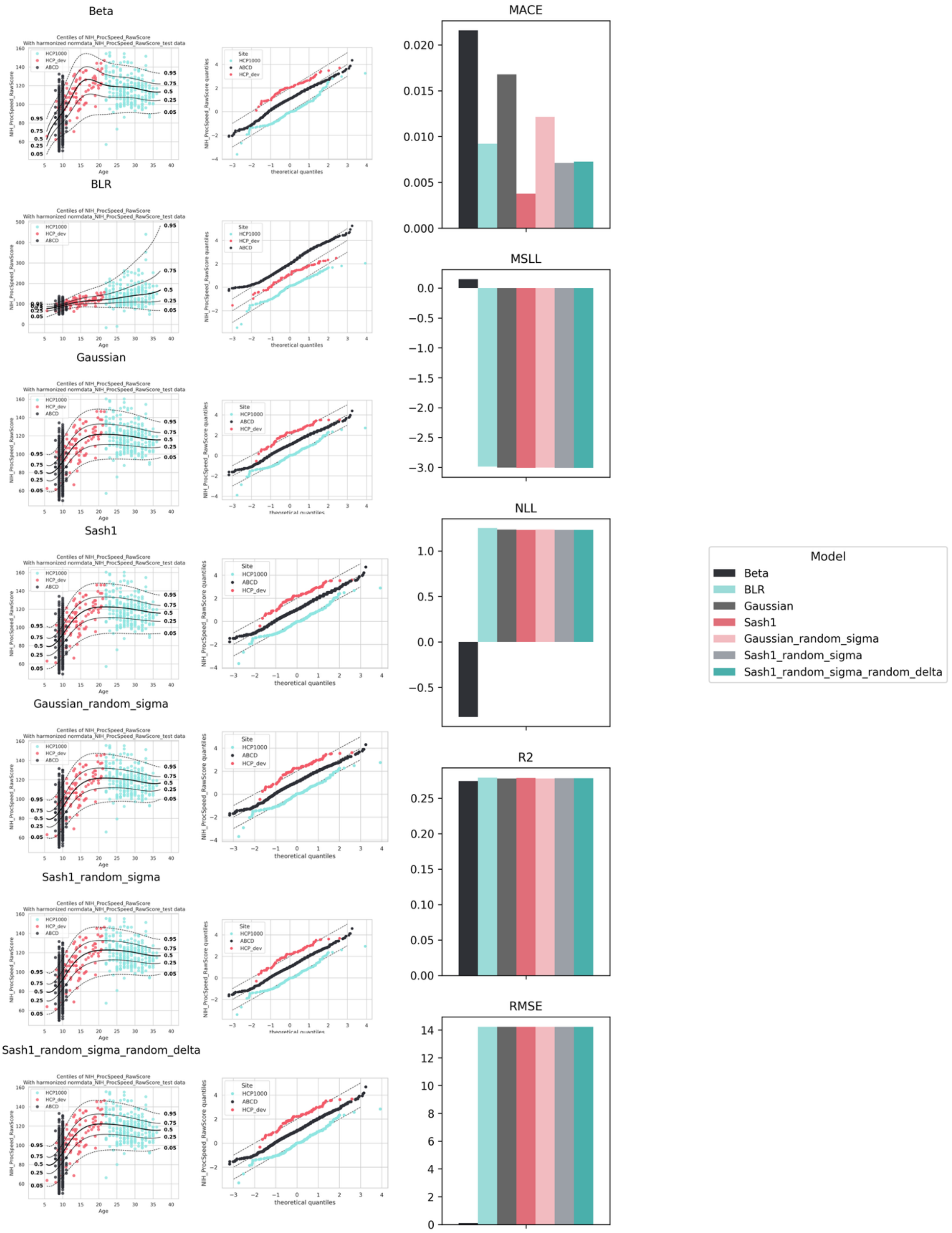


*Figure 27 Model comparison for the NIH Processing Speed Scores. For each model in the left column, the centile plot is shown. In the middle column, the corresponding QQ plot is demonstrated, assessing the distributional fit. The rightmost column summarizes the predictive performance metrics across all models using (MACE, MSLL, NLL, $R^2$, and the RMSE). Bars represent metric values for each model, enabling direct comparison of calibration, goodness-of-fit, and predictive accuracy.*

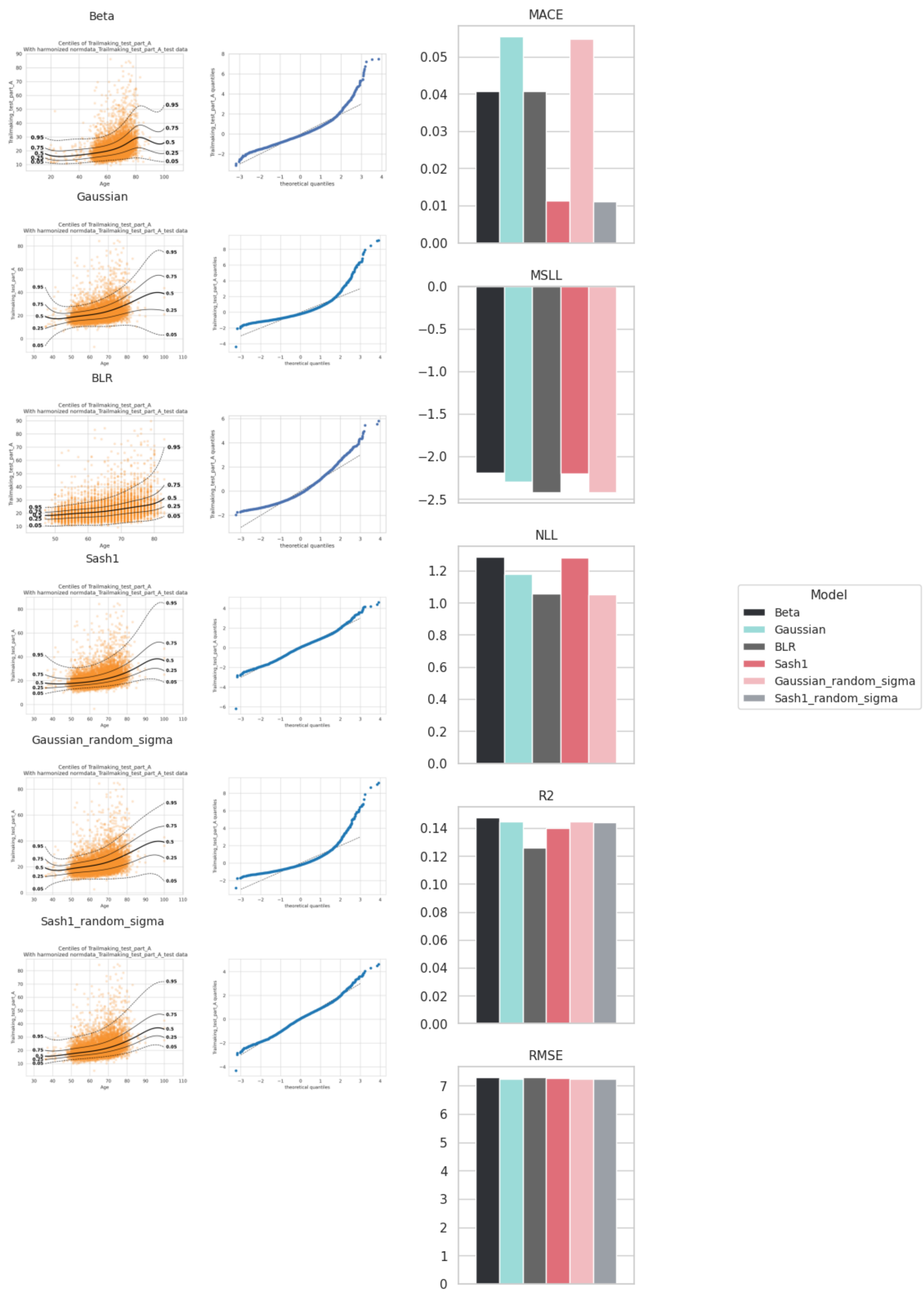


*Figure 30 Model comparison for the Trailmaking Test Part A Scores. For each model in the left column, the centile plot is shown. In the middle column, the corresponding QQ plot is demonstrated, assessing the distributional fit. The rightmost column summarizes the predictive performance metrics across all models using (MACE, MSLL, NLL, $R^{2}$, and the RMSE). Bars represent metric values for each model, enabling direct comparison of calibration, goodness-of-fit, and predictive accuracy.*

### Federated learning transfer results

There are two types of federated learning functions available for cognitive reference charts: extend and transfer, for local adaptation. First, for the extend function, the reference model is updated with the newly learned information, creating a model that can generate predictions for the local dataset, as well as the original data[18]. For the transfer analysis, the reference model can be transferred to held-out data, producing smaller, site-specific models that retain partial knowledge of the reference model through their priors.

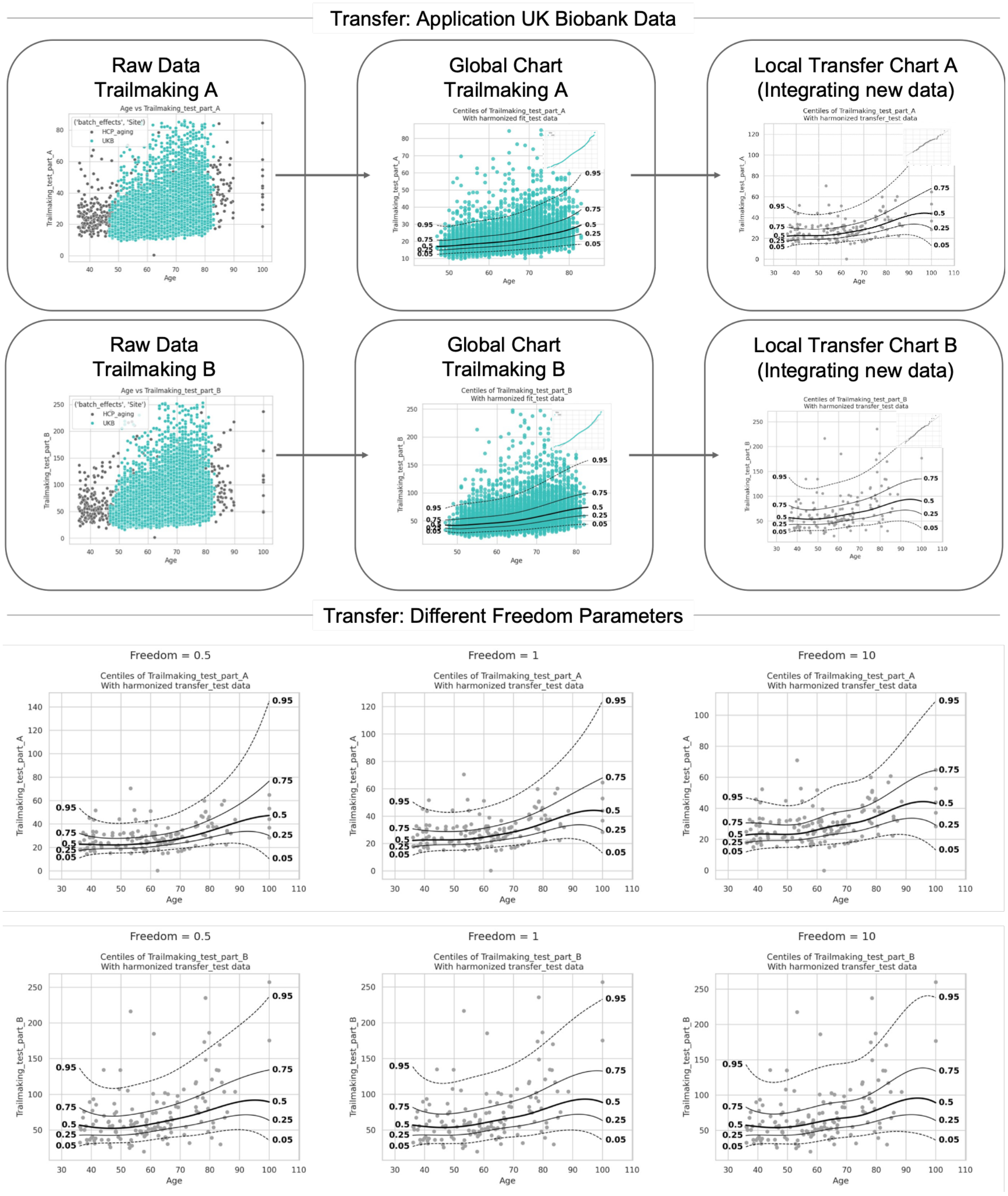


*Figure 28 Comparison of transfer learning freedom parameters. At the top, the results of the transferred models for the Trail Making Tests A and B are shown. The first panel shows scatterplots of the raw data for UKB and HCP-Aging. The second panel shows the estimated cognitive reference charts for the two tests based on the UKB dataset. The third panel shows the transferred models incorporating the additional HCP aging dataset. At the bottom, the centile*

*plots are shown for Trail Making Test A and B, obtained after transferring a pre-trained HBR normative model to the HCP aging dataset using three values of the transfer freedom parameter (0.5, 1, and 10), with freedom < 1 enforcing a tighter prior, and freedom > 1 allowing greater deviation from the reference model. We can observe that for a Freedom parameter of 10, the model becomes overly flexible to the new dataset.*

1. Normative Framework: No Transfer Applied

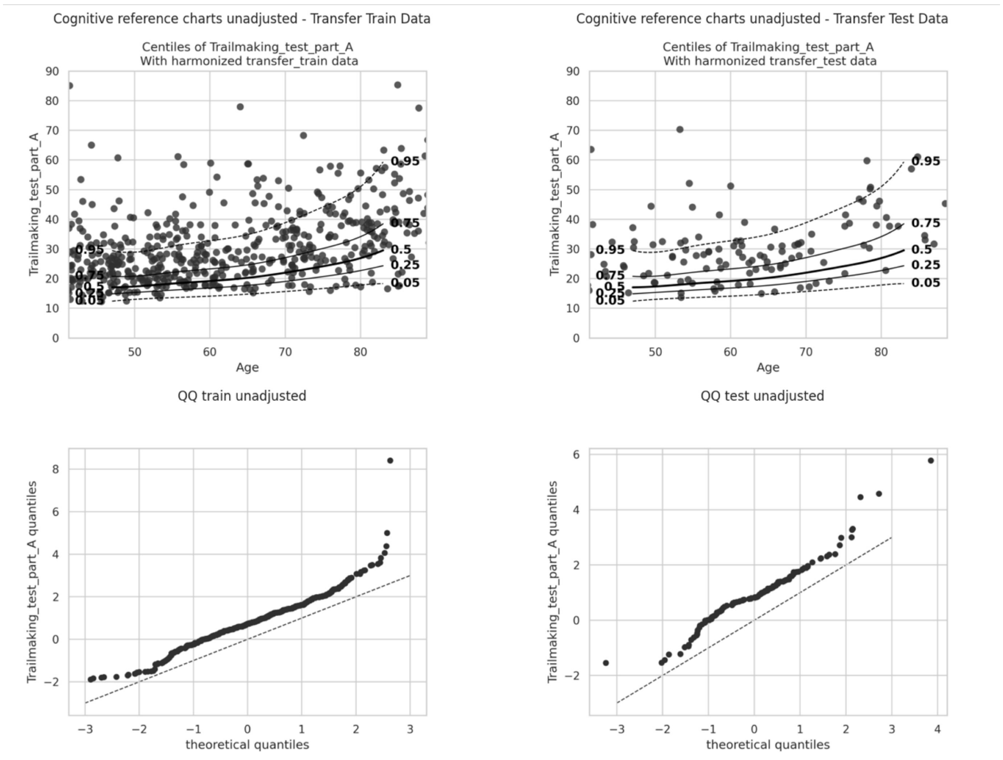


2. Normative Framework: Transfer Applied

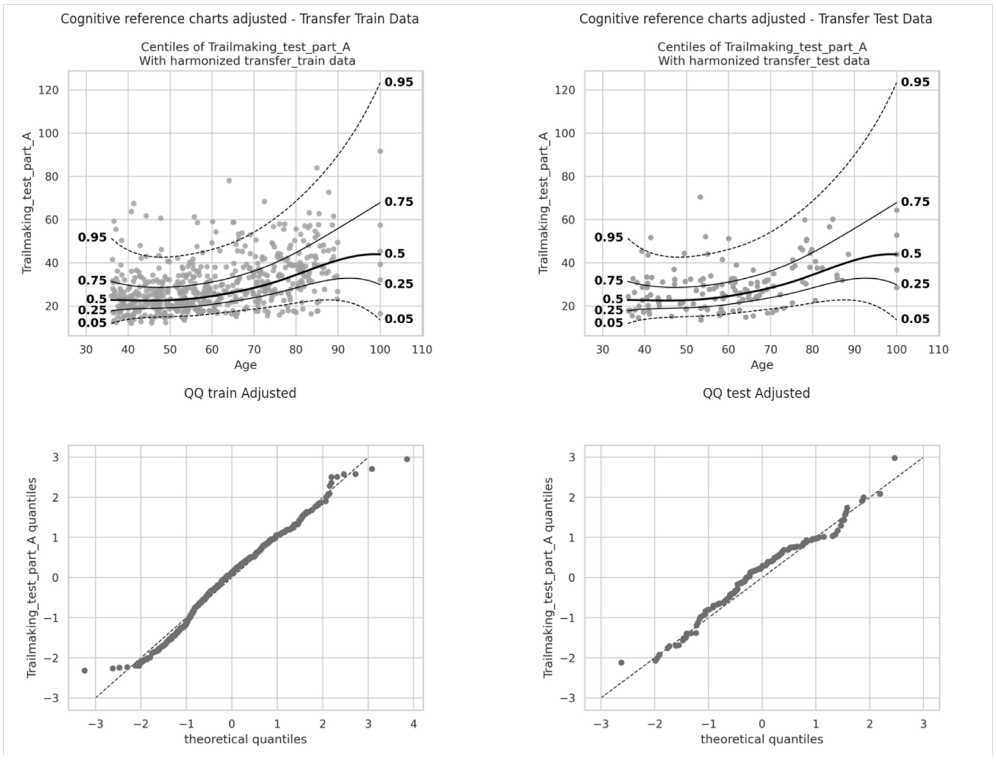


*Figure 29 Evaluation of normative model performance in the HCP-Aging cohort before and after transfer learning learning for Trail Making Test part A. (1) Application of the UK Biobank-trained normative model to HCP-Aging data without site adaptation. (2) Application of the same model following transfer learning to adjust for site-specific effects using HCP-Aging transfer-training data. For each panel, centile plots (top) and QQ plots (bottom) are shown for the transfer-training (left) and transfer-test (right) datasets. We can observe that for the untransferred model application site effects remain.*

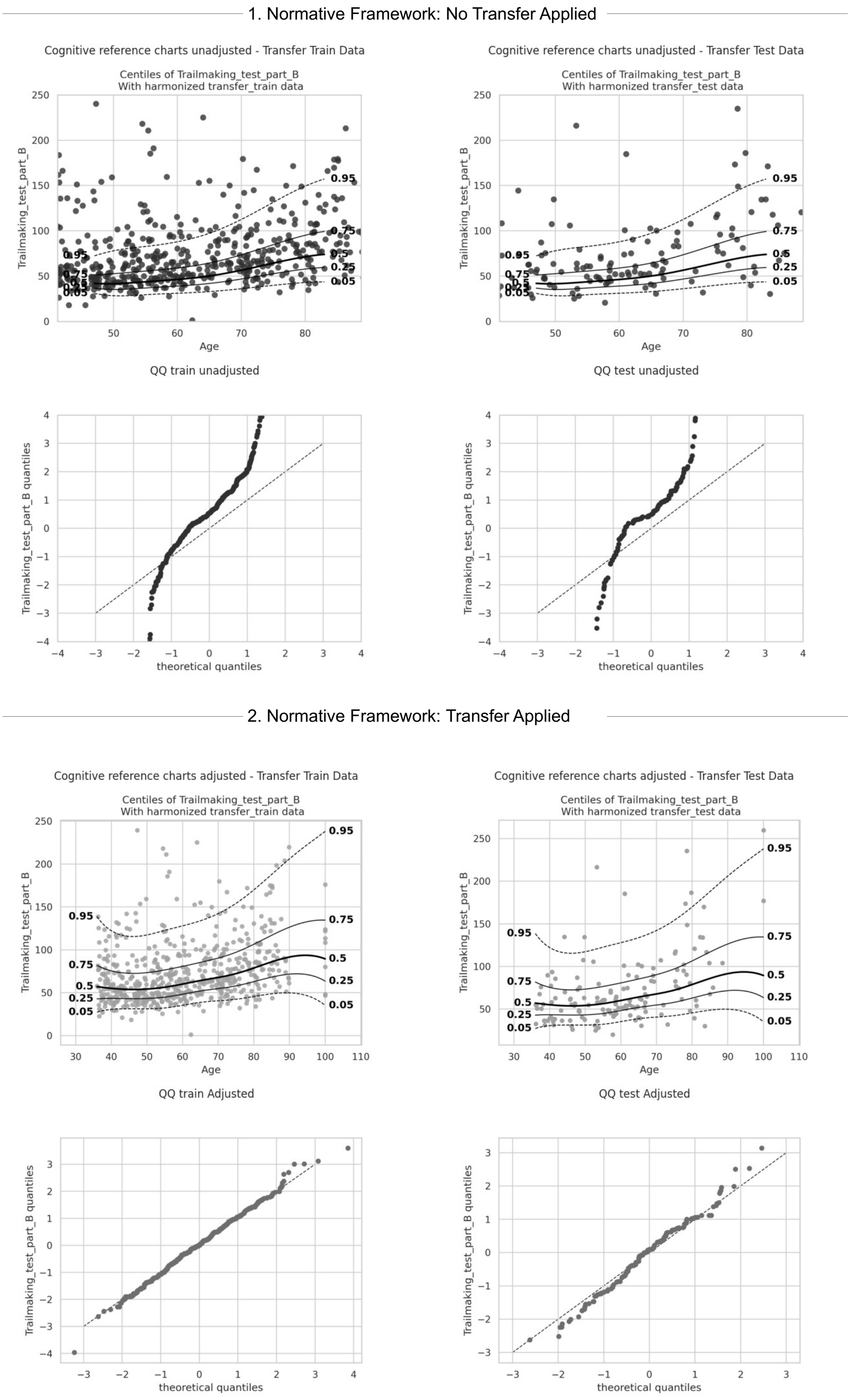


*Figure 30 Evaluation of normative model performance in the HCP-Aging cohort before and after transfer learning for Trail Making Test part B. (1) Application of the UK Biobank-trained normative model to HCP-Aging data without site adaptation. (2) Application of the same model following transfer learning to adjust for site-specific effects using HCP-Aging transfer-training data. For each panel, centile plots (top) and QQ plots (bottom) are shown for the transfer-training (left) and transfer-test (right) datasets. We can observe that for the untransferred model application, the estimated deviation scores deviate highly from the expected Gaussian trend.*
